# Identification of plasma proteomic markers underlying polygenic risk of type 2 diabetes and related comorbidities

**DOI:** 10.1101/2024.03.15.24304200

**Authors:** Douglas P. Loesch, Manik Garg, Dorota Matelska, Dimitrios Vitsios, Xiao Jiang, Scott C. Ritchie, Benjamin B. Sun, Heiko Runz, Christopher D. Whelan, Rury R. Holman, Robert J. Mentz, Filipe A. Moura, Stephen D. Wiviott, Marc S. Sabatine, Miriam S. Udler, Ingrid A. Gause-Nilsson, Slavé Petrovski, Jan Oscarsson, Abhishek Nag, Dirk S. Paul, Michael Inouye

## Abstract

**Introduction:** Type 2 diabetes (T2D) is a heterogeneous disorder for which disease-causing pathways are incompletely understood. Here, we mapped genetic risk for T2D and its comorbidities to proteins, mechanistic pathways and clinical outcomes using proteogenomic data from a population-scale biobank and two randomized controlled trials.

**Methods:** We tested polygenic scores (PGS) for T2D and its cardiometabolic comorbidities, plus five partitioned T2D PGS (beta cell, lipodystrophy, liver lipid, obesity, and liver lipid), for association with 2,922 circulating proteins in 54,306 multi-ancestry participants (of which 42,452 were unrelated and without prevalent cardiometabolic disease) from the UK Biobank (UKB). Then, we tested the PGS-associated proteins for association with incident cardiometabolic complications in two cardiovascular outcome trials among T2D patients with proteogenomic data: EXSCEL (N=2,823) and DECLARE-TIMI 58 (N=915). We assessed causality using two-sample Mendelian randomization and mediation.

**Results:** We identified 839 unique proteins significantly associated with any T2D PGS and 1,005 proteins that were associated with at least one cardiometabolic PGS. Some PGS-associated proteins such as TFF3, EFEMP1, and MMP12 were in turn associated with renal and cardiovascular trial outcomes. PGS association patterns revealed shared pathways, e.g., complement cascade, cholesterol metabolism, IGF signaling. The proteins underlying these pathways, such as LPA, C1S, and IGFBP2, were consistently associated with clinical trial outcomes or identified via causal inference.

**Conclusions:** This proteogenomic study revealed proteins and mechanistic pathways underlying T2D and related comorbidities, advancing our understanding of T2D pathobiology and identifying putative biomarkers. All our results are available in an online data portal (https://public.cgr.astrazeneca.com/t2d-pgs/v1/).

## Introduction

Diabetes mellitus is a complex, multifactorial metabolic disorder diagnosed via a single clinical feature, hyperglycaemia.^1,2^ Among the major diagnoses of diabetes mellitus, type 2 diabetes (T2D) has the largest worldwide disease burden.^1^ T2D is itself highly heterogenous, including individuals with different degrees of insulin resistance and beta-cell dysfunction.^3^ The vast majority of patients with T2D have at least one additional comorbidity, such as hypertension, obesity or hyperlipidaemia.^4^ Genome-wide association studies (GWAS) have identified genetic associations that have advanced our understanding of pathophysiological pathways underlying T2D,^5,6^ though connecting individual T2D risk variants to specific pathways remains challenging. Analytical strategies to understand the heterogenous nature of T2D have included performing GWAS within clinically-defined T2D clusters^7^, or performing downstream clustering of genetic variants identified in GWAS for a broad T2D patient population.^5^

Many genetic variants associated with T2D that have been identified through GWAS have small effect sizes and map to non-protein-coding regions where their impact on biological pathways is not directly apparent. Polygenic risk scores (PRS), also known as polygenic scores (PGS), aggregate the small effects of these variants, and support risk prediction and stratification, including for T2D.^8,9^ Partitioned polygenic scores (pPS), derived from the genetic clustering of GWAS-identified T2D variants, have also been developed to capture biological processes underlying T2D genetic risk.^10–12^ PGS can also improve our understanding of disease pathophysiology. For example, Ritchie et al. tested cardiometabolic PGS for their association with protein expression levels, uncovering molecular mechanisms underlying polygenic risk.^13^ A more recent study by Steffen et al. incorporated T2D heterogeneity in their framework by testing the association between T2D pPS and protein levels.^14^ Leveraging PGS to identify the proteomic signature of disease can highlight key pathways and potentially find novel targets that may be missed by traditional disease-gene mapping approaches.

Here, we mapped genetic risk for T2D and its comorbidities to proteins, mechanistic pathways, and clinical outcomes. We constructed T2D PGS (a genome-wide PGS and a PGS restricted to GWAS-significant variants), five T2D pPS^10^ (beta cell, lipodystrophy, liver lipid, obesity, proinsulin), a BMI PGS, and PGS for common T2D cardiometabolic comorbidities (chronic kidney disease - CKD, coronary artery disease - CAD, non-alcoholic fatty liver disease - NAFLD). Then, we tested these PGS for associations with 2,922 plasma proteins in 54,306 participants from the UK Biobank Pharma Proteomics Project (UKB-PPP).^15,16^ We used pathway enrichment to identify key biological circuits and performed Mendelian randomization (MR) and mediation analyses to infer the PGS-associated proteins that might play a causal role. Finally, we tested whether the PGS-associated proteins were associated with clinical trial outcomes in two randomized controlled trials (RCT) of cardiovascular outcomes in T2D patients; Exenatide Study of Cardiovascular Event Lowering (EXSCEL)^17^ and Dapagliflozin Effect on Cardiovascular Events (DECLARE)-TIMI 58^18^ (2,823 and 915 participants with genomic and proteomic data, respectively). Together, our analyses identify the proteomic signatures of T2D-related polygenic scores which highlight specific protein biomarkers, mechanistic pathways, and putative therapeutic targets.

## Methods

See **Supplementary Figure 1** for a visual representation of the methods.

### Cohort description

The UK Biobank (UKB) is a deeply phenotyped population-based cohort comprised of approximately 500,000 subjects with array genotyping, exome sequencing, and whole genome sequencing data and linkage to electronic health care record data with over 14 years of follow-up time.^19–21^ The UK Biobank Pharma Proteomics Project (UKB-PPP) is a private-public partnership that has assayed 2,923 unique proteins (2,922 after excluding one protein without sufficient measurements) in a subset of 54,306 UKB participants using the Olink Explore proteomics platform.^15,16^ See **Supplementary Figure 2** for proteomic intersection between the three cohorts.

EXSCEL examined the cardiovascular effects of once-weekly exenatide, a glucagon-like peptide-1 (GLP-1) agonist, in T2D patients with a median follow-up time of 3.2 years.^17^ In a subset of trial participants (N=2,823), both genotyping and SomaScan proteomics data were generated (see **Supplementary Table 1**). DECLARE-TIMI 58 was a phase 3 RCT that examined the cardiovascular effect of dapagliflozin, an inhibitor of sodium-glucose co-transporter-2 (SGLT2), in patients with T2D with multiple risk factors for or established atherosclerotic cardiovascular disease with a median follow-up time of 4.2 years.^18^ Similar to EXSCEL, for a subset of participants both genotyping and Olink proteomics data were available (N=915).

See Extended Methods in **Supplementary Information** for a description of genotyping and proteomics quality control.

### UK Biobank phenotype definitions

For the UKB, we used ICD10 codes and clinically meaningful “Union” phenotypes constructed by merging relevant ICD10 codes (release from Feb 2022), as described previously.^22^ For identifying prevalent cardiometabolic conditions that needed to be excluded from analyses, we used the following ICD10 codes: E10-E14 (any diabetes diagnosis), K76.0 (NAFLD), N18 (CKD), and I20-I25 (ischaemic heart disease). We also excluded UKB participants with elevated (> 40 U/L) alanine aminotransferase (ALT) and aspartate aminotransferase (AST) at baseline^23^, defined in supplementary tables as NAFLD_AST_ALT. Incident cases were defined using ICD10 codes (E11 for T2D, K76.0 for NAFLD, N18 for CKD, I25 for CAD) when the earliest date of diagnosis (determined by fields 41270, 40001, 40002) occurred after baseline (when a sample was donated for proteomics). Note that relatively few cases were diagnosed within 30 days of sample collection (**Supplementary Table 2**).

For the generation of summary statistics for Mendelian randomization, we used the NAFLD_AST_ALT definition of NAFLD to maximize sample size, E11 for T2D, N18 for CKD, the union term of 120-I25 for ischaemic heart disease, and body mass index measured at baseline. For traits in diabetic UKB participants, we used obesity, defined as having a baseline BMI >= 30 kg/m^2^, and the ICD10 codes in **Supplementary Table 2,**which contains a complete description of UKB phenotype definitions and cohort sample size.

### Randomized controlled trials (RCTs) phenotype definitions

We used four outcomes that were available in both trials, i.e., time to insulin initiation, time to the composite cardiovascular outcome (major cardiovascular events or MACE, comprising of cardiovascular death, non-fatal myocardial infarction, or non-fatal ischemic stroke), time to hospitalization for heart failure, time to the renal outcome (for EXSCEL, two consecutive measurements of eGFR < 30 ml/min/1.73m^2^ and for DECLARE-TIMI 58, a composite renal outcome comprising of a sustained decrease of 40% or more in eGFR to < 60 ml/min/1.73m^2^, new end-stage renal disease, or death from renal causes). Note that competing risk of death was addressed through censoring and the use of cox regression.^24^ DECLARE-TIMI 58 cardiovascular endpoints were adjudicated by independent adjudication committees. For insulin initiation, we excluded participants already on insulin therapy. See **supplementary table 3** for a description of phenotypes.

### Polygenic score estimation

To avoid overfitting when estimating the PGS in the UKB, we retrieved genome-wide association study (GWAS) summary statistics from external studies that did not contain UKB participants (**Supplementary Table 4**), maximizing both sample size and diversity. We trained genome-wide T2D, CAD, BMI, and CKD PGS using PRS-CS^25^ (when only one GWAS per trait was available) or PRS-CSx^26^ with these GWAS summary statistics. We ran PRS-CSx and PRS-CS with phi set to ‘auto’. For NAFLD, we used a 77-variant score comprising of only GWAS-significant variants from a GWAS of unexplained chronic ALT elevation^27^ as a complete set of publicly-available well-powered NAFLD GWAS summary statistics were not readily available. For the partitioned T2D polygenic scores (pPS), we obtained the variants and cluster weights generated by Udler et al. corresponding to five distinct genetic clusters, i.e., beta cell, lipodystrophy, liver lipids, obesity, and proinsulin.^10^ To enable comparisons between genome-wide and GWAS-significant PGS, we also generated a set of GWAS-significant T2D summary statistics using the clump procedure implemented in PLINK v1.9 (p-value < 5×10^-8^, R^2^ < 0.1, 250 kilobase window around each index variant). All PGS for UKB, UKB-PPP, EXSCEL, and DECLARE-TIMI 58 were estimated using post-QC imputed data and PLINK v2.00a4LM.^28^ See **Supplementary Tables 5-6**, **Supplementary Figures 3-8**, and **Supplementary Information** for a complete description and PGS validation.

### PGS and protein associations in UK Biobank

For testing PGS for association with protein expression levels, we modelled our analysis on the same internal replication structure as used in the UKB-PPP consortium pQTL GWAS. First, we restricted the analysis to unrelated participants (resolved to the 2^nd^ degree) without a baseline diagnosis of diabetes or a major cardiometabolic condition at data collection (UKB’s baseline timepoint; N= 42,452) to reduce confounding due to reverse causality as previously suggested by Ritchie et al.^13^ Then, we stratified the cohort into the consortium-identified discovery subset consisting of European-ancestry participants (N= 28,105) and the replication subset of remaining pan-ancestry participants (N= 14,347). We tested each PGS for association with protein expression levels in the discovery subset using linear regression in R, using the same covariates (age, age^2^, sex, age*sex, age^2^*sex, batch, UKB centre, array, time to analysis, genetic PCs 1-20) as the UKB-PPP consortium pQTL GWAS^15^. Significant PGS-protein associations after a Bonferroni correction accounting for 10 scores and 2,922 proteins (p-value < 1.7×10^-6^) were then moved forward to be tested in the replication subset. To account for the difference in sample sizes in the two subsets, we applied an FDR correction to our replication analysis separately to each PGS.^13^ Associations were considered internally replicated with an FDR-adjusted p-value < 0.05.

To assess whether a PGS-protein association was driven by a single locus, we repeated the PGS-protein association analyses after adjusting for independent *cis* and *trans* pQTLs obtained from the UKB-PPP consortium’s pQTL GWAS. Similarly, we repeated PGS-protein association analyses after adjusting for BMI to describe the influence of BMI on the PGS associations. Finally, to explore the impact of genetic ancestry on the transferability of PGS-protein associations, we tested PGS-protein associations after stratifying the UKB-PPP cohort by predicted ancestry. We then compared the effect sizes (beta coefficients) of the PGS on circulating protein levels between ancestry groups using Pearson’s r and the slope of the regression line fitted to the beta coefficients.

### Mediation analyses in UKB

For mediation analyses, we utilised incident cases (prevalent cases were excluded) and the medflex R package^13,29^ (version 0.6-10) to perform mediation analysis with natural effects models. In this framework, we set the PGS as the exposure and the protein as the mediator. We considered a mediation model to be significant if the mediation (indirect) p-value and the total p-value were both significant after a Bonferroni correction (p-value < 1×10^-6^); the direct effect p-value was allowed to be not significant as it is possible that the PGS’s effect is primarily mediated through the tested protein. As a sensitivity analysis, we performed mediation in both all participants and only European ancestry; we filtered out proteins that were not significant in both scenarios.

### 2-sample Mendelian randomization (MR)

We performed two-sample Mendelian randomization (MR) analysis to conduct a proteome-wide scan for proteins suspected to play a potential causal role in the development of T2D, a T2D comorbidity, or a T2D complication. To identify weak instruments, we calculated the F-statistic^30^ for each instrument (F-statistic = {3^2^⁄Se^2^). We performed MR on all proteins with 3 or more *cis* pQTLs using the MendelianRandomization^31^ R package (version 0.7.0) and applying the simple and weighted median, IVW, and MR-Egger methods, finding the consensus effect by taking the median of the estimate, standard error, 95% confidence interval, and p-value across all methods. We addressed violations of the pleiotropy assumption by excluding results with an MR-Egger intercept p-value < 0.05.^13,32^ For traits available in the entire UKB cohort, we applied a Bonferroni correction, while for traits in type 2 diabetics (ICD10 code E11) we applied an FDR correction due to the smaller sample sizes. See **Supplementary Information** for a description of MR sensitivity analyses including statistical colocalization and the GWAS summary statistics used in MR.

### Reverse Mendelian randomization

We obtained MR instruments for reverse MR from the T2D, CKD, NAFLD, BMI, and ischemic heart disease GWAS data that we utilized in our forward MR analysis. We performed LD pruning using PLINK v1.90b6.18’s clumping procedure with a 500 KB window size, a minimum p-value threshold of 5×10^-8^, and R^2^ < 0.001 using UKB-PPP as the LD reference. We matched each instrument with variants from the pQTL GWAS data. We then performed reverse MR using the MendelianRandomization R package in the same manner as described above.

### Association and mediation of proteins with trial outcomes in EXSCEL and DECLARE-TIMI 58

We tested proteins for their association with the time to EXSCEL outcomes with the survival package in R 3.6.1 (https://github.com/therneau/survival) and replicated significant associations in DECLARE-TIMI 58. We used Cox proportional hazards regression and adjusted for age, sex, age*sex, genetic PCs 1-10, trial arm, and (+/-) BMI. As in the case of the UKB, analyses were repeated with a BMI adjustment to identify associations likely mediated by BMI/obesity. For both EXSCEL and DECLARE-TIMI 58, two timepoints were available for proteomics: baseline plus 12 months for EXSCEL, and baseline plus 6 months for DECLARE. In both studies, we performed the proportional hazards regression analysis three times, using the baseline measurements, the repeat measurement, and the difference (delta) between the two measurements, respectively, as exposures. For EXSCEL, each SomaLogic aptamer was tested separately and reported. In the case of statistically significant associations in the clinical trial time-to-event analyses, (Bonferroni threshold of p-value < 1.3×10^-5^), we assessed the proteins using more comprehensive models that included clinical risk factors to evaluate their suitability as a biomarker. See **Supplementary Information** for a description of the clinical risk factors.

In the scenarios where a PGS is associated with both an outcome and a protein (see **Supplementary Information**), and the protein is in turn also associated with the same outcome, we performed mediation analysis using the medflex R package (version 0.6-10) in the same manner as above, with the indirect p-value adjusted using the FDR approach.

### Pathway Enrichment

For all sets of proteins identified by the PGS-protein analyses, we used the gProfiler^33^ tool (https://biit.cs.ut.ee/gprofiler/gost) to test if these sets of encoded genes were enriched for KEGG, WikiPathways, or REACTOME pathways. Note that we restricted the statistical domain to the genes whose protein products are captured by the Olink panels.

## Results

### Associations of PGS with protein expression levels

Identifying circulating proteins that associate with PGS for T2D and its cardiometabolic comorbidities can shed light on proteins important in the development of T2D and related comorbidities. Also, this analysis can help define mechanisms through which a PGS exerts its effect and describe the overall proteomic signature of a PGS. The genome-wide T2D PGS (PGS_T2D_gw_) was associated with 713 proteins in the UKB discovery set (**Table 1**); of these, 686 replicated in the UKB replication set (FDR < 5%). The proteins that were among the top 1% in terms of variance (R^2^) explained by the PGS_T2D_gw_ include PON3, CKB, APOF, and IGFBP2 (**Figure 1A**). Out of the 713 proteins significantly associated with the PGS_T2D_gw_ in the discovery set, 341 remained significant after adjustment for BMI (**Figure 1B**), demonstrating the close interplay between T2D genetic risk, BMI, and circulating protein levels.

**Figure 1:**
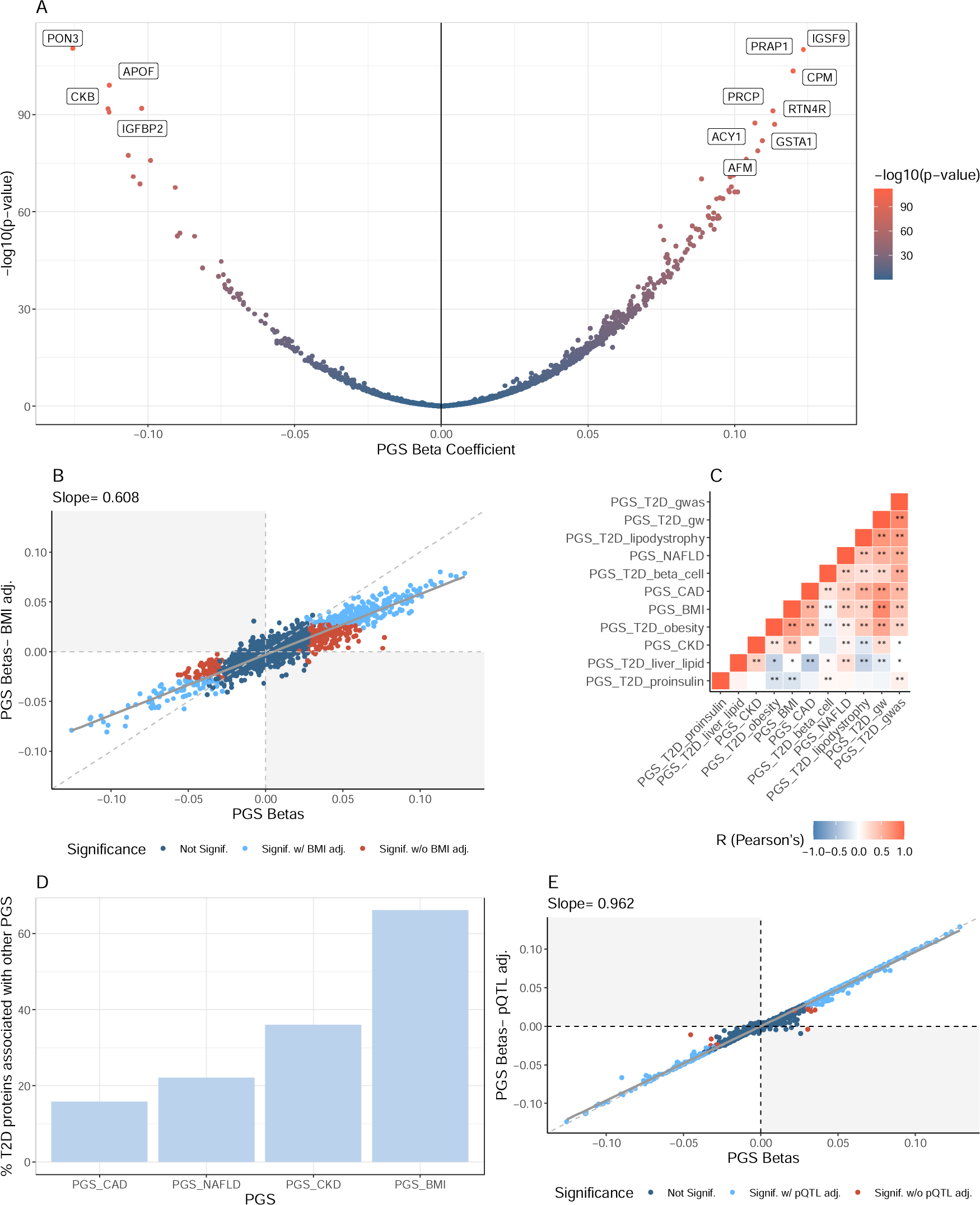
PGS associations with circulating proteins. A: Volcano plot of PGS_T2D_gw_-protein - log_10_ p-values and beta coefficients with the colour indicating −log10 p-values. Labelled proteins are among the top 1% in terms of variance (R^2^) explained by the PGS_T2D_gw_. B: Beta-beta plot of PGS_T2D_gw_ effect sizes on circulating proteins with (y-axis) and without (x-axis) BMI adjustment. The diagonal is dashed grey while regression line is solid grey. Each point represents a protein; light blue points indicate replicated proteins that remained significant with the adjustment, red points indicate replicated proteins that were no longer significant after the adjustment, and dark blue points indicate proteins that did not significantly replicate prior to adjusting for BMI or pQTLs. C: Correlations of PGS effect sizes from the regression on circulating protein levels. Red indicates pairs of PGS with positively correlated effect sizes; blue indicates negatively correlated effect sizes. D. Bar plot indicating the overlap between proteins significantly associated with the T2D PGS and the other cardiometabolic PGS. The x-axis is the PGS and the y-axis is the percentage of T2D PGS-associated proteins that are also associated with another PGS (e.g., over 60% of proteins were also associated with the BMI PGS). E: Beta-beta plot of PGS_T2D_gw_ effect sizes on circulating proteins with (y-axis) and without (x-axis) pQTL adjustment, with the same definitions as panel B, albeit for a pQTL adjustment.

**Table 1:** Summary of PGS associations. PGS Category: descriptive information for the PGS. PGS: Name of tested polygenic scores. Top Protein Association: protein with the lowest p-value in the combined UKB-PPP cohort (discovery + replication) for each PGS. N significant proteins: number of significant proteins after replication. The brackets specify the total number of proteins that were significant in discovery the subset. N significant after pqtl/BMI adjustment: number of replicated proteins remaining significant after pQTL and BMI adjustments. The brackets specify the total number of proteins that were significant after adjustment in the discovery subset. For the BMI PGS and the obesity pPS, we did not also adjust for BMI. % Unique Assoc.: percentage of significant PGS-associated proteins that were only associated with that PGS.

| PGS Category | PGS | Top protein assoc. | N significant Proteins | N significant after pqtI/BMI adjustment | % Unique Assoc. |
| --- | --- | --- | --- | --- | --- |
| **T2D (Genome-wide)** | PGS<sub>T2D_gw</sub> | PON3 (-) | 686 (713) | 325 (331) | 15 |
| **T2D (GWAS-significant only)** | PGS<sub>T2D_gwas</sub> | PAM (-) | 67 (84) | 50 (58) | 1 |
| **Partitioned T2D polygenic scores** | PGS<sub>T2D_beta_cell</sub> | ABO (+) | 17 (26) | 1 (2) | 24 |
|  | PGS<sub>T2D_lipodystrophy</sub> | LPL (-) | 102 (131) | 95 (110) | 0 |
|  | PGS<sub>T2D_liver_lipid</sub> | MAMDC4 (-) | 268 (322) | 7 (7) | 18 |
|  | PGS<sub>T2D_obesity</sub> | LEP (+) | 6 (12) | 4 (9) | 0 |
|  | PGS<sub>T2D_proinsulin</sub> | FOLR3 (-) | 0 (0) | 0 (0) | - |
| **Other cardiometabolic scores** | PGS<sub>CAD</sub> | GRN (+) | 136 (175) | 74 (85) | 5 |
|  | PGS<sub>CKD</sub> | BTN2A1 (+) | 559 (582) | 499 (511) | 27 |
|  | PGS<sub>BMI</sub> | LEP (+) | 645 (693) | 625 (667) | 9 |
|  | PGS<sub>NAFLD</sub> | ASS1 (+) | 222 (267) | 164 (183) | 16 |

The partitioned T2D scores and the PGS_T2D_gwas_ (i.e., the PGS derived from GWAS-significant variants) were associated with fewer proteins compared to the genome-wide T2D score (PGS_T2D_gw_) (**Supplementary Figures 9-13, Supplementary Table 7**). Despite comprising of fewer variants than PGS_T2D_gw_, the PGS_T2D_beta_cell_, PGS_T2D_liver_lipid_, and PGS_T2D_gwas_ were significantly associated with proteins that were not associated with any other PGS (see **Table 1**). When comparing the effect sizes of the different T2D PGS for the circulating proteins, the PGS_T2D_gw_ beta coefficients were negatively correlated with the PGS_T2D_liver_lipid_ (r = −0.15, p = 6.9×10^-12^) and not correlated with PGS_T2D_proinsulin_ (p > 0.05, **Figure 1C)**. Overall, this suggests that the partitioned T2D scores capture protein associations representing perturbations in specific biological pathways that may be obscured when variant effects are aggregated in a genome-wide score (PGS_T2D_gw_).

We also tested a selection of cardiometabolic PGS representing common T2D comorbidities (CAD, CKD, BMI, and NAFLD) for their association with protein expression levels (Panel A from **Supplementary Figures 10-17**) to determine their proteomic signatures and compare them with that of the T2D scores. The proteins associated with the PGS_T2D_gw_ were frequently associated with one or more cardiometabolic PGS (**see Figure 1D**). Overall, 66% were also significantly associated with the PGS_BMI_, 36% were associated with the PGS_CKD_, 22% were associated with the PGS_NAFLD_, and 16% were also associated with the PGS_CAD_. The effect sizes of these scores on the circulating protein levels were all positively correlated with that of the PGS_T2D_gw_ (p-value < 0.05, **Figure 1C**), supporting the epidemiological observation that cardiometabolic diseases are highly interconnected.^34,35^ However, for select proteins, the directions of effect of the T2D_T2D_gw_ on circulating levels were in fact opposite when compared to the effects of another PGS (e.g., the effect of PGS_T2D_gw_ and PGS_BMI_ on TIMP4 levels, see **Supplementary Information**, **supplementary figure 18, and Supplementary Table 8**), likely reflecting more complicated relationships such as compensatory responses.^36–38^ In the case of the partitioned scores, the correlation patterns seem to point to proteomic mechanisms that explain their differential associations with T2D comorbidities. Notably, the protein effect sizes of the PGS_T2D_liver_lipid_ were negatively correlated with those of the PGS_CAD_ (r = −0.39, p=1.7×10^-125^), but were positively correlated with the PGS_CKD_ (r= 0.31, p = 9.7×10^-87^), mirroring the decreased risk of CAD and increased risk of CKD that have been described for this score in previous studies.^10,11^

The correlations of PGS effect sizes on circulating proteins between genetically predicted European and predicted non-European ancestries were less than 1, reflecting the fact that PGS translatability across ancestries is a potential concern^39^ (see **Supplementary Figures 19-23, Supplementary Table 9**). However, this correlation was substantially improved when only comparing statistically significant PGS-protein associations. See **Supplementary Information** for further discussion of cross-ancestry PGS-protein associations. We also tested all PGS for their association with circulating proteins in patients with prevalent T2D from the UKB, EXSCEL, and DECLARE-TIMI 58, results of which are available in the Supplementary Information, **Supplementary Figures 24-25**, and **Supplementary Tables 10-14)**.

### Polygenicity of PGS-protein associations and widespread effects of GCKR

An important consideration regarding PGS-protein associations is whether they are driven by a single pQTL tagged by the PGS or represent the cumulative, polygenic effect of the PGS. To this end, we included pQTLs in our PGS-protein regression models. After adjusting for both *cis* and *trans* pQTLs (see Methods), the vast majority of protein-PGS_T2D_ associations remained significant (648 out of 686; see **Figure 1E**, **Table 1, Supplementary Table 7**), demonstrating that these associations were indeed polygenic in nature. This was largely true for all evaluated PGS. However, for the PGS_T2D_liver_lipid_ and the PGS_T2D_beta_cell_, most protein associations were not significant after pQTL adjustment (**Supplementary Figure 9C and 11C**). In the case of the PGS_T2D_liver_lipid_, the index variant at the *GCKR* locus (rs1260326) explained the bulk of its association signature (256 out of 268 proteins), highlighting the pleiotropic effect of this variant on circulating proteins. In contrast, most PGS_T2D_beta_cell_ associations were explained by 9 different pQTLs, with two from the ABO locus, one from the ANPEP locus, one from the FGFBP3 locus, and the remainder from intergenic regions.

### Causal inference using Mendelian randomization

To determine if a PGS-protein association is due to forward causality (i.e., the PGS perturbs a protein’s expression level which leads to disease), we utilized two-sample Mendelian randomization (MR) with *cis* pQTLs as instruments. We evaluated five traits cohort-wide (ischemic heart disease, CKD, T2D, NAFLD, and BMI) and comorbidities/complications related to T2D, i.e., retinopathy, hypertension. We identified 24 proteins with plausible evidence for causality for a trait (adjusted p-value < 0.05 and/or colocalization evidence; **Figure 2**), of which 21 were also significantly associated with at least one PGS (**Supplementary Tables 15-16**) and 13 with one or more T2D PGS in our analysis above. Notably, ERBB4, significantly associated with the PGS_BMI_ and the PGS_T2D_liver_lipid_, is thought to be a mediator of the development of metabolic disorders (e.g., T2D and NAFLD) in individuals with obesity^40–43^

**Figure 2:**
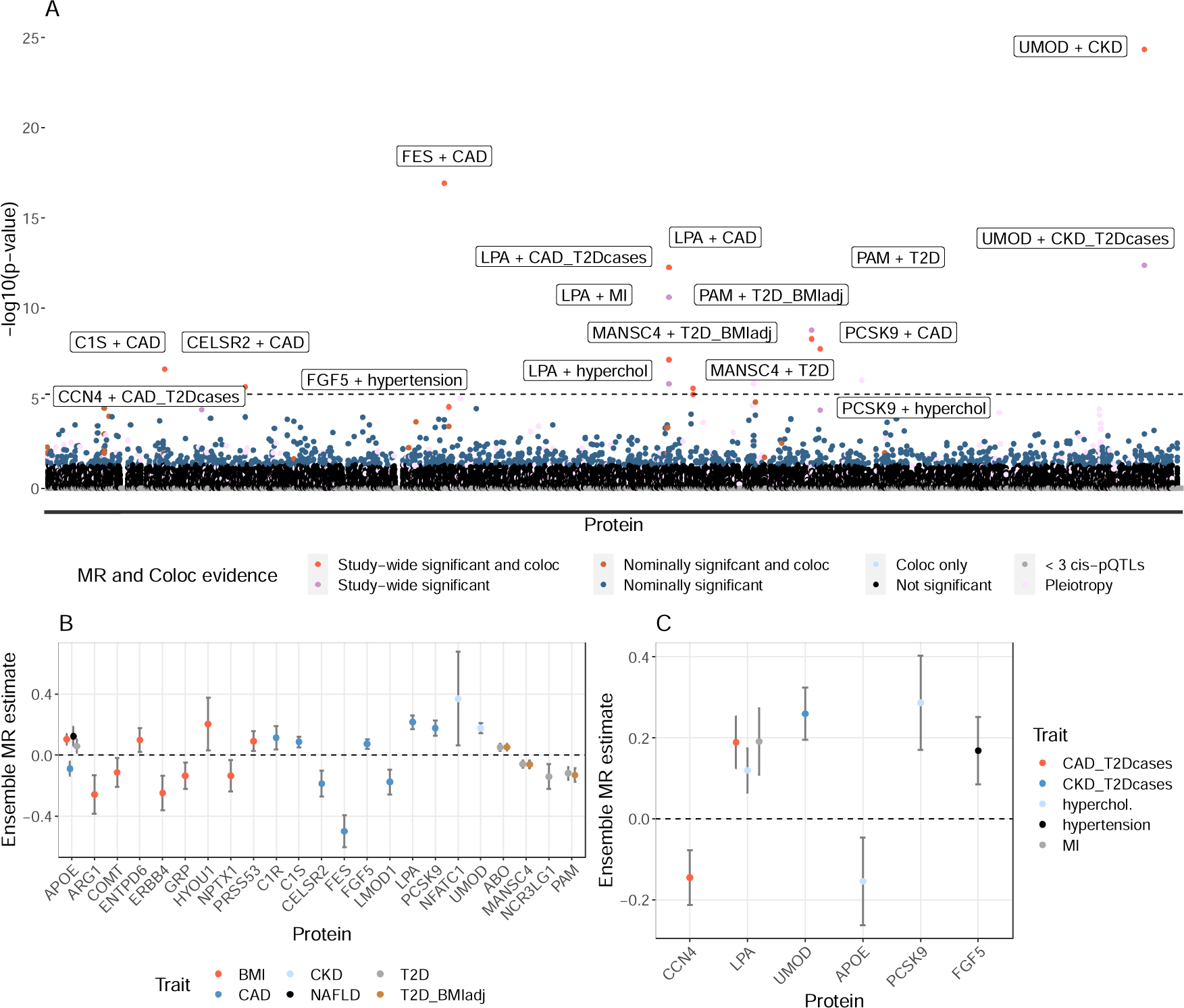
Two-sample Mendelian Randomization analysis in UKB. A: Manhattan plot of MR results. Labels refer to the protein and trait combination (e.g., UMOD + CKD). Traits appended with “_T2Dcases” refer to comorbidities analysed in T2D cases only. Note that CKD includes the ICD10 codes N18.1-N18.5. CAD referrers to ischaemic heart disease ICD10 codes (I20-I25). See Supplementary Table 2 for phenotype definitions. P-values were calculated from the median of all MR methods. The dashed line indicates Bonferroni significance. B: Forrest plot of MR estimates for phenotypes assessed in all UKB participants and proteins with strong causal evidence. C. Forrest plot of MR estimates for proteins with strong causal evidence for phenotypes assessed only UKB diabetic patients.

For T2D, we performed three sets of analyses: MR using summary statistics unadjusted for BMI, MR using BMI-adjusted summary statistics, and MR using summary statistics expanded to include all diabetes-related ICD10 codes (E10-E14). PAM, ABO, and MANSC4 had strong MR evidence in all scenarios, though PAM pQTLs only colocalized with T2D risk variants when we included all diabetic codes. The effect of NCR3LG1 on T2D risk was likely mediated by BMI as it was no longer significant after BMI adjustment. While we did not detect pleiotropy using MR-Egger at the ABO locus, this region is known for its extensive pleiotropy.^44^ Notably, the circulating levels of ABO were significantly associated with the PGS_T2D_beta_cell_, but not the overall PGS_T2D_gw_, again highlighting the utility of the partitioned T2D scores. Colocalization evidence was generally lacking for traits evaluated in T2D patients, likely due to limited statistical power (see **Supplementary Table 16**).

While 1,414 proteins had 3 more independent *cis* pQTLs, an additional 625 proteins without at least 3 *cis* pQTLs nevertheless had 3 or more *trans* pQTLs (or a combination of *cis* and *trans*). We considered the MR results for the proteins meeting this criterion provided there was not any statistical evidence of pleiotropy (Egger intercept p-value > 0.05; **Supplementary Table 15**). Notably, FURIN had *cis* colocalization evidence and a significant MR estimate when using 2 *cis-*pQTLs and 5 *trans*-pQTLs. FURIN has previously been implicated in cardiovascular disease^45–47^. In addition, we found significant MR estimates for APOBR with BMI, GREPEL1 and SDHB with NAFLD, and ASS1 with NAFLD in patients with T2D. Since MR with *trans* pQTLs needs to be done with caution, we generated dose-response curves for these proteins to ensure that MR estimates were not driven by a single *trans* pQTL (see **Supplementary Figure 26**).

### The mediation of PGS effects by circulating proteins in the UKB

An orthogonal approach to MR for inferring the causal pathway is mediation, which tests whether a protein mediates the effect of a PGS on incident disease risk. Mediation can be used to support MR findings, and it can also provide information on directionality for proteins that lack the requisite number of instruments for MR. Among the UKB participants with proteomics data, 2,081 were diagnosed with T2D during 14 years of follow-up time. In our mediation analysis, 536 of the 686 PGS_T2D_gw_-associated proteins significantly mediated the effect of the PGS on incident T2D risk. After adjusting for BMI, this is reduced to 236 proteins (**Supplementary Figure 27A, Supplementary Table 18**). We also performed mediation using the partitioned scores (**Supplementary Figure 27B**): 81 proteins mediated the PGS_T2D_lipodystrophy_ score (82 after BMI adjustment) and 5 mediated PGS_T2D_beta_cell_ score (3 after BMI adjustment). Of particular note, FAM3D, the top mediating protein for PGS_T2D_beta_cell_ is thought to be involved in glucagon and glucose regulation.^48–50^ We also performed mediation of the PGS_CKD_ with incident CKD, the PGS_NAFLD_ with incident NAFLD, and the PGS_CAD_ with incident CAD, finding 513 (472 after BMI adjustment), 190 (171 after BMI adjustment), and 84 (31 after BMI adjustment) mediating proteins, respectively (**Supplementary Figure 27A**). In our *cis*-pQTL MR results for the four traits evaluated via mediation, 3 out of 8 proteins were also significant in mediation when using a Bonferroni adjustment, which increased to 14 out of 16 when employing a FDR correction. Since the PGS used in mediation are not restrained to *cis* variants, we also compared it to our MR results when using both *cis* and *trans* pQTLs. In this case, 8 out of 13 Bonferroni-adjusted proteins and 22 out of 28 FDR-adjusted proteins overlapped.

### Reverse Mendelian Randomization with cardiometabolic traits

Next, we employed reverse MR to identify instances of reverse causation, where the cardiometabolic trait (T2D, ischemic heart disease, NAFLD, CKD, BMI) alters the protein expression levels (**Methods**). We found that circulating levels of 40 proteins were influenced by T2D (37 in European-ancestry analysis), including GDF15 (**Supplementary Table 17**) and several proteins strongly associated with the PGS_T2D_gw_ (e.g., APOF, PON3, PRCP). In the case of GDF15, elevated serum levels have been reported in T2D and it is known to play a role in regulating food intake and metabolism.^51,52^ However, proteins identified in the forward MR analysis for T2D (ABO, PAM, and MANSC4) were not implicated in our reverse MR. It is worth noting that proteins identified via reverse MR could be causal for other comorbidities or could still influence T2D risk via feedback mechanisms.

For the other cardiometabolic disorders, we found 4 proteins influenced by CAD risk (MMP12, CNTN4, PAMR1, PCOLICE), 19 for CKD, 79 for NAFLD, and 513 for BMI (**Supplementary Table 17**). The high number of proteins influenced by BMI genetic risk indicate that the levels of many circulating proteins are impacted by adiposity levels. None of the proteins we identified in our forward MR were associated with the trait in reverse MR, including the proteins that showed an association with BMI. However, when we overlaid our mediation results with the reverse MR results, a proportion of mediating proteins had evidence for reverse causality, ranging from 3.7% of proteins for CKD and PGS_CKD_ to 39% for NAFLD and the PGS_NAFLD_. Such proteins could either indicate confounding or the presence of feedback mechanisms and require further investigation.

### Time-to-event analyses of PGS-associated proteins in EXSCEL and DECLARE

To further explore the clinical relevance of PGS-associated proteins, we tested them for their association with clinical trial endpoints in two trials with available proteomics data (assayed at baseline): EXSCEL and DECLARE-TIMI 58. Both were cardiovascular outcome trials with similar indications: patients with T2D and either established cardiovascular disease or multiple risk factors (Methods). We first tested a model including only basic covariates (demographics and treatment arm) to find proteins associated with the outcome, followed by models that included additional risk factors to identify potential protein biomarkers (**Supplementary Figure 28, Supplementary Tables 19-20**).

In EXSCEL, the baseline levels of 281 PGS-associated proteins were significantly associated with the renal outcome, 157 proteins were associated with the MACE outcome, 55 with the HHF outcome, and 5 with the insulin initiation outcome. After adjusting for relevant clinical risk factors, these lists were reduced to 79 (renal), 127 (MACE), 12 (HHF), and 2 for insulin initiation (C2 and PLXNB2), respectively. For the MACE outcome, 35 were also independent of NT-proBNP, while for HHF, 5 were independent of NT-proBNP. In DECLARE, 81 proteins of the proteins associated with any EXSCEL outcome (without clinical risk factors) were available for replication, of which 28 significantly replicated using the corresponding outcome in DECLARE. When adjusting for clinical risk factors, TFF3 significantly replicated for the renal outcome (**Figure 6**), 6 proteins for the MACE endpoint (**Supplementary Figure 29**), and 3 for the HHF endpoint (**Supplementary Figure 30**). Notably, EFEMP1 remained significantly associated with the HHF endpoint after adjusting for NT-proBNP.

Both trials featured repeat measurements with proteins assayed at baseline and at a second timepoint (12 months for EXSCEL, 6 months for DECLARE). For both trials, hazard ratios for all four outcomes were highly correlated (r = 0.96 in EXSCEL and 0.89 in DECLARE, p-values < < 2×10^-16^), though hazard ratios were on average larger at the second time point (**Supplementary Figure 31**). In total, 42 proteins significantly replicated using either timepoint; of these proteins, 59% significantly replicated at both timepoints, 7% only replicated at baseline, and 33% replicated only at the second timepoint. Using the second timepoint for protein measurement increased the number of proteins associated with a trial endpoint, particularly for the renal outcome (**Figure 3C and D**) where EPHB4, TNFRSF1A, HAVCR1, and CD93, in addition to the already described TFF3, significantly replicated after adjusting for clinical risk factors in DECLARE.

**Figure 3:**
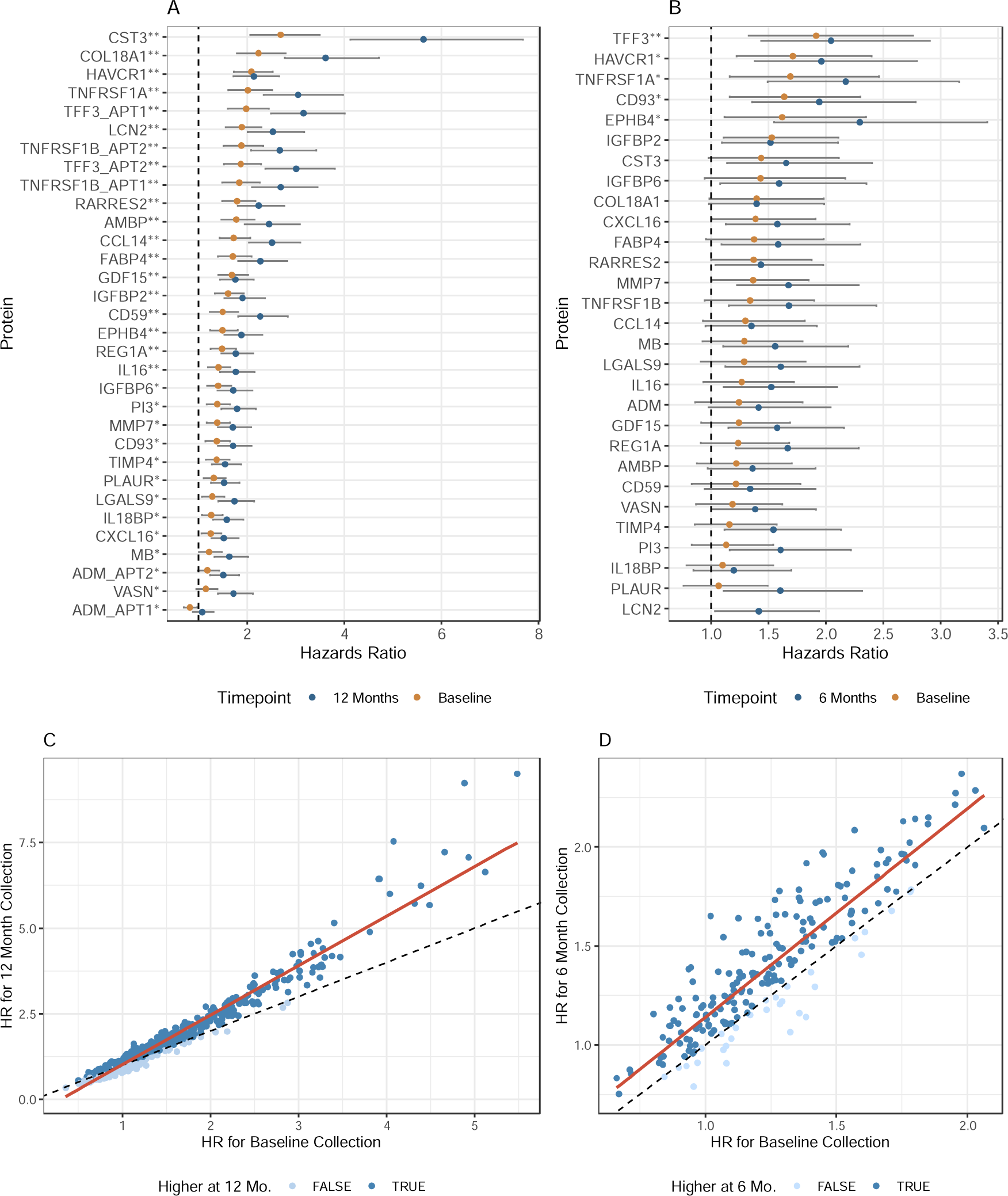
Renal outcome biomarkers in randomized controlled trials discovered in EXSCEL and replicated in DECLARE-TIMI 58. All proteins displayed here were significantly associated with the renal outcome in EXSCEL after adjusting for clinical factors, were available in DECLARE, and were nominally associated using the demographics-only model. A. Hazard ratios of the proteins associated with EXSCEL’s renal outcome after adjusting for clinical risk factors, with yellow indicating the proteins were measured at baseline and blue indicating the proteins were measured as 12 months. Proteins with a “**” were significant at both timepoints, while proteins with a “*” were significant at only one timepoint. B. Hazard ratios of the proteins associated with DECLARE’s composite renal outcome after adjusting for clinical risk factors, with yellow indicating the proteins were measured at baseline and blue indicating the proteins were measured as 6 months. Proteins with a “**” significantly replicated at both time points and proteins with a “*” significantly replicated at one timepoint. C. Scatterplot of hazards ratios (unadjusted for clinical risk factors) at baseline (x-axis) and 12 months (y-axis) for EXSCEL. The regression line is in red, while the dashed black line indicates where the hazard ratios would be in complete concordance. D. Scatterplot of hazards ratios (unadjusted for clinical risk factors) at baseline (x-axis) and 6 months (y-axis) for DECLARE. The regression line is in red, while the dashed black line indicates where the hazard ratios would be in complete concordance.

### PGS mediation in EXSCEL and DECLARE

Since MACE was significantly associated with the CAD PGS in EXSCEL (see **Supplementary Information**, **Supplementary Figures 32-33, Supplementary Tables 21-22** for PGS associations with clinical outcomes), we utilized a mediation framework to evaluate CAD PGS-associated proteins to determine how the PGS and the protein relate to each other in the causal pathway. We found evidence that the CAD PGS mediates its effect through the circulating protein levels of C9, LBP, ITIH4, APOM, and HS6ST2 (**Supplementary Figure 34 A, Supplementary Table 23**). In DECLARE, the BMI PGS was significantly associated with HHF and the CAD PGS was significantly associated with MACE. In our DECLARE mediation analysis, we did not identify any proteins that mediated the CAD PGS’s effect. However, for the BMI PGS and HHF, we found evidence for 63 proteins mediating its effect (13 after employing a Bonferroni correction instead of FDR; see **Supplementary Figure 34B, Supplementary Table 24**). When including BMI in the models, 17 proteins still appeared to mediate the BMI PGS’s effect, though at a nominal significance level (p < 0.05; **Supplementary Figure 34C, Supplementary Table 25**).

### Pathway Enrichment reveals shared mechanisms across cardiometabolic disorders

Enrichment analyses can identify critical pathways for the etiology of cardiometabolic pathways beyond individual protein associations. The PGS_T2D_gw_ was significantly enriched for 20 pathways after p-value adjustment (**Methods**). Notably, 15 of the pathways were also enriched for another PGS, 11 of which were enriched for three PGS (**Supplementary Figure 35**). Shared pathways are of a particular interest due to their potential involvement across cardiometabolic disorders.

The insulin-like growth factor binding proteins (IGFBPs) pathway was enriched in the PGS_T2D_gw_, PGS_BMI_, and the PGS_T2D_liver_lipid_ protein association sets. IGFBP2 was among the top protein associations with the PGS_T2D_, while IGFBP4 and IGFBP6 were strongly associated with the PGS_CKD_ (**Figure 4A**). This pathway also includes notable cardiovascular risk proteins, including PCSK9 and FGF23, which interact with IGF and IGFBPs via FAM20C.^53^ While causal effects for IGFBPs on the tested diseases were not supported in our MR analysis, IGFBP2 and IGFBP6 were implicated with T2D and CKD, respectively, in our mediation framework. In the RCTs, many IGF-related proteins were significantly associated with outcomes (**Figure 4B and D**). IGFBP2 was a robust biomarker of reduced kidney function in EXSCEL and nominally significant in DECLARE (**Figure 3A and B**). In contrast, lower levels of circulating IGFBP2 were associated with incident T2D risk (**Figure 4A**), corroborating previous observations regarding the apparent inverse relationship between incident diabetes and diabetic kidney disease.^54^ Higher levels of IGFBP2 also increased the probability of experiencing one of the events comprising the MACE outcome (**Figure 4C and E**).

**Figure 4:**
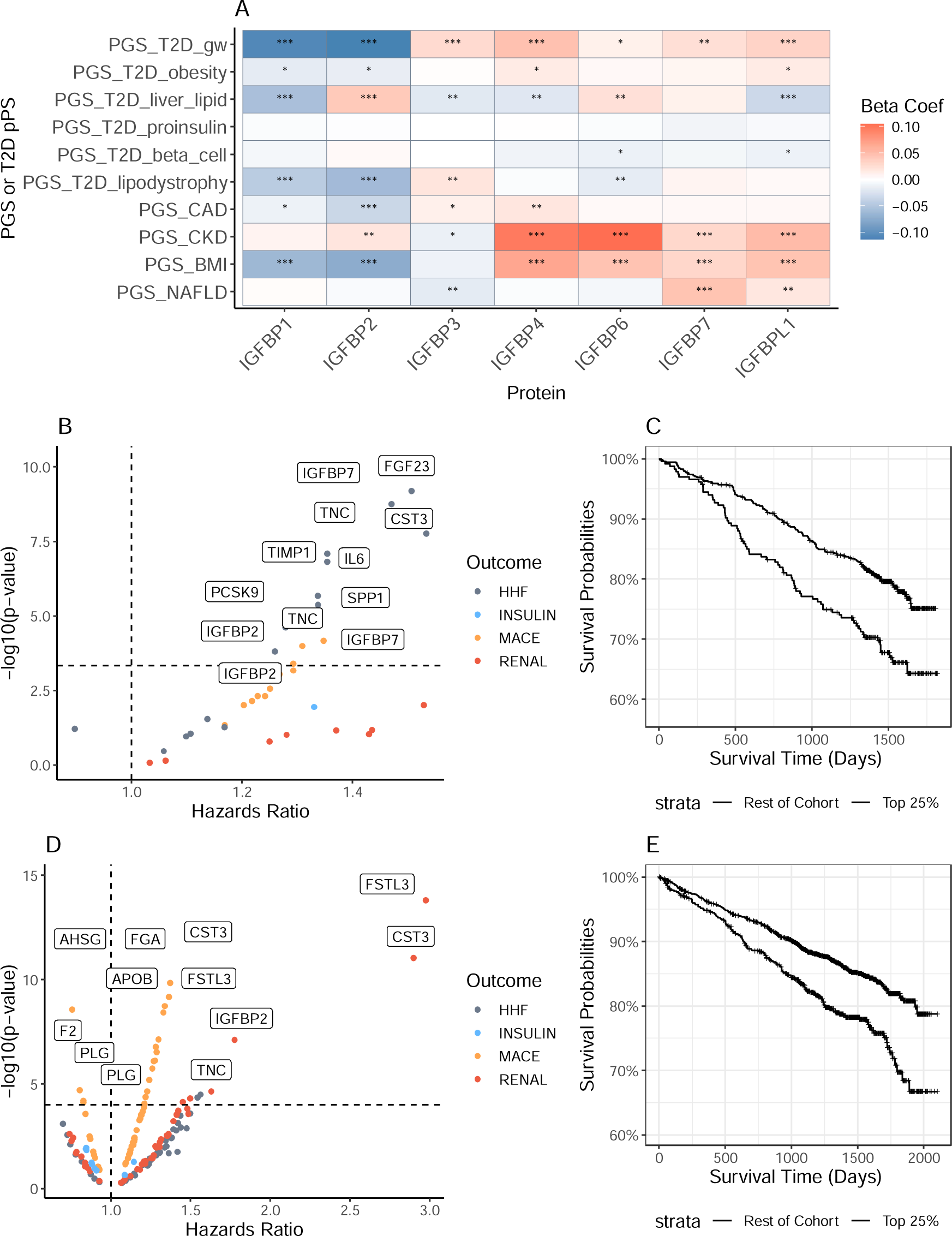
IGF regulation by IGFBPs pathway. A: PGS associations with IGF binding proteins. A single asterisk (*) indicates the association was nominally significant, while two (**) indicates significance using FDR and three (***) indicates significance using a Bonferroni correction. B: Associations of proteins in this pathway with clinical trial outcomes in DECLARE using cox regression. C. Kaplan Meier curve demonstrating the impact of IGFBP2 levels on the MACE endpoint in DECLARE. D: Associations of proteins in this pathway with clinical trial outcomes in EXSCEL using cox regression E. Kaplan Meier curve demonstrating the impact of IGFBP2 levels on the MACE endpoint in EXSCEL.

Pathways involving the complement system were enriched in the sets of proteins associated with the PGS_T2D_gw_ and PGS_CAD_ (**Supplementary Figure 35, Supplementary Figure 36**). Proteins in the complement and coagulation cascades pathway were identified as causal for cardiovascular disease in MR (C1R and C1S) and mediation (14 proteins mediated the effect of PGS_T2D_gw_ and the protein F9 mediated the effect of the PGS_CAD_, **Supplementary Table 18**). In the RCTs, C2 was associated with time to insulin initiation in EXSCEL. In addition, C2 and CD59 from the complement cascade and PLAUR from the coagulation cascade were associated with the time to MACE and HHF in both EXSCEL and DECLARE. (**Supplementary Figure 36 D and E**).

## Discussion

The complex biological basis of T2D (e.g., numerous GWAS risk variants, gene targets, and pathways) is reflected in its diverse clinical consequences (e.g., differences in disease progression and comorbidity development). Identifying additional genes and pathways and mapping our existing knowledge to clinical outcomes remains a major challenge. Large-scale proteogenomic analyses present an opportunity to implicate specific, potentially causal, proteins with T2D and its comorbidities. We tested several different T2D polygenic scores (PGS), including five partitioned scores capturing pathophysiological pathways underlying T2D ^10^, and a number of PGSs for T2D co-morbidities (BMI, CKD, CAD, NAFLD), for their association with circulating proteins in healthy, unrelated individuals in the UK Biobank.^15^

Our findings yielded insights into the proteomic consequences of T2D polygenic risk. Notably, the partitioned T2D PGS seemed to capture unique aspects of T2D biology (189 proteins were associated with the partitioned scores but not the PGS_T2D_gw_), thus, serving as proof of concept for the development of such scores. For example, the PGS_T2D_beta_cell_ was strongly associated with ABO, which, in turn, was associated with T2D in our MR analysis, and was mediated by FAM3D, two causal relationships not detected by the overall PGS_T2D_gw_. Further, we found the effect sizes of PGS_T2D_liver_lipid_ and the PGS_T2D_gw_ were negatively correlated (**Figure 1**). It seems plausible that the *GCKR* locus (index variant: rs1260236), captured by PGS_T2D_liver_lipid_, drives a specific subtype of T2D with unique phenotypic consequences.^10,11^

Pathway enrichment of PGS-associated proteins can help guide target identification for therapeutics (**Supplementary Figure 33**). Pathways shared across multiple cardiometabolic disorders could be particularly attractive in this regard. We have highlighted the regulation of IGF by IGFBPs pathway and the complement and coagulation cascades pathway. Therapies targeting the complement system have been developed or proposed for a wide range of diseases, including inflammatory kidney disorders and cardiovascular disease.^55–58^ Other notable pathways enriched in the PGS associations with strong causal evidence include the plasma lipoprotein assembly, remodelling, and clearance pathway (PCSK9, LPA, APOE, and FURIN with cardiovascular disease; APOE with both BMI and NAFLD). The influence of PCSK9 and LPA on cardiovascular disease is well-known.^59–61^

We found several proteins with an inverse relationship between T2D and CKD risk. For example, low levels of IGFBP2 were strongly associated with T2D risk, while higher levels were associated with CKD risk. IGFBP2’s link with T2D has been previously described, but its functional role in obesity, insulin resistance, and ultimately diabetes remains incompletely understood.^62^ In total, the PGS_T2D_gw_ and the PGS_CKD_ had opposing directions of effect for 65 proteins, more than any other T2D-cardiometabolic disease combination we tested (**Supplementary Figure 18**). Overall, the directions of effect for the PGS_T2D_gw_ and the PGS_CKD_ were positively correlated, so investigating the complexities in the interface of T2D-CKD is an area of further exploration.

To explore the chain of causality underlying PGS-protein associations, we utilized a mediation framework and MR (see **Figure 2**, **Supplementary Figure 26**). We then leveraged two randomized controlled T2D trials (RCTs), EXSCEL and DECLARE-TIMI 58, to examine how predictive PGS-associated proteins were of clinical trial outcomes (**Supplementary Tables 19 and 20**). Several proteins implicated in this analysis, such as TFF3 and EFEMP1, could potentially explain mechanisms behind the development of diabetic comorbidities.^63–65^ We also described 6 proteins that mediated the CAD PGS’s effect on MACE in EXSCEL (C9, LBP, ITIH4, APOM, CES1, and HS6ST2), providing supporting evidence that they might serve as biomarkers for cardiovascular disease in diabetic patients.^66–68^

We developed an interactive portal that permits users to interrogate the results of our analyses (https://public.cgr.astrazeneca.com/t2d-pgs/v1/; **Supplementary Figure 37**). Our portal enables querying by protein, pathway, or PGS. The underlying data is then available for download. Our portal allows researchers to validate their findings and gather evidence for target identification. Furthermore, the portal puts decision-making power into the hands of the user. For example, we elected to employ strict p-value thresholds and emphasize *cis* analyses. This resulted in the exclusion of NUCB2 (narrowly missed p-value threshold), plus FAM3D and LGALS4 (only significant in *cis* + *trans* MR), from our T2D MR results, all of which either play a role in glucose regulation ^50,69^ or have been previously linked to diabetes.^70,71^

Our study has its limitations. First, the trans-ancestry portability problem is commonly observed in biomedical research, including with PGS (see **Supplementary Information**), though we sought to mitigate this by employing multi-ancestry PGS and trans-ancestry analyses. Second, since our study included UKB data, we selected scores that did not make use of the UKB (e.g., relatively older and smaller GWAS) to avoid overfitting, which may attenuate our theoretical statistical power. Third, despite excluding subjects with prevalent cardiometabolic diagnoses, the PGS-related analyses were not immune to reverse causality as several proteins strongly associated with the PGS_T2D_gw_ were implicated in our reverse MR analysis. It is also possible that we are capturing instances of feedback mechanisms or simple Michaelis-Menten kinetics. Fourth, defining phenotypes in the UKB via electronic health records could have an impact on analyses. Notably, we used a biomarker-based definition of NAFLD for performing the GWAS for our MR analysis to boost statistical power but used the ICD10 definition to identify incident cases for mediation analysis. While the biomarker-based definition might not be as robust as a biopsy-based diagnosis, the three most significantly associated loci identified in our NAFLD GWAS (PNPLA3, HSD17B13, TM6SF2) have all been previously associated with NAFLD.^72^ Finally, the PGS, and by extension the mediation analyses using the PGS, could be impacted by horizontal pleiotropy. The same limitation applies to MR analyses using *trans* pQTLs, though we did include a test for pleiotropy in our analytical framework.

Overall, our study elucidated the proteomic signatures of polygenic risk for T2D and comorbidities in both a population-based setting and clinical trials, highlighted the intersections and distinctiveness of the corresponding biological pathways, provided evidence for existing therapeutic and potentially new targets, and constructed an interactive web portal for the broader research community to access our results.

## Data Availability

All data produced in the present study are available upon reasonable request to the authors. Polygenic scores will be uploaded to the PGS Catalog.

## Acknowledgments

We thank the participants and investigators of the UK Biobank study who made this work possible (Resource Application Numbers 26041 and 65851). We are grateful to the research and development leadership teams at the 13 participating UKB-PPP member companies (Alnylam Pharmaceuticals, Amgen, AstraZeneca, Biogen, Bristol-Myers Squibb, Calico, Genentech, Glaxo Smith Klein, Janssen Pharmaceuticals, Novo Nordisk, Pfizer, Regeneron and Takeda) for funding the study. M.I. and S.C.R. were supported by core funding from the British Heart Foundation (RG/18/13/33946), NIHR Cambridge Biomedical Research Centre (BRC-1215-20014; NIHR203312) [*]. MI was also supported by the Cambridge BHF Centre of Research Excellence (RE/18/1/34212), BHF Chair Award (CH/12/2/29428) and by Health Data Research UK (Molecules to Health Records programme), which is funded by the Medical Research Council (UKRI), the National Institute for Health Research, the British Heart Foundation, Cancer Research UK, the Economic and Social Research Council (UKRI), the Engineering and Physical Sciences Research Council (UKRI), Health and Care Research Wales, Chief Scientist Office of the Scottish Government Health and Social Care Directorates, and Health and Social Care Research and Development Division (Public Health Agency, Northern Ireland.

## Conflicts of Interest

D.P.L., M.G., D.M., D.V., X.J., I.A.G., S.P., J.O., A.N., and D.S.P. are employees of AstraZeneca and may hold AstraZeneca stock options. B.B.S. and H.R. are employees of Biogen and may hold stock options. C.D.W. is an employee of Janssen Pharmaceuticals, a Johnson & Johnson company, and may hold stock options. R.R.H. reports personal fees from Anji Pharmaceuticals, AstraZeneca and Novartis. R.J.M. received research support and honoraria from Abbott, American Regent, Amgen, AstraZeneca, Bayer, Boehringer Ingelheim, Boston Scientific, Cytokinetics, Fast BioMedical, Gilead, Innolife, Eli Lilly, Medtronic, Medable, Merck, Novartis, Novo Nordisk, Pfizer, Pharmacosmos, Relypsa, Respicardia, Roche, Rocket Pharmaceuticals, Sanofi, Verily, Vifor, Windtree Therapeutics, and Zoll. M.I. is a trustee of the Public Health Genomics (PHG) Foundation, a member of the Scientific Advisory Board of Open Targets and has research collaborations with Nightingale Health and Pfizer which are unrelated to this study.

## Data and code availability

All results (filtered for a nominal p-value of < 0.05) are available in the supplementary materials, a subset of which are also available via the web portal. The web portal allows download of the underlying data. Requests to access to UK Biobank data can be made here: https://www.ukbiobank.ac.uk/enable-your-research/apply-for-access. Clinical trial data can be accessed following AstraZeneca’s data sharing policies: https://www.astrazenecaclinicaltrials.com/our-transparency-commitments/. All PGS used in the study will be deposited at the PGS Catalog (https://www.pgscatalog.org/) and code used in this study will be deposited on GitHub.

## Contributions

D.P.L., A.N., D.S.P., S.C.R, and M.L. conceptualised this study. D.P.L. performed analyses and drafted the main text and supplementary materials. A.N., D.S.P., and M.L. supervised the study. D.M. and D.V. developed the web portal. M.G. and X.J. assisted with data curation. B.B.S., H.R., C.D.W., and S.P. contributed to the development of the UKB-PPP resource. R.J.M. and R.R.H. led the EXSCEL clinical trial, genotyping, and proteomics data collection. F.A.M, S.D.W., and M.S.S. led DECLARE-TIMI 58 clinical trial and contributed to the genotyping, and proteomics data collection. M.S.U. developed the partitioned T2D polygenic scores. All authors read, commented on, and agreed upon the submitted manuscript.

## Footnotes

* The views expressed are those of the authors and not necessarily those of the NIHR or the Department of Health and Social Care.

## Notes

### Funding Statement

UK Biobank proteomics data was funded by a consortia of 13 participating pharmaceutical companies (Alnylam Pharmaceuticals, Amgen, AstraZeneca, Biogen, Bristol Myers Squibb, Calico, Genentech, GlaxoSmithKline, The Janssen Pharmaceutical Companies of Johnson & Johnson, Novo Nordisk, Pfizer, Regeneron and Takeda). The DECLARE-TIMI 58 and EXSCEL clinical trial were funded by AstraZeneca.

### Author Declarations

Information on EXSCEL and DECLARE-TMI 58 trials can be found on clinicaltrials.gov (NCT01144338 for EXSCEL and NCT01144338 for DECLARE). The trial protocols were approved by institutional review board at each participating site. UK Biobank has approval from the North West Multi-centre Research Ethics Committee (MREC) as a Research Tissue Bank (RTB).

## References

1. Cole JB, Florez JC. Genetics of diabetes and diabetes complications. Nat Rev Nephrol. 2020;16(7):377–390. doi:10.1038/s41581-020-0278-5

2. Meigs JB. The Genetic Epidemiology of Type 2 Diabetes: Opportunities for Health Translation. Curr Diab Rep. 2019;19(8):62. doi:10.1007/s11892-019-1173-y

3. Ahlqvist E, Prasad RB, Groop L. Subtypes of Type 2 Diabetes Determined From Clinical Parameters. Diabetes. 2020;69(10):2086–2093. doi:10.2337/dbi20-0001

4. Iglay K, Hannachi H, Joseph Howie P, et al. Prevalence and co-prevalence of comorbidities among patients with type 2 diabetes mellitus. Curr Med Res Opin. 2016;32(7):1243–1252. doi:10.1185/03007995.2016.1168291

5. Suzuki K, Hatzikotoulas K, Southam L, et al. Multi-ancestry genome-wide study in >2.5 million individuals reveals heterogeneity in mechanistic pathways of type 2 diabetes and complications. Published online March 31, 2023:2023.03.31.23287839. doi:10.1101/2023.03.31.23287839

6. Vujkovic M, Keaton JM, Lynch JA, et al. Discovery of 318 new risk loci for type 2 diabetes and related vascular outcomes among 1.4 million participants in a multi-ancestry meta-analysis. Nat Genet. 2020;52(7):680–691. doi:10.1038/s41588-020-0637-y

7. Mansour Aly D, Dwivedi OP, Prasad RB, et al. Genome-wide association analyses highlight etiological differences underlying newly defined subtypes of diabetes. Nat Genet. 2021;53(11):1534–1542. doi:10.1038/s41588-021-00948-2

8. Khera AV, Chaffin M, Aragam KG, et al. Genome-wide polygenic scores for common diseases identify individuals with risk equivalent to monogenic mutations. Nat Genet. 2018;50(9):1219–1224. doi:10.1038/s41588-018-0183-z

9. Yun JS, Jung SH, Shivakumar M, et al. Polygenic risk for type 2 diabetes, lifestyle, metabolic health, and cardiovascular disease: a prospective UK Biobank study. Cardiovasc Diabetol. 2022;21(1):131. doi:10.1186/s12933-022-01560-2

10. Udler MS, Kim J, von Grotthuss M, et al. Type 2 diabetes genetic loci informed by multi-trait associations point to disease mechanisms and subtypes: A soft clustering analysis. PLoS Med. 2018;15(9):e1002654. doi:10.1371/journal.pmed.1002654

11. DiCorpo D, LeClair J, Cole JB, et al. Type 2 Diabetes Partitioned Polygenic Scores Associate With Disease Outcomes in 454,193 Individuals Across 13 Cohorts. Diabetes Care. 2022;45(3):674–683. doi:10.2337/dc21-1395

12. Kim H, Westerman KE, Smith K, et al. High-throughput genetic clustering of type 2 diabetes loci reveals heterogeneous mechanistic pathways of metabolic disease. Diabetologia. 2023;66(3):495–507. doi:10.1007/s00125-022-05848-6

13. Ritchie SC, Lambert SA, Arnold M, et al. Integrative analysis of the plasma proteome and polygenic risk of cardiometabolic diseases. Nat Metab. 2021;3(11):1476–1483. doi:10.1038/s42255-021-00478-5

14. Steffen BT, Tang W, Lutsey PL, et al. Proteomic analysis of diabetes genetic risk scores identifies complement C2 and neuropilin-2 as predictors of type 2 diabetes: the Atherosclerosis Risk in Communities (ARIC) Study. Diabetologia. 2023;66(1):105–115. doi:10.1007/s00125-022-05801-7

15. Sun BB, Chiou J, Traylor M, et al. Plasma proteomic associations with genetics and health in the UK Biobank. Nature. 2023;622(7982):329-338. doi:10.1038/s41586-023-06592-6

16. Dhindsa RS, Burren OS, Sun BB, et al. Rare variant associations with plasma protein levels in the UK Biobank. Nature. 2023;622(7982):339-347. doi:10.1038/s41586-023-06547-x

17. Holman RR, Bethel MA, Mentz RJ, et al. Effects of Once-Weekly Exenatide on Cardiovascular Outcomes in Type 2 Diabetes. N Engl J Med. 2017;377(13):1228–1239. doi:10.1056/NEJMoa1612917

18. Wiviott SD, Raz I, Bonaca MP, et al. Dapagliflozin and Cardiovascular Outcomes in Type 2 Diabetes. N Engl J Med. 2019;380(4):347–357. doi:10.1056/NEJMoa1812389

19. Bycroft C, Freeman C, Petkova D, et al. The UK Biobank resource with deep phenotyping and genomic data. Nature. 2018;562(7726):203-209. doi:10.1038/s41586-018-0579-z

20. Canela-Xandri O, Rawlik K, Tenesa A. An atlas of genetic associations in UK Biobank. Nat Genet. 2018;50(11):1593–1599. doi:10.1038/s41588-018-0248-z

21. Littlejohns TJ, Holliday J, Gibson LM, et al. The UK Biobank imaging enhancement of 100,000 participants:lllrationale, data collection, management and future directions. Nat Commun. 2020;11(1):2624. doi:10.1038/s41467-020-15948-9

22. Wang Q, Dhindsa RS, Carss K, et al. Rare variant contribution to human disease in 281,104 UK Biobank exomes. Nature. 2021;597(7877):527-532. doi:10.1038/s41586-021-03855-y

23. Zhu L, Fang Z, Jin Y, et al. Association between serum alanine and aspartate aminotransferase and blood pressure: a cross-sectional study of Chinese freshmen. BMC Cardiovasc Disord. 2021;21:472. doi:10.1186/s12872-021-02282-1

24. Austin PC, Lee DS, Fine JP. Introduction to the Analysis of Survival Data in the Presence of Competing Risks. Circulation. 2016;133(6):601–609. doi:10.1161/CIRCULATIONAHA.115.017719

25. Ge T, Chen CY, Ni Y, Feng YCA, Smoller JW. Polygenic prediction via Bayesian regression and continuous shrinkage priors. Nat Commun. 2019;10(1):1776. doi:10.1038/s41467-019-09718-5

26. Ruan Y, Lin YF, Feng YCA, et al. Improving Polygenic Prediction in Ancestrally Diverse Populations. Nat Genet. 2022;54(5):573–580. doi:10.1038/s41588-022-01054-7

27. Vujkovic M, Ramdas S, Lorenz KM, et al. A multiancestry genome-wide association study of unexplained chronic ALT elevation as a proxy for nonalcoholic fatty liver disease with histological and radiological validation. Nat Genet. 2022;54(6):761–771. doi:10.1038/s41588-022-01078-z

28. Chang CC, Chow CC, Tellier LC, Vattikuti S, Purcell SM, Lee JJ. Second-generation PLINK: rising to the challenge of larger and richer datasets. GigaScience. 2015;4:7. doi:10.1186/s13742-015-0047-8

29. Steen J, Loeys T, Moerkerke B, Vansteelandt S. medflex: An R Package for Flexible Mediation Analysis using Natural Effect Models. J Stat Softw. 2017;76:1–46. doi:10.18637/jss.v076.i11

30. Rasooly D, Peloso GM, Pereira AC, et al. Genome-wide association analysis and Mendelian randomization proteomics identify drug targets for heart failure. Published online May 26, 2023:2022.04.14.22273877. doi:10.1101/2022.04.14.22273877

31. Yavorska OO, Burgess S. MendelianRandomization: an R package for performing Mendelian randomization analyses using summarized data. Int J Epidemiol. 2017;46(6):1734–1739. doi:10.1093/ije/dyx034

32. Bowden J, Davey Smith G, Burgess S. Mendelian randomization with invalid instruments: effect estimation and bias detection through Egger regression. Int J Epidemiol. 2015;44(2):512–525. doi:10.1093/ije/dyv080

33. Raudvere U, Kolberg L, Kuzmin I, et al. g:Profiler: a web server for functional enrichment analysis and conversions of gene lists (2019 update). Nucleic Acids Res. 2019;47(W1):W191–W198. doi:10.1093/nar/gkz369

34. Mak KH, Vidal-Petiot E, Young R, et al. Prevalence of diabetes and impact on cardiovascular events and mortality in patients with chronic coronary syndromes, across multiple geographical regions and ethnicities. Eur J Prev Cardiol. 2022;28(16):1795–1806. doi:10.1093/eurjpc/zwab011

35. Einarson TR, Acs A, Ludwig C, Panton UH. Prevalence of cardiovascular disease in type 2 diabetes: a systematic literature review of scientific evidence from across the world in 2007-2017. Cardiovasc Diabetol. 2018;17(1):83. doi:10.1186/s12933-018-0728-6

36. Coral DE, Fernandez-Tajes J, Tsereteli N, et al. A phenome-wide comparative analysis of genetic discordance between obesity and type 2 diabetes. Nat Metab. 2023;5(2):237–247. doi:10.1038/s42255-022-00731-5

37. Hirano T. Pathophysiology of Diabetic Dyslipidemia. J Atheroscler Thromb. 2018;25(9):771–782. doi:10.5551/jat.RV17023

38. Hall JE, Mouton AJ, da Silva AA, et al. Obesity, kidney dysfunction, and inflammation: interactions in hypertension. Cardiovasc Res. 2020;117(8):1859–1876. doi:10.1093/cvr/cvaa336

39. Martin AR, Kanai M, Kamatani Y, Okada Y, Neale BM, Daly MJ. Clinical use of current polygenic risk scores may exacerbate health disparities. Nat Genet. 2019;51(4):584–591. doi:10.1038/s41588-019-0379-x

40. Vulf M, Bograya M, Komar A, et al. NGR4 and ERBB4 as Promising Diagnostic and Therapeutic Targets for Metabolic Disorders. Front Biosci Elite Ed. 2023;15(2):14. doi:10.31083/j.fbe1502014

41. Chiang KM, Chang HC, Yang HC, et al. Genome-wide association study of morbid obesity in Han Chinese. BMC Genet. 2019;20(1):97. doi:10.1186/s12863-019-0797-x

42. Salinas YD, Wang L, DeWan AT. Multiethnic genome-wide association study identifies ethnic-specific associations with body mass index in Hispanics and African Americans. BMC Genet. 2016;17(1):78. doi:10.1186/s12863-016-0387-0

43. Santiago-Marrero I, Liu F, Wang H, Arzola EP, Xiong WC, Mei L. Energy Expenditure Homeostasis Requires ErbB4, an Obesity Risk Gene, in the Paraventricular Nucleus. eNeuro. 2023;10(9):ENEURO.0139-23.2023. doi:10.1523/ENEURO.0139-23.2023

44. Pickrell JK, Berisa T, Liu JZ, Ségurel L, Tung JY, Hinds DA. Detection and interpretation of shared genetic influences on 42 human traits. Nat Genet. 2016;48(7):709–717. doi:10.1038/ng.3570

45. Liu ZW, Ma Q, Liu J, Li JW, Chen YD. The association between plasma furin and cardiovascular events after acute myocardial infarction. BMC Cardiovasc Disord. 2021;21:468. doi:10.1186/s12872-021-02029-y

46. Wang YK, Tang JN, Han L, et al. Elevated FURIN levels in predicting mortality and cardiovascular events in patients with acute myocardial infarction. Metabolism. 2020;111:154323. doi:10.1016/j.metabol.2020.154323

47. Mishra A, Malik R, Hachiya T, et al. Stroke genetics informs drug discovery and risk prediction across ancestries. Nature. 2022;611(7934):115-123. doi:10.1038/s41586-022-05165-3

48. Cao T, Yang D, Zhang X, et al. FAM3D inhibits glucagon secretion via MKP1-dependent suppression of ERK1/2 signaling. Cell Biol Toxicol. 2017;33(5):457–466. doi:10.1007/s10565-017-9387-8

49. Zhang X, Yang W, Wang J, Meng Y, Guan Y, Yang J. FAM3 gene family: A promising therapeutical target for NAFLD and type 2 diabetes. Metabolism. 2018;81:71–82. doi:10.1016/j.metabol.2017.12.001

50. Moser C, Gosselé KA, Balaz M, et al. FAM3D: A gut secreted protein and its potential in the regulation of glucose metabolism. Peptides. 2023;167:171047. doi:10.1016/j.peptides.2023.171047

51. Breit SN, Brown DA, Tsai VWW. The GDF15-GFRAL Pathway in Health and Metabolic Disease: Friend or Foe? Annu Rev Physiol. 2021;83:127–151. doi:10.1146/annurev-physiol-022020-045449

52. Wang D, Day EA, Townsend LK, Djordjevic D, Jørgensen SB, Steinberg GR. GDF15: emerging biology and therapeutic applications for obesity and cardiometabolic disease. Nat Rev Endocrinol. 2021;17(10):592–607. doi:10.1038/s41574-021-00529-7

53. Xu R, Tan H, Zhang J, Yuan Z, Xie Q, Zhang L. Fam20C in Human Diseases: Emerging Biological Functions and Therapeutic Implications. Front Mol Biosci. 2021;8:790172. doi:10.3389/fmolb.2021.790172

54. Narayanan RP, Fu B, Heald AH, et al. IGFBP2 is a biomarker for predicting longitudinal deterioration in renal function in type 2 diabetes. Endocr Connect. 2012;1(2):95–102. doi:10.1530/EC-12-0053

55. Anliker-Ort M, Dingemanse J, van den Anker J, Kaufmann P. Treatment of Rare Inflammatory Kidney Diseases: Drugs Targeting the Terminal Complement Pathway. Front Immunol. 2020;11:599417. doi:10.3389/fimmu.2020.599417

56. Emmens RW, Wouters D, Zeerleder S, van Ham SM, Niessen HWM, Krijnen PAJ. On the value of therapeutic interventions targeting the complement system in acute myocardial infarction. Transl Res J Lab Clin Med. 2017;182:103–122. doi:10.1016/j.trsl.2016.10.005

57. Patriquin CJ, Kuo KHM. Eculizumab and Beyond: The Past, Present, and Future of Complement Therapeutics. Transfus Med Rev. 2019;33(4):256–265. doi:10.1016/j.tmrv.2019.09.004

58. Schartz ND, Tenner AJ. The good, the bad, and the opportunities of the complement system in neurodegenerative disease. J Neuroinflammation. 2020;17(1):354. doi:10.1186/s12974-020-02024-8

59. Sundararaman SS, Döring Y, van der Vorst EPC. PCSK9: A Multi-Faceted Protein That Is Involved in Cardiovascular Biology. Biomedicines. 2021;9(7):793. doi:10.3390/biomedicines9070793

60. Kamstrup PR. Lipoprotein(a) and Cardiovascular Disease. Clin Chem. 2021;67(1):154-166. doi:10.1093/clinchem/hvaa247

61. Alagarsamy J, Jaeschke A, Hui DY. Apolipoprotein E in Cardiometabolic and Neurological Health and Diseases. Int J Mol Sci. 2022;23(17):9892. doi:10.3390/ijms23179892

62. Boughanem H, Yubero-Serrano EM, López-Miranda J, Tinahones FJ, Macias-Gonzalez M. Potential Role of Insulin Growth-Factor-Binding Protein 2 as Therapeutic Target for Obesity-Related Insulin Resistance. Int J Mol Sci. 2021;22(3):1133. doi:10.3390/ijms22031133

63. Zhang T, Zhang Y, Tao J, Rong X, Yang Y. Intestinal Trefoil Factor 3: a new biological factor mediating gut-kidney crosstalk in diabetic kidney disease. Endocrine. Published online December 26, 2023. doi:10.1007/s12020-023-03559-5

64. Zhi K, Yin R, Guo H, Qu L. PUM2 regulates the formation of thoracic aortic dissection through EFEMP1. Exp Cell Res. 2023;427(2):113602. doi:10.1016/j.yexcr.2023.113602

65. Li W, Lou X, Zha Y, et al. Single-cell RNA-seq of heart reveals intercellular communication drivers of myocardial fibrosis in diabetic cardiomyopathy. eLife. 2023;12:e80479. doi:10.7554/eLife.80479

66. Jasra SK, Badian C, Macri I, Ra P. Recognition of Early Myocardial Infarction by Immunohistochemical Staining with Cardiac Troponin-I and Complement C9*. J Forensic Sci. 2012;57(6):1595–1600. doi:10.1111/j.1556-4029.2012.02172.x

67. Huo Y, Lai Y, Feng Q, Wang Q, Li J. Serum ITIH4 in coronary heart disease: a potential anti-inflammatory biomarker related to stenosis degree and risk of major adverse cardiovascular events. Biomark Med. 2022;16(18):1279–1288. doi:10.2217/bmm-2022-0673

68. Pretorius E, Mbotwe S, Kell DB. Lipopolysaccharide-binding protein (LBP) reverses the amyloid state of fibrin seen in plasma of type 2 diabetics with cardiovascular co-morbidities. Sci Rep. 2017;7(1):9680. doi:10.1038/s41598-017-09860-4

69. Nakata M, Yada T. Role of NUCB2/nesfatin-1 in glucose control: diverse functions in islets, adipocytes and brain. Curr Pharm Des. 2013;19(39):6960–6965. doi:10.2174/138161281939131127130112

70. Dieden A, Gudmundsson P, Korduner J, et al. Galectin-4 is associated with diabetes and obesity in a heart failure population. Sci Rep. 2023;13(1):20285. doi:10.1038/s41598-023-47426-9

71. Li XS, Yan CY, Fan YJ, Yang JL, Zhao SX. NUCB2 polymorphisms are associated with an increased risk for type 2 diabetes in the Chinese population. Ann Transl Med. 2020;8(6):290. doi:10.21037/atm.2020.03.02

72. Trépo E, Valenti L. Update on NAFLD genetics: From new variants to the clinic. J Hepatol. 2020;72(6):1196–1209. doi:10.1016/j.jhep.2020.02.020

