## Supplemental Information for "Identification of plasma proteomic markers underlying polygenic risk of type 2 diabetes and related comorbidities"

### **Extended Methods**

*Genotyping quality control and imputation*

EXSCEL was genotyped using the Human Global Screening Array-MD-24 Beadchips v2.0 (GSA) and DECLARE was genotyped using the Infinium Global Screening Array-24 v3.0 GSA + Multi Disease (GSAMD). Quality control (QC) for imputation was performed on the genotype data using PLINK v2.00a4LM^1^, removing samples with high heterozygosity (> 4 SD from the mean), discordance between reported and predicted sex, or a genotyping missingness rate > 0.05. Variants were removed if they had a HWE p-value < 1x10^-12^, a missingness rate > 0.05, or a minor allele frequency (MAF) < 0.001. Ancestry and relationship inference was conducted using KING 2.3.0^2^ and 1000 Genomes Project data (1KGP) as an ancestral reference panel.^3,4^ Subjects were assigned to a 1KGP super-population based on a genetic ancestry probability cut-off of >= 0.90. Unless otherwise stated, relative pairs were resolved to the second-degree using KING-generated kinship coefficients. Imputation in EXSCEL and DECLARE was then conducted using BEAGLE 4.0^5^ and the high coverage 1KGP data as the imputation reference panel in a pipeline modelled after the FinnGen imputation pipeline.^4,6^ UKB genotyping and imputation has been previously described.^7^ For the estimation of polygenic scores in EXSCEL, DECLARE, and UKB, imputed variants were excluded if they had a HWE p-value < 1x10^-6^ within a given ancestry, a MAF < 0.01, a missingness > 0.05, and an imputation INFO score < 0.5. Relationship inference in UKB was also conducted using KING and ancestry inference was previously performed using PEDDY 0.4.2^8^ with the 1KGP as the ancestral reference panel. KING and PEDDY methods for ancestry inferences are highly similar, both using 1KGP-based principal components and a SVM classifier, resulting in consistent ancestry classification as confirmed on a subset of UKB participants.

*Proteomics quality control*

The UKB PPP proteomics data were generated using the Olink Explore platform. Quality control was performed by the UKB-PPP consortium and is described in Sun et al. 2022.^9^ For EXSCEL, proteomics data were generated for 2,949 genotyped individuals using the SomaScan assay at baseline and the 12-month timepoint. DECLARE-TIMI 58 proteomics data were generated for 934 genotyped individuals at the baseline and the 6-month timepoints using the Olink Target 96 Cardiovascular II, Cardiovascular III, Cardiometabolic panels. Proteomics quality control for the UKB was performed by the UKB-PPP consortium.^9^ We followed their protocol to standardise quality control in EXSCEL and DECLARE. Samples were removed if their median expression level, IQR, or standardized PCs 1 and 2 of the corresponding proteome data were greater than 5 SD from the mean. Samples were also removed if there were any associated assay or quality control warnings issued by the company generating the proteomics data. For DECLARE, the Olink-generated normalized protein expression (npx) levels were rank-based inverse normalized. Note that covariates were not regressed from the npx values. For EXSCEL, the SomaScan data were processed using an approach modelled after Sun et al. 2018^10^, i.e., protein expression data were first converted to the log10 scale, followed by regression of age, sex, PCs 1-10, and time from sample collection, and finally rank-based inverse normalized.

*PGS Validation*

To validate the PGS, we tested their associations with traits defined using ICD10 codes and/or biomarkers (**Supplementary Table 2**, **Supplementary Figure 3-8**) in all unrelated UKB-PPP participants (resolved to the 2^nd^ degree using KING 2.3.0^2^) and again after stratifying the cohort by predicted ancestry using R and adjusting for age, age^2^, sex, age*sex, age^2^*sex, UKB centre, array, and PCs 1-20. For the CAD, BMI, and T2D, PRS-CSx inferred three sets of PGS (a “European” PGS, an “East Asian” PGS, and a meta-analysis of the two). In these cases, we retained the PGS with the strongest association with CAD, BMI, and T2D in the all-participants analysis. Note for the NAFLD PGS, we evaluated against four definitions of NAFLD/NASH to determine robustness: the ICD10 code K76.0, APRI (Aspartate to Platelet Ratio Index) score > 0.7 (NAFLD_APRI); alanine aminotransferase (ALT) and aspartate aminotransferase (AST) > 40 U/L (NAFLD_AST_ALT); and Fibrosis-4 index > 3.25 (NAFLD_FIB4).

*GWAS and pQTL summary statistics for Mendelian randomization*

We extracted conditionally independent pQTLs from the pQTL GWAS summary statistics generated by the UKB PPP consortium using the European ancestry 35K discovery subset to use as instruments for our two-sample Mendelian randomization (MR) analysis. All pQTLs had a p-value < 3.4x10^-11^, the study-wide significant threshold used by the pQTL GWAS.^9^ The pQTLs were defined as *cis* if they were within +/- 1MB of a gene’s transcription start site. For this same set of pQTLs, we also extracted summary statistics using the pan-ancestry combined UKB-PPP cohort for our two-sample trans-ancestry MR analysis.

We then generated trait-level GWAS summary statistics that excluded UKB-PPP participants. Individuals were retained for the GWAS if they passed an internal set of QC criteria including sex concordance, heterozygosity, ploidy, were assigned a continental PEDDY-predicted^8^ ancestry with a probability >= 0.90, and were not included in the UKB-PPP study. We utilized REGENIE for the GWAS, which employs a two-step approach as previously described.^11^ For the first step, we used quality-controlled genotyped variants (MAF >1%, genotyping rate >99%, HWE p-value >10^−15^, <10% missingness and LD pruning using 1000 variant windows, 100 sliding windows and r^2^<0.8), while for the second step we used imputed variants with a MAC > 50 and an INFO score > 0.7. For traits assessed in the full cohort (T2D, BMI, CKD, CAD, NAFLD), we performed each GWAS within each of the predicted continental ancestries, adjusting for age, sex, and PCs 1-10, and (+/-) BMI. We then meta-analysed each GWAS using a fixed-effects meta-analysis. For comorbidities and complications assessed in prevalent diabetics (Supplementary Table 2), we only used predicted European-ancestry subjects due to sample size concerns.

*Mendelian randomization sensitivity analyses*

For sensitivity analyses, we repeated MR analyses using both *cis* and *trans* pQTLs. We also performed both single-ancestry (using only European-ancestry summary statistics) and pan-ancestry (using pan-ancestry summary statistics from the full UKB-PPP cohort and the meta-analysed trans-ancestry GWAS) MR analyses. We then performed reverse MR in the same manner as above, but with the exposure and outcome swapped. Finally, for any variant with GWAS p-value < 1x10^-6^ for both the trait and the protein, we conducted a colocalization analysis using the coloc package (version 5.1.0.1) in R 4.2.2 on a +/- 250 kilobase window centred on the variant. We used the summary statistics-based method (coloc.abf) and considered the GWAS variant and the pQTL to colocalize following recommended criteria (PP.H3 +PP.H4 > 0.99 and PP.H4/PP.H3 > 5).^12–14^ For loci that harboured multiple significant loci for the trait GWAS (p-value < 1x10^-6^), we also used the coloc.susie^15^ method with LD matrices computed with the imputed UKB-PPP data and PLINK v1.90b6.18.^1,16^

*Description of risk factors included in clinical trial time-to-event analyses*

In the case of statistically significant associations in the clinical trial time to event analyses, (Bonferroni corrected p-value < 0.05), we assessed the proteins using more comprehensive models to evaluate their suitability as a biomarker. These models included both the above covariates along with additional risk factors specific to the outcome of interest. For heart failure outcomes, we also adjusted for coronary artery disease, atrial fibrillation, baseline eGFR, baseline UACR (diagnosis of albuminuria for EXSCEL), and prior heart failure.^17^ For MACE, we also adjusted for hypertension, hypercholesterolemia, obesity (BMI >= 30 kg/m²), smoking (current, or smoking cessation ≤3 months), positive family history (parent or sibling with cardiovascular disease before age 65), and atherosclerotic disease (prior myocardial infarction, percutaneous coronary intervention/coronary artery bypass grafting, cerebrovascular accident/transient ischemic attack, or peripheral arterial disease).^18^ For renal outcomes, we adjusted for atherosclerotic cardiovascular disease at baseline, heart failure at baseline, systolic blood pressure at baseline, T2D duration, HbA1c at baseline, eGFR at baseline, urine albumin-to-creatinine ratio (ACR) at baseline, and haemoglobin at baseline.^19^ Note for EXSCEL, certain analytes were only assessed intermittently, resulting in sample size loss when adjusting for these covariates. Consequently, we replaced ACR with a baseline diagnosis of albuminuria and performed a sensitivity analysis with and without haemoglobin. Despite the substantial difference in sample sizes, the hazard ratios were highly correlated (r=0.91, p-value < 2.2x10^-16^), thus we omitted the haemoglobin adjustment for EXSCEL. For insulin initiation, we adjusted for cardiovascular disease, CKD, number of diabetic complications (e.g., diabetic retinopathy, neuropathy, amputations), and duration of diabetes.^20^ Finally, for cardiovascular outcomes, we included a third model that also adjusted for NT-proBNP to identify proteins that were independent of this biomarker.

*PGS-protein associations in randomized controlled trials*

We tested all available proteins for their associations with the PGS in EXSCEL and DECLARE, adjusting for age, age^2^, sex, age*sex, age^2^*sex, time from sample collection (if available), batch (if available), PCs1-10, and (+/-) BMI using the protein expression data measured at baseline prior to randomization. As a sensitivity analysis, we first included all study participants in the analyses followed by restricting them to only inferred European-ancestry participants. We applied an FDR correction to the resultant p-values.

*Association of PGS with outcomes in randomized controlled trials*

We tested PGS for their association with the time to EXSCEL and DECLARE outcomes with the survival package in R 3.6.1 (<https://github.com/therneau/survival>). We used cox proportional hazards regression and adjusted for age, sex, age*sex, PCs1-10, trial arm, and (+/-) BMI.

### **Discordance between T2D and cardiometabolic PGS-protein associations**

As T2D status confers risk for many other cardiometabolic disorders, the expectation would be that most cardiometabolic PGS-protein associations would be concordant in their direction of effect, as evidenced by their high overall correlation. However, for a select set of proteins, their directions of effect were in fact opposite with that of the PGS_T2D_gw_ (**Supplementary Figure 19 A, Supplementary Table 8)**. Overall, 217 proteins were discordant between the PGS_T2D_gw_ and at least one other PGS (p-value < 5.2x10^-6^ for both scores).

In total, 5 proteins were with PGS_BMI_ (C7, CD300LG, GFRAL, SERPINA4, TIMP4, TTR), 1 protein for the PGS_CAD_ (PLA2G7), 4 with the PGS_NALFD_ (BCHE, ENTPD5, HSD11B1, KDR, KLKB1, ORM1, PALM, PLA2G7), and 65 with the PGS_CKD_ (including proteins such as UMOD and MANSC4). We then evaluated whether the discordance identified via PGS are reflected in the associations of the circulating protein with disease status. Overall, 82.5% of proteins identified via PGS discordance were also discordant in their associations with disease status (**Supplementary Figure 19 B**).

Discordances in protein associations between the PGS_T2D_gw_ and the partitioned T2D scores are notable considering both sets of scores are comprised of T2D risk variants. For the partitioned scores, 2 proteins were discordant between the PGS_T2D_gw_ and the PGS_T2D_beta_cell_ (LGALS4 and SELE). Another 2 proteins were discordant between the PGS_T2D_gw_ and the PGS_T2D_lipodystophy_ (CLMP and LEP). The PGS_T2D_lipodystophy_ is associated with lower BMI, so the negative association with LEP (leptin) and CLMP (adrenomedullin) seem to follow that pattern as both proteins have known associations with increased BMI.^21,22^ Given the overall negative correlation of PGS_T2D_gw_ and PGS_T2D_liver_lipid_ beta coefficients, we unsurprisingly found 172 proteins to be discordant between the T2D PGS and the liver lipid pPS. Again, this suggests a different molecular signature for the liver lipid pPS, and by proxy, *GCKR*.

### **Cross-ancestry comparisons of PGS-protein associations**

Due to the longstanding biases by ancestry in GWAS datasets and biobanks^23^, the cross-ancestry portability of PGS analyses remains a concern. When possible, we used multi-ancestry GWAS summary statistics for our PGS estimation and PRS-CSx, which is able to leverage the linkage disequilibrium structure of multiple GWAS populations to better model multi-ancestry PGS.^24^ PRS-CSx estimates a set of posterior effect sizes for each GWAS ancestry and a fixed effects meta-analysis of these posterior effect sizes, and we retained the PGS with the strongest association with the target trait in the multi-ancestry subset of the UKB-PPP cohort. However, the PGS effect sizes on the target trait were generally greater in European-ancestry UKB-PPP participants versus the other ancestries (supplementary table 6), reflecting both lower statistical power and the PGS capturing less trait variance in non-European populations. For example, the PGS_T2D_gw_ had an odds ratio (OR) of 2.41 in European-ancestry participants for T2D compared to an OR of 1.76 in South Asian participants and 1.42 in African ancestry participants.

Furthermore, the UKB-PPP, like the UKB at large, is predominantly European-ancestry (92%). The PGS only captured at most 1.5% of trait variance for protein expression levels; such associations were not likely to be detected given small sample sizes of non-European UKB-PPP participants. Consequently, we instead compared effect sizes of the PGS in genetically predicted European versus non-European cohorts (supplementary table 9). The two largest subsets were genetically-predicted South Asian (n=820) and African (n=1,126) participants. The PGS_T2D_gw_ effect sizes had a correlation of 0.58 (p-value =9.5x10^-268^) in South Asian versus European UKB-PPP participants, which improved to 0.75 when only including the 695 proteins that significantly replicated (p-value = 5.2x10^-126^). The other cardiometabolic PGS performed similarly except for the PGS_CKD_ where the correlations remained relatively low (r=0.20 for all proteins; R=0.23 for replicated proteins). For the partitioned T2D scores, the PGS_T2D_proinsulin_ effect sizes were negatively correlated, albeit weakly (r=-0.06, p-value = 1.3x10^-3^). The correlation of the remaining scores’ effect sizes ranged from 0.24 (PGS_T2D_obesity_) to 0.42 (PGS_T2D_lipodystrophy)_. Like the other cardiometabolic scores, these correlations improved dramatically when only examining the replicated PGS-protein associations, most notability the PGS_T2D_beta_cell_ improving to 0.92 (p-value = 2.9x10^-11^).

In the predicated African-ancestry subset of the UKB-PPP, the correlations of the PGS effect sizes with European-ancestry participants were notability lower, which fits with previous observations regarding how the PGS translate across ancestries.^23^ The PGS_T2D_gw_, for example, had a correlation of 0.27 (p-value = 6x10^-52^), though again this was improved by subsetting to only replicated PGS-protein associations (r=0.41, p=2.6x10^-29^). Similar patterns were observed for the other cardiometabolic PGS, though the PGS_CKD_ effect sizes more correlated between African and European participants (r=0.39, p-value = 3.7x10^-105^, all proteins) than between South Asian and European participants (see above). The effect size correlations of the five partitioned T2D scores were highly variable, with two modestly correlated (PGS_T2D_proinsulin_=0.07 and PGS_T2D_obesity_=0.1), one negatively correlated (PGS_T2D_beta_cell_=-0.05), and two not correlated (p-value > 0.05 for PGS_T2D_lipodystrophy_ and PGS_T2D_liver_lipid_). Not including the PGS_T2D_proinsulin_, correlations were dramatically improved by only including significant PGS-protein associations (see Supplementary Figures 19-23).

These results reflect previous studies showing that the reduction in PGS predictive accuracy reflects demographic history with populations more genetically similar to European-ancestry populations (e.g. South Asian) compared to less genetically similar populations (e.g., African ancestry).^23^ This stems from the European-ancestry bias in GWAS data. Despite including non-European GWAS, most cases underlying our PGS were still European. The demographics of the UKB-PPP further limited our ability to characterize non-European PGS-protein associations. However, statistically-significant PGS-protein association were more likely to have similar effect sizes across populations, ideally indicating that we are capturing true biological associations. Nevertheless, replicating these findings in a well-powered non-European cohort and increasing diversity in omics data overall remains an area of active development.

### **PGS associations with circulating proteins in patients with diabetes (UKB, EXSCEL, DECLARE-TIMI 58)**

We sought to explore how PGS inform protein levels in participants with established disease, using both the UKB and independent studies, and to what extent our findings would be generalisable in a clinical trial setting. In total, 1972 proteins overlap between EXSCEL and UKB. Of these, 928 proteins were significantly associated with a PGS in UKB, and 98 protein-PGS associations significantly replicated after adjusting for multiple testing (**Supplementary Table 10**). To compare the PGS effect sizes on circulating proteins in the clinical trials to that of the UKB, we utilized UKB-PPP diabetic participants as they more closely resembled the trial population and estimated the correlation of effect sizes for each score in the UKB and EXSCEL (e.g., the correlation of PGS_BMI_ effect sizes in EXSCEL and UKB). The PGS_BMI_, PGS_CKD_, PGS_NAFLD_, and PGS_T2D_liver_lipid_ effect sizes were highly correlated between UKB and EXSCEL (Pearson’s r > 0.32 and p-values < 0.05; **Supplementary Table 11, Supplementary Figure 24**). The proinsulin pPS was the only score where the effect sizes were not significantly correlated between EXSCEL and the UKB. Of the 276 proteins available in DECLARE, 206 were significantly associated with a PGS in UKB. Of these, 38 PGS-protein associations significantly replicated after adjusting for multiple tests (**Supplementary Table 12**). The PGS_BMI_, PGS_CKD_, PGS_NAFLD_, PGS_T2D_beta_cell_, and PGS_T2D_liver_lipid_ effect sizes on circulating proteins were highly correlated (Pearson’s r > 0.42, p < 0.05; **Supplementary Table 13, Supplementary Figure 25**). Out of the scores we evaluated in DECLARE and UKB, only the PGS_T2D_obesity_ effect sizes were not significantly correlated.

While we have substantially lower power when analysing PGS for protein associations in diabetics compared to the full UKB cohort, it can be informative to identify what proteins remain significant (**Supplementary Table 14**). For incident diabetics, the PGS_T2D_gw_ was not significantly associated with any proteins, but the GWAS-significant PGS_T2D_gwas_ was associated with 129 proteins (3 after BMI adjustment: ADAMTS16, CCL19, and PAM). The PGS_CKD_, PGS_NAFLD_, PGS_BMI_, and PGS_T2D_liver_lipid_ were all associated with 48 or more proteins. The PGS_CAD_ was significantly associated with LPA, the PGS_T2D_lipodystrophy_ was associated with CKB and IGFBP2 (both significant after BMI adjustment), and the PGS_T2D_beta_cell_ was significantly associated with CFC1, LEP, and RN4R (none after BMI adjustment). In prevalent diabetics, the association patterns differed somewhat. The PGS_T2D_gw_ was significantly associated with 37 proteins while the GWAS-significant T2D score was associated with 148 proteins. The PGS_T2D_lipodystrophy_ was instead associated with LEP and the beta cell pPS was associated with ABO, CD209, FGA, and SELE. The PGS_T2D_liver_lipid_ was associated with fewer proteins in prevalent diabetics (CD4, CHI3L1, LDL4, LEP4, and PCSK9) compared to 48 proteins in incident diabetics. The difference in power of the genome-wide PGS_T2D_gw_ versus the PGS_T2D_gwas_ to detect protein associations in diabetics stands in contrast to its power in the full UKB cohort (non-diabetics) and its overall association with diabetes (**Supplementary** **Figure 3**).

### **PGS associations with outcomes in randomized controlled trials (EXSCEL and DECLARE)**

To evaluate whether PGS influenced clinical trial outcomes in addition to circulating protein levels, we tested each PGS for association with the time to events for MACE, hospitalization for heart failure, a renal composite endpoint, and insulin initiation; **Supplementary Figures 32 and 33, Supplementary Table 19 and 20**). For this analysis, we used all genotyped individuals in EXSCEL (N=4,564) and DECLARE (N=12,620). In DECLARE, the PGS_CAD_ was significantly associated with the MACE endpoint (HR=1.2, Adj. P= 8.6x10^-8^), the PGS_BMI_ was significantly associated with the hospitalization for heart failure endpoint (HR=1.3, Adj. P=7.2x10^-5^), and the PGS_T2D_gw_ was significantly associated with the insulin initiation endpoint (HR=0.91, Adj. P=0.04). In EXSCEL, the PGS_CAD_ was significantly associated with MACE (HR=1.81, adj. p-value = 0.001). Note that the partitioned scores were not significantly associated with the four clinical trial outcomes we evaluated.

We found the direction of effect for the PGS_T2D_gw_ with the insulin initiation outcome in DECLARE to be contrary to expectations. Therefore, we tested the PGS_T2D_gw_ for association with insulin use at baseline, diabetic neuropathy at baseline, and diabetic retinopathy at baseline. The PGS_T2D_gw_ was significantly associated for all three complications in both DECLARE (p-value < 0.004) and EXSCEL (p-value < 0.01). For each complication, there was a positive direction of effect where participants with a higher PGS_T2D_gw_ were more likely to have that diabetic complication. Also, as the PGS_CKD_ was not significantly associated with the renal outcome in either trial, we also tested it for association with baseline eGFR, confirming that the PGS_CKD_ functioned as expected (DECLARE: beta=-3.16, p-value = 2x10^-68^; EXSCEL: beta=-3.03, p-value = 1.1x10^-14^).

### **Supplementary Information References**

1.	Chang CC, Chow CC, Tellier LC, Vattikuti S, Purcell SM, Lee JJ. Second-generation PLINK: rising to the challenge of larger and richer datasets. *GigaScience*. 2015;4:7. doi:10.1186/s13742-015-0047-8

2.	Manichaikul A, Mychaleckyj JC, Rich SS, Daly K, Sale M, Chen WM. Robust relationship inference in genome-wide association studies. *Bioinformatics*. 2010;26(22):2867-2873. doi:10.1093/bioinformatics/btq559

3.	1000 Genomes Project Consortium, Auton A, Brooks LD, et al. A global reference for human genetic variation. *Nature*. 2015;526(7571):68-74. doi:10.1038/nature15393

4.	Byrska-Bishop M, Evani US, Zhao X, et al. High-coverage whole-genome sequencing of the expanded 1000 Genomes Project cohort including 602 trios. *Cell*. 2022;185(18):3426-3440.e19. doi:10.1016/j.cell.2022.08.004

5.	Browning BL, Browning SR. Genotype Imputation with Millions of Reference Samples. *Am J Hum Genet*. 2016;98(1):116-126. doi:10.1016/j.ajhg.2015.11.020

6.	Peltonen L, Jalanko A, Varilo T. Molecular genetics of the Finnish disease heritage. *Hum Mol Genet*. 1999;8(10):1913-1923. doi:10.1093/hmg/8.10.1913

7.	Bycroft C, Freeman C, Petkova D, et al. The UK Biobank resource with deep phenotyping and genomic data. *Nature*. 2018;562(7726):203-209. doi:10.1038/s41586-018-0579-z

8.	Pedersen BS, Quinlan AR. Who’s Who? Detecting and Resolving Sample Anomalies in Human DNA Sequencing Studies with Peddy. *Am J Hum Genet*. 2017;100(3):406-413. doi:10.1016/j.ajhg.2017.01.017

9.	Sun BB, Chiou J, Traylor M, et al. Genetic regulation of the human plasma proteome in 54,306 UK Biobank participants. Published online June 18, 2022:2022.06.17.496443. doi:10.1101/2022.06.17.496443

10.	Sun BB, Maranville JC, Peters JE, et al. Genomic atlas of the human plasma proteome. *Nature*. 2018;558(7708):73-79. doi:10.1038/s41586-018-0175-2

11.	Mbatchou J, Barnard L, Backman J, et al. Computationally efficient whole-genome regression for quantitative and binary traits. *Nat Genet*. 2021;53(7):1097-1103. doi:10.1038/s41588-021-00870-7

12.	Ritchie SC, Lambert SA, Arnold M, et al. Integrative analysis of the plasma proteome and polygenic risk of cardiometabolic diseases. *Nat Metab*. 2021;3(11):1476-1483. doi:10.1038/s42255-021-00478-5

13.	Giambartolomei C, Zhenli Liu J, Zhang W, et al. A Bayesian framework for multiple trait colocalization from summary association statistics. *Bioinforma Oxf Engl*. 2018;34(15):2538-2545. doi:10.1093/bioinformatics/bty147

14.	Guo H, Fortune MD, Burren OS, Schofield E, Todd JA, Wallace C. Integration of disease association and eQTL data using a Bayesian colocalisation approach highlights six candidate causal genes in immune-mediated diseases. *Hum Mol Genet*. 2015;24(12):3305-3313. doi:10.1093/hmg/ddv077

15.	Wallace C. A more accurate method for colocalisation analysis allowing for multiple causal variants. *PLOS Genet*. 2021;17(9):e1009440. doi:10.1371/journal.pgen.1009440

16.	Purcell S, Neale B, Todd-Brown K, et al. PLINK: A Tool Set for Whole-Genome Association and Population-Based Linkage Analyses. *Am J Hum Genet*. 2007;81(3):559-575.

17.	Segar MW, Patel KV, Hellkamp AS, et al. Validation of the WATCH‐DM and TRS‐HFDM Risk Scores to Predict the Risk of Incident Hospitalization for Heart Failure Among Adults With Type 2 Diabetes: A Multicohort Analysis. *J Am Heart Assoc*. 2022;11(11):e024094. doi:10.1161/JAHA.121.024094

18.	Brady W, de Souza K. The HEART score: A guide to its application in the emergency department. *Turk J Emerg Med*. 2018;18(2):47-51. doi:10.1016/j.tjem.2018.04.004

19.	Moura FA, Berg DD, Bellavia A, et al. Risk Assessment of Kidney Disease Progression and Efficacy of SGLT2 Inhibition in Patients With Type 2 Diabetes. *Diabetes Care*. 2023;46(10):1807-1815. doi:10.2337/dc23-0492

20.	Pilla SJ, Yeh HC, Juraschek SP, Clark JM, Maruthur NM. Predictors of Insulin Initiation in Patients with Type 2 Diabetes: An Analysis of the Look AHEAD Randomized Trial. *J Gen Intern Med*. 2018;33(6):839-846. doi:10.1007/s11606-017-4282-9

21.	Obradovic M, Sudar-Milovanovic E, Soskic S, et al. Leptin and Obesity: Role and Clinical Implication. *Front Endocrinol*. 2021;12:585887. doi:10.3389/fendo.2021.585887

22.	Eguchi J, Wada J, Hida K, et al. Identification of adipocyte adhesion molecule (ACAM), a novel CTX gene family, implicated in adipocyte maturation and development of obesity. *Biochem J*. 2005;387(Pt 2):343-353. doi:10.1042/BJ20041709

23.	Martin AR, Kanai M, Kamatani Y, Okada Y, Neale BM, Daly MJ. Clinical use of current polygenic risk scores may exacerbate health disparities. *Nat Genet*. 2019;51(4):584-591. doi:10.1038/s41588-019-0379-x

24.	Ruan Y, Lin YF, Feng YCA, et al. Improving Polygenic Prediction in Ancestrally Diverse Populations. *Nat Genet*. 2022;54(5):573-580. doi:10.1038/s41588-022-01054-7

### **Supplementary Figures**

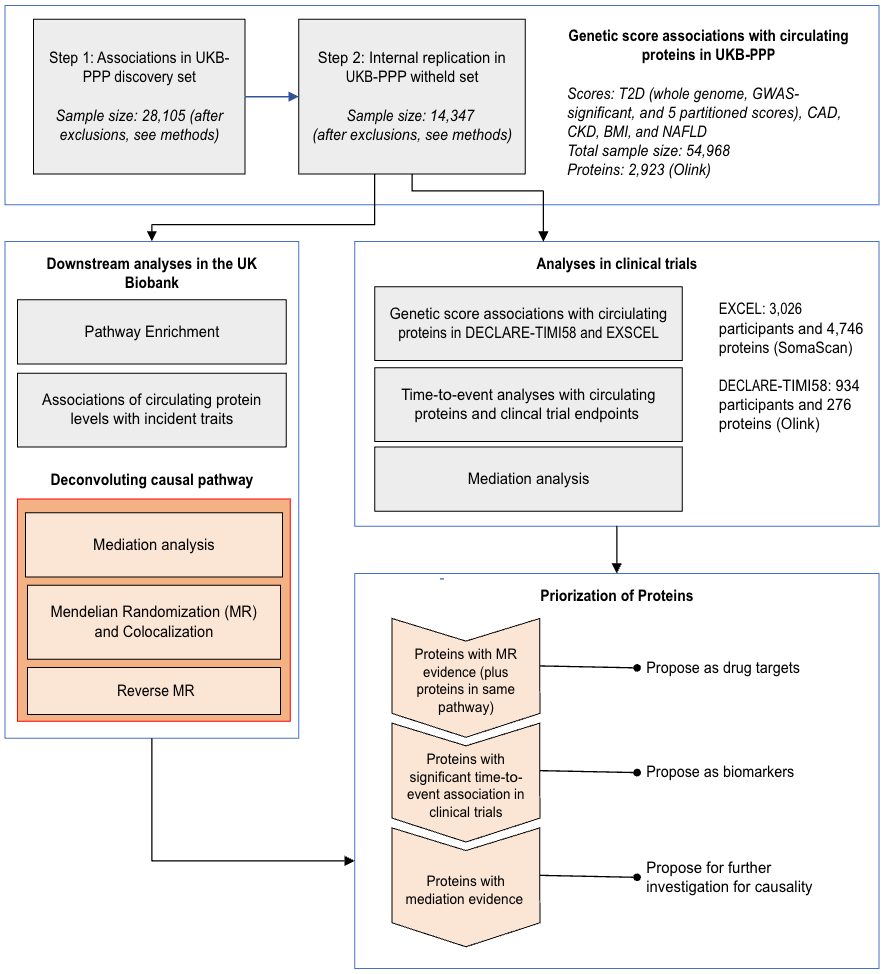

Supplementary Figure 1: Flowchart demonstrating study design.

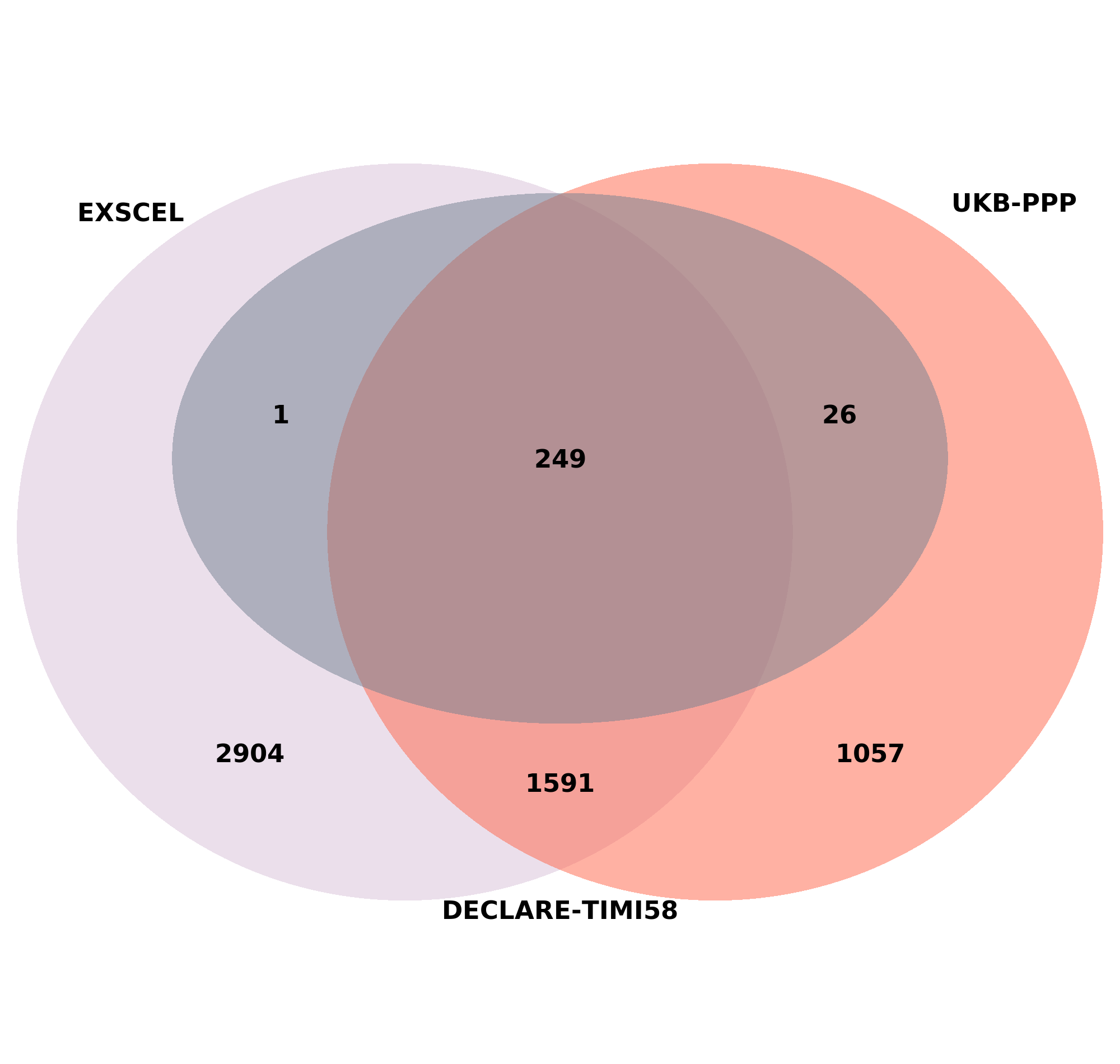

**B.**

**A.**

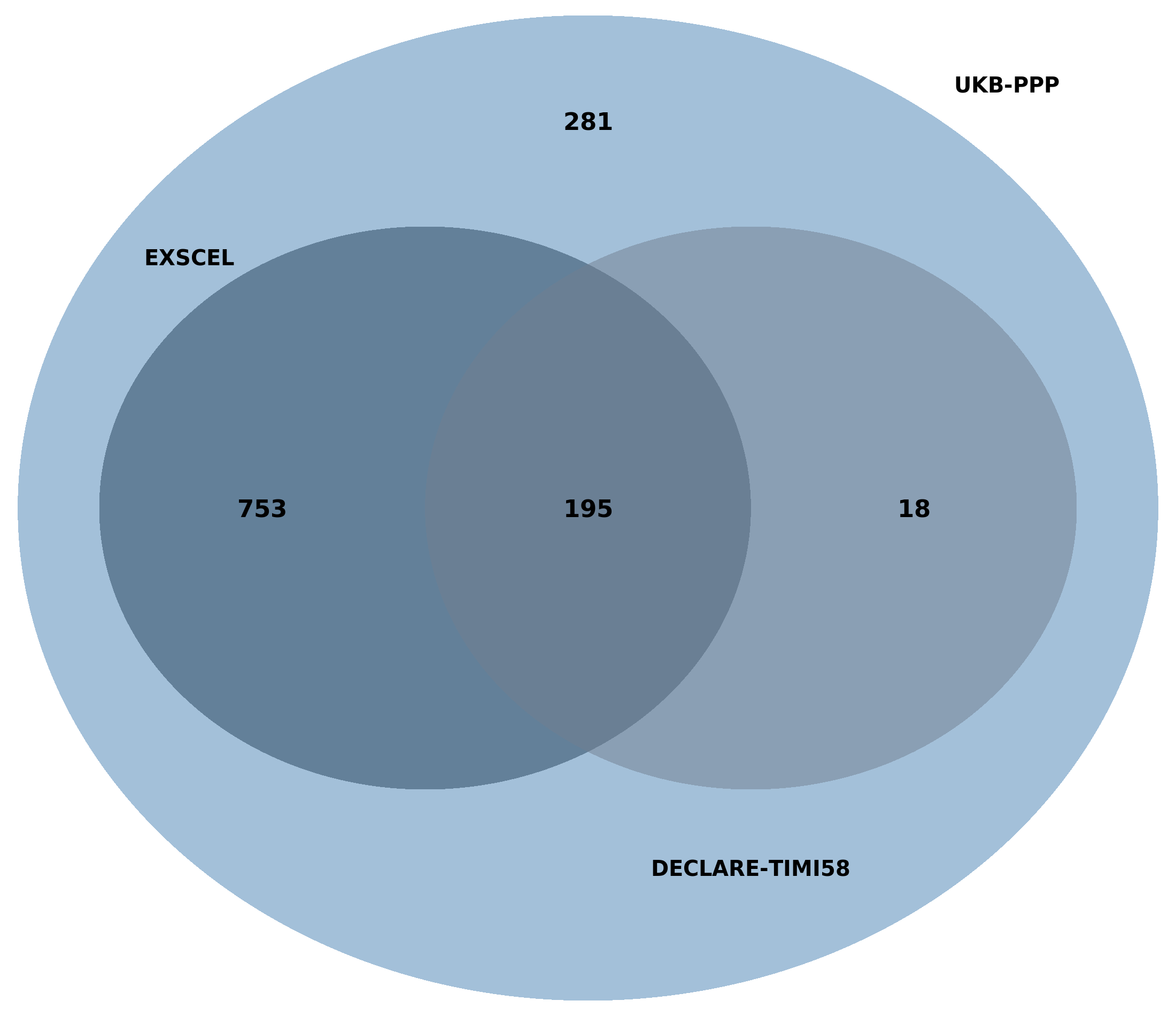

**Supplementary Figure 2:** Proteomics overlap. A: Venn diagram of protein intersections across UKB, EXSCEL, and DECLARE-TIMI58. The purple circle corresponds to EXSCEL, the red circle corresponds to UKB-PPP, and the grey circle (completely internal to the other sets) corresponds to DECLARE-TIMI58. B: Venn diagram of PGS-significant protein intersections across UKB, EXSCEL, and DECLARE-TIMI58. Note that restricts proteins to only those available in UKB. The large blue circle are PGS-significant proteins in UKB, the dark blue circle within the UKB circle corresponds to EXSCEL, and the grey circle to the right corresponds to DECLARE.

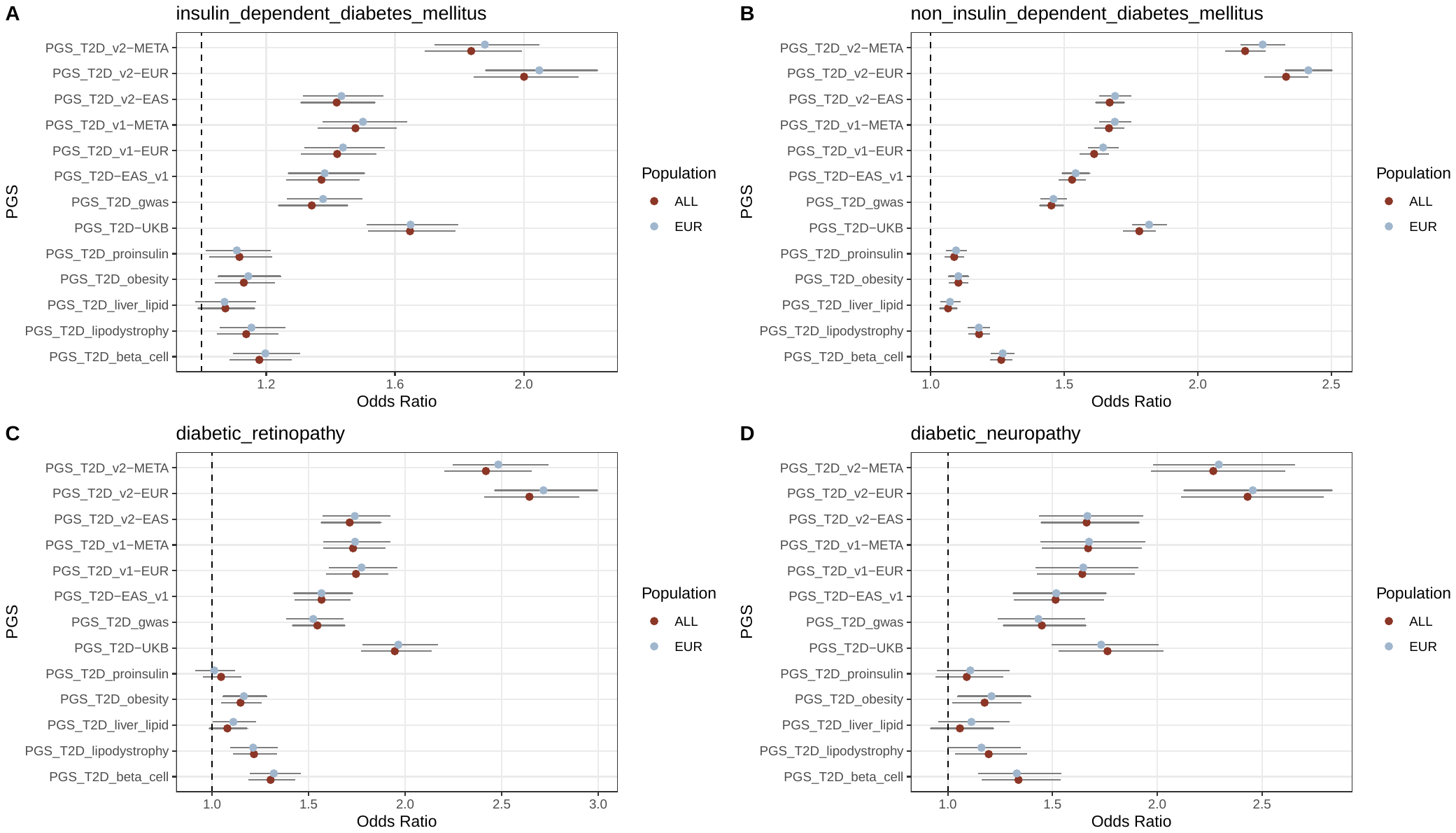

**Supplementary Figure 3: T2D PGS associations with T2D-related traits in UKB-PPP participants.** Various T2D PGS (including the partitioned T2D polygenic scores) were tested for association with insulin-dependent diabetes, non-insulin dependent diabetes, diabetic retinopathy, and diabetic retinopathy using logistic regression. The best performing genome-wide PGS by effect size (odds ratio) for non-insulin dependent diabetes was selected for the study and referred to as the PGS_T2D_gw_ in the text (T2D_PGS_v2-EUR in this figure). Population labels were either all (red) or European-only (blue) inferred using KING.

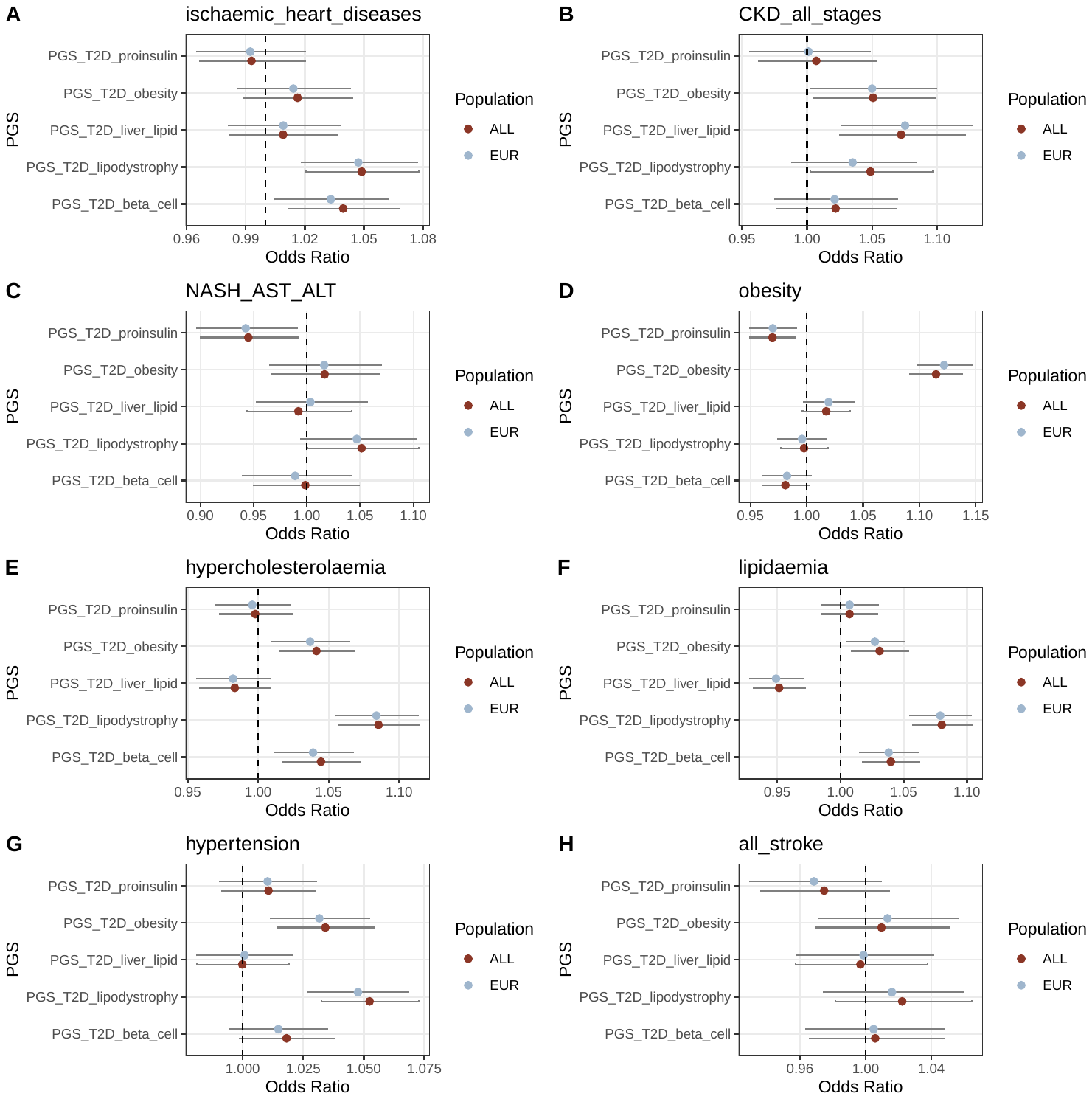

**Supplementary Figure 4: T2D partitioned polygenic score (pPS) associations with selected traits in UKB-PPP participants.** The T2D pPS were tested for association with select cardiometabolic traits in the UKB-PPP participants. Population labels were either all (red) or European-only (blue) inferred using KING. Note that figure F refers to the ICD10 code E78: disorders of lipoprotein metabolism and other lipidaemias.

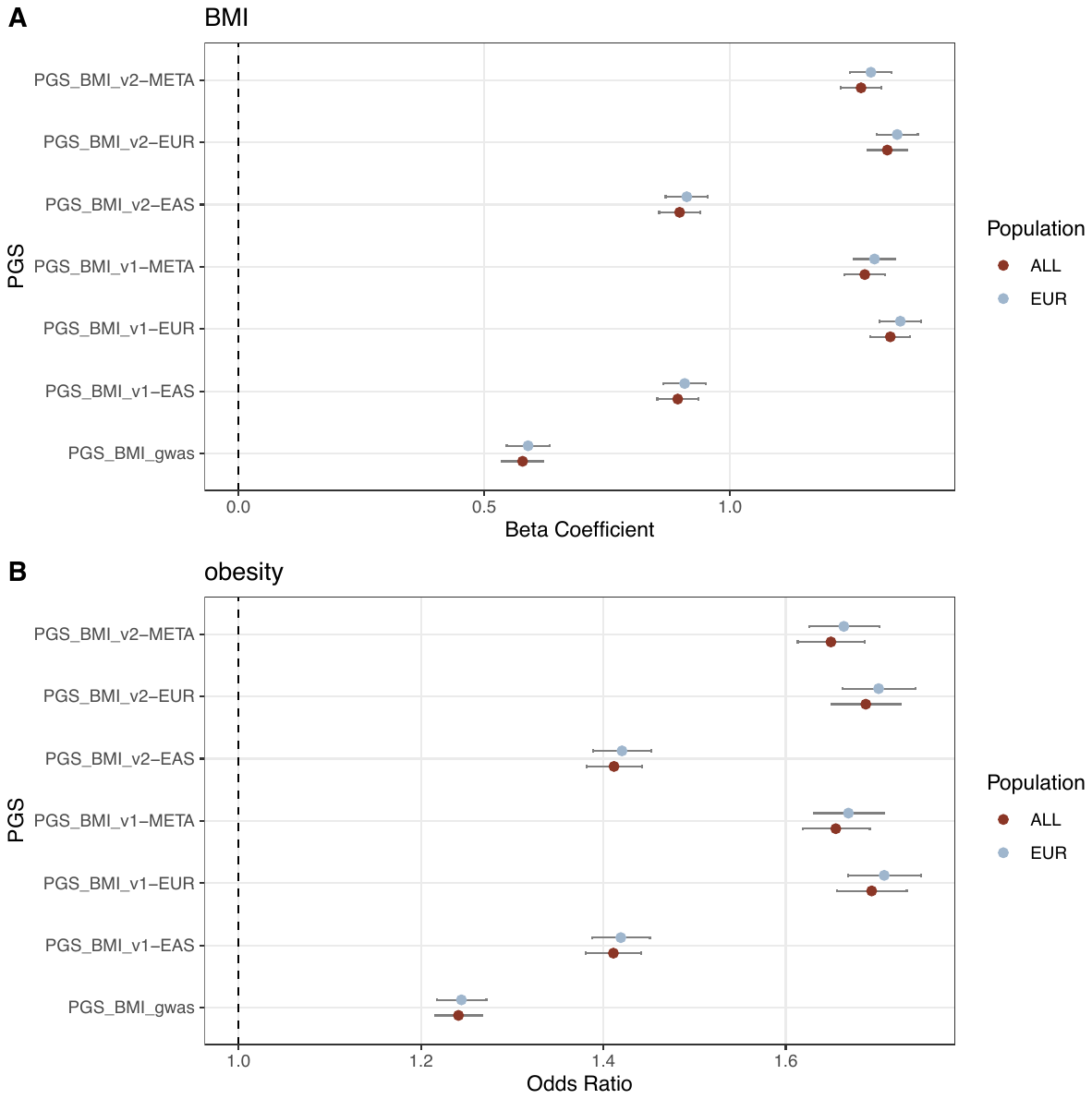

**Supplementary Figure 5: BMI PGS associations with BMI-related traits in UKB-PPP participants.** BMI PGS were tested for association with BMI and obesity (> 30 BMI). The best performing PGS by effect size (beta coefficient) for BMI was selected for the study and referred to as the PGS_BMI_ in the text (here labelled BMI_PGS_v1-EUR). Population labels were either all (red) or European-only (blue) inferred using KING.

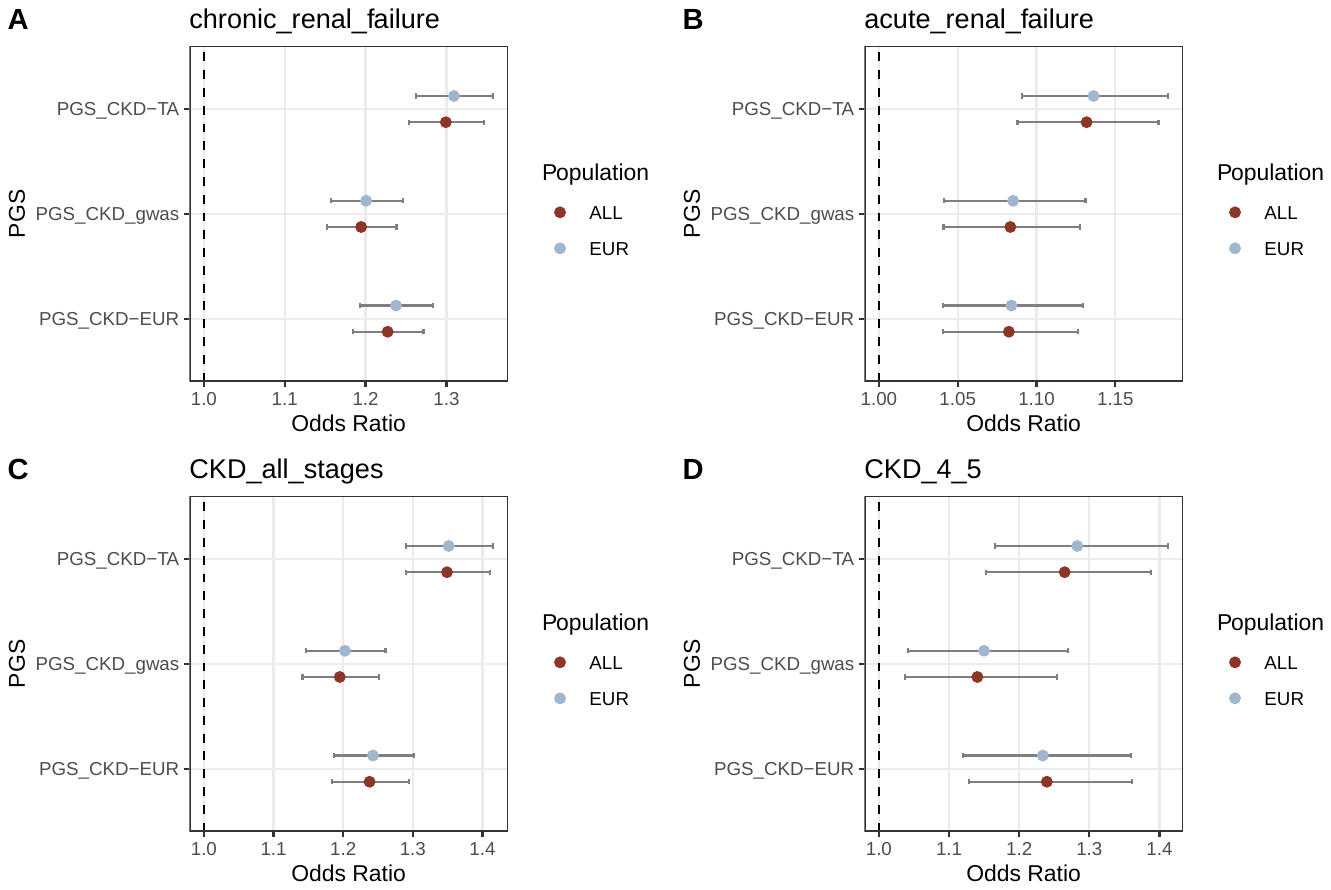

**Supplementary Figure 6: CKD PGS associations with renal-related traits in UKB-PPP participants.** CKD PGS were tested for association with chronic renal failure, acute renal failure, CKD (all stages), and CKD (stages 4 and 5 only). The best performing PGS by odds ratio for CKD (all stages) was selected for the study and referred to as the PGS_CKD_ in the text (labelled as PGS_CKD-TA in this figure). Population labels were either all (red) or European-only (blue) inferred using KING.

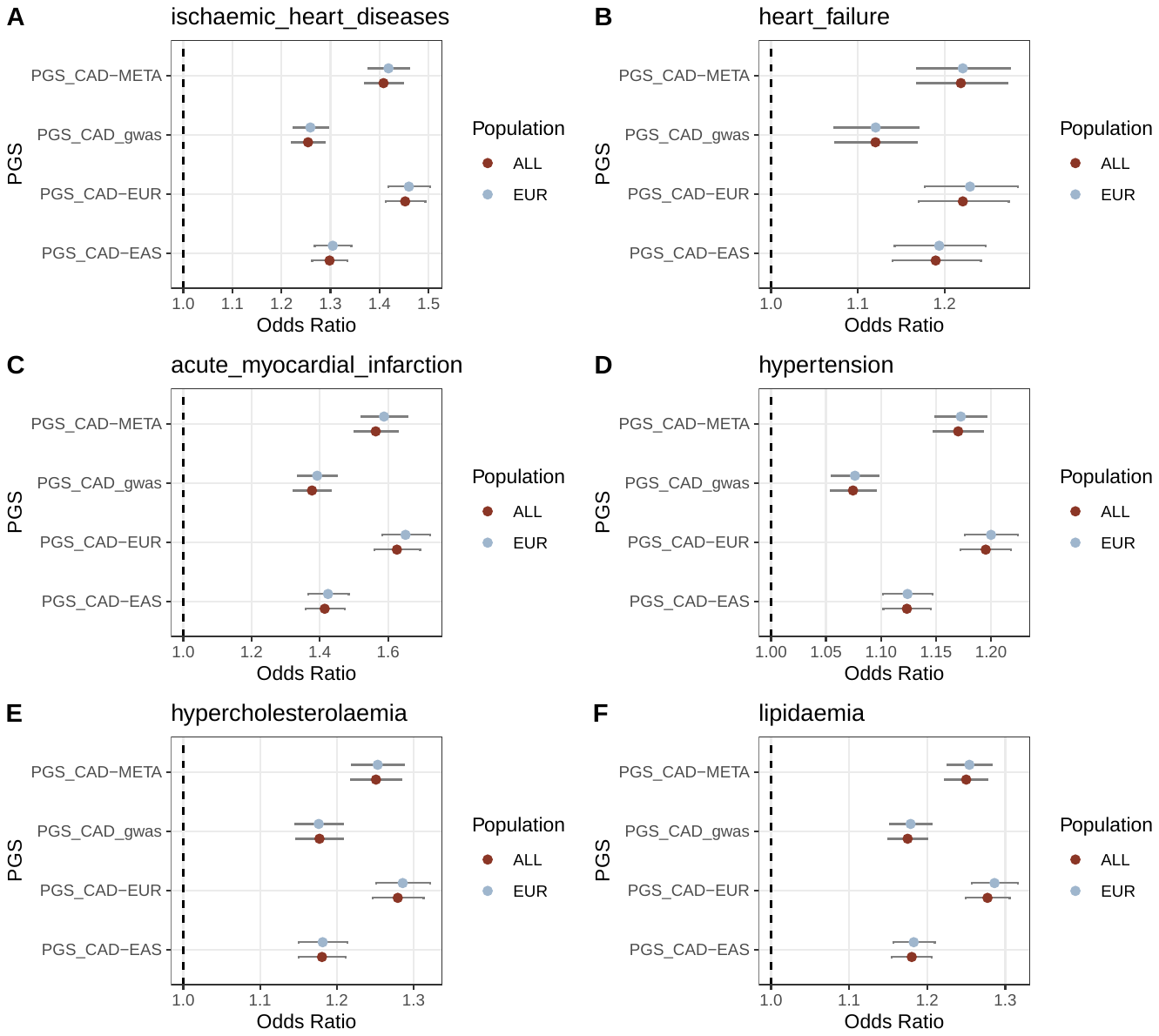

**Supplementary Figure 7: CAD PGS associations with cardiovascular-related traits in UKB-PPP participants.** CAD PGS were tested for association with ischaemic heart disease, heart failure, acute myocardial infarction, hypertension, hypercholesteremia, and lipidaemia to ensure robustness in the score. The best performing PGS by effect size (odds ratio) for ischaemic heart disease was selected for the study and referred to as the PGS_CAD_ in the text (PGS_CAD-EUR in this figure). Population labels were either all (red) or European-only (blue) inferred using KING.

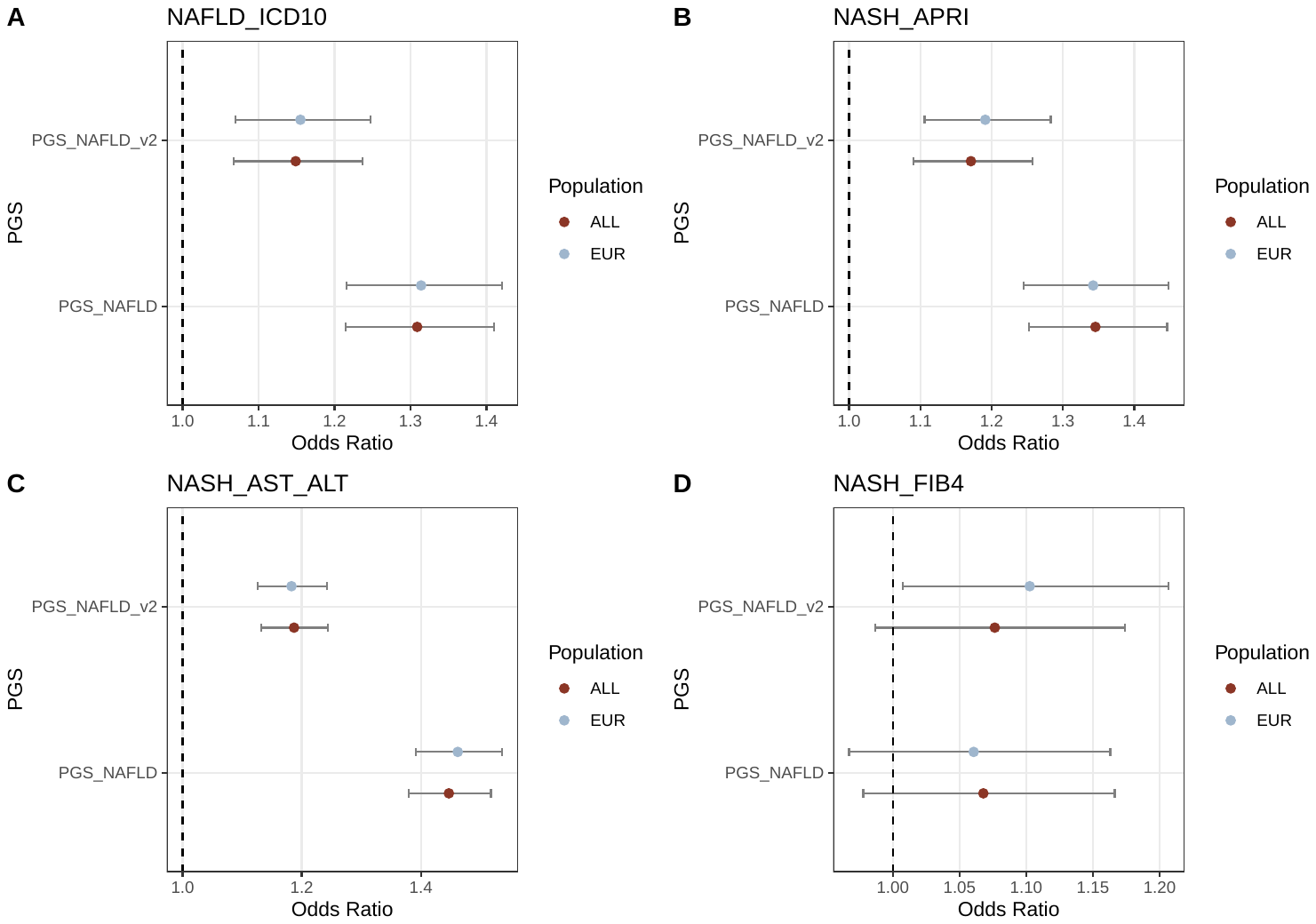

**Supplementary Figure 8: NAFLD (NASH) PGS associations with liver-related traits in UKB-PPP participants.** Two NAFLD PGS (both using only GWAS-significant variants) were tested for association with NAFLD from ICD10 codes and three biomarker-based definitions. The best performing PGS by odds ratio for NAFLD (using AST and ALT) was selected for the study and referred to as the PGS_NAFLD_ in the text (PGS_NAFLD in this figure). Population labels were either all (red) or European-only (blue) inferred using KING.

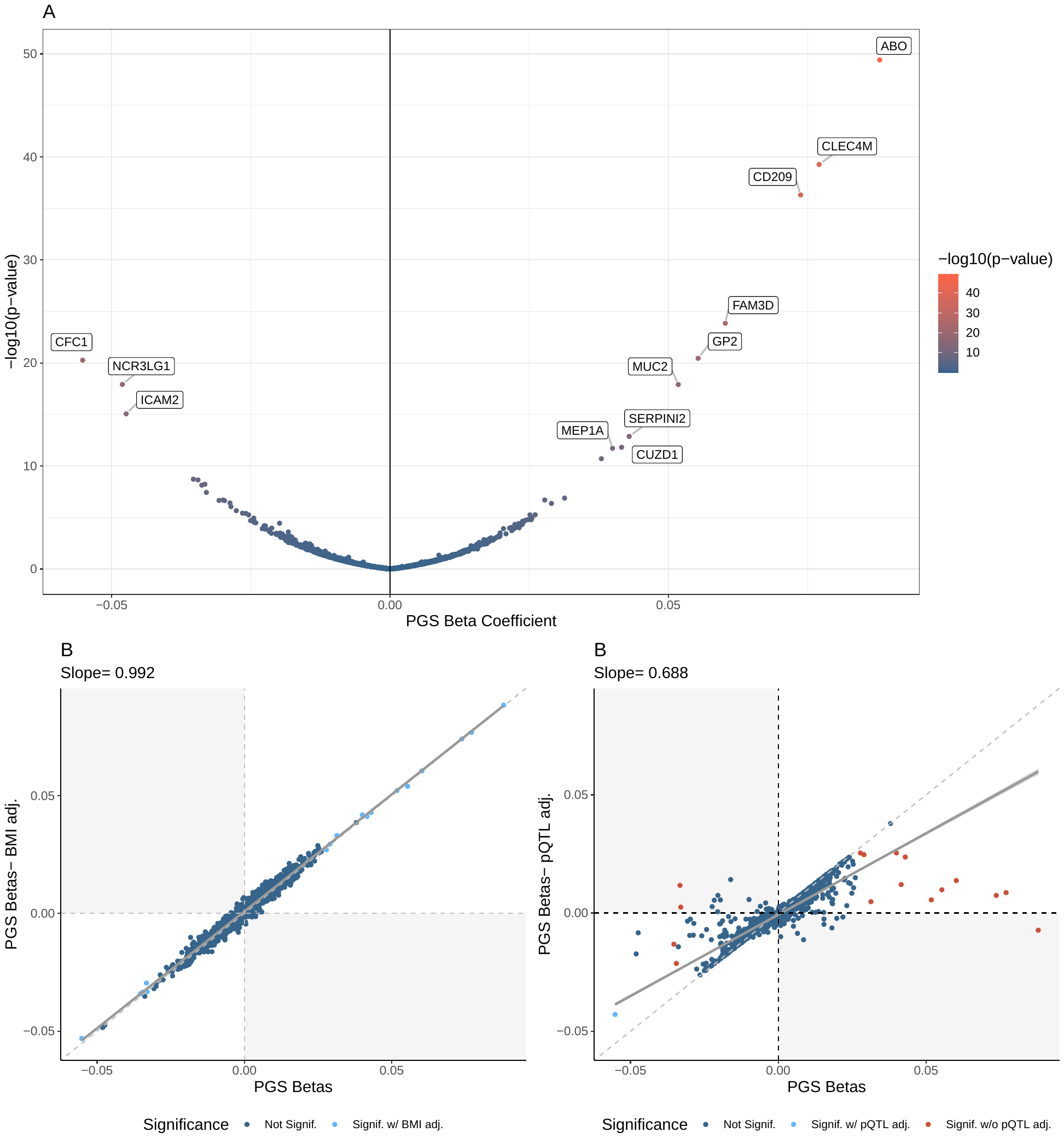

**Supplementary Figure 9:** A: Volcano plot of PGS_T2D_beta_cell_ associations. Labelled proteins were the top 0.5% of PGS_T2D_beta_cell_ associations by R^2^. B: Beta-beta plot of pPS-protein associations with (Y-axis) and without BMI-adjustment (X-axis). The regression line is solid grey while the diagonal is dashed grey. C. Beta-beta plot of pPS-protein associations with (Y-axis) and without pQTL-adjustment (X-axis). The regression line is solid grey while the diagonal is dashed grey. See Figure 1 for a description of the colouring schemes for panels A-C.

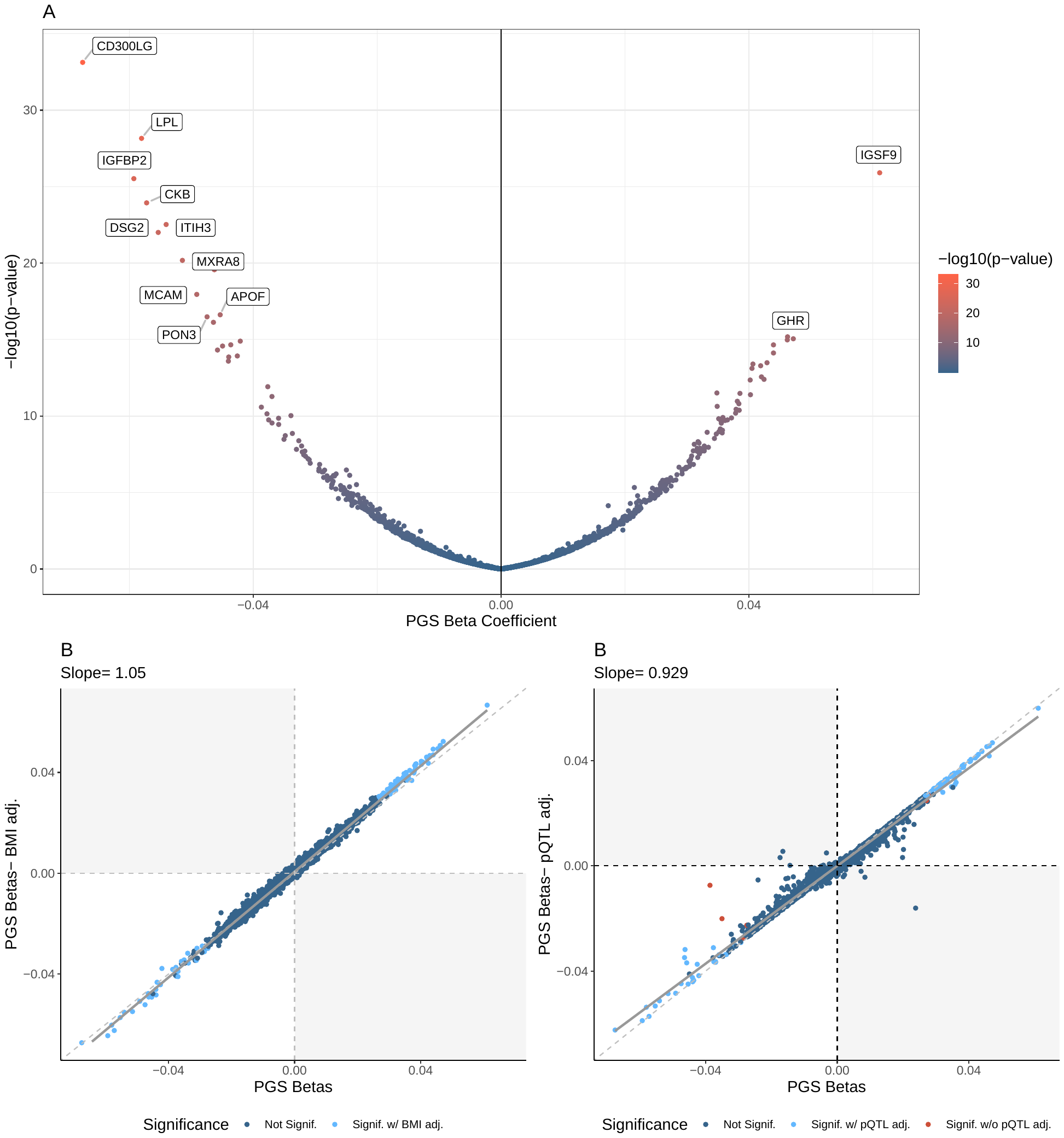

**Supplementary Figure 10:** A: Volcano plot of PGS_T2D_lipodystrophy_ associations. Labelled proteins were the top 0.5% of PGS_T2D_lipodystrophy_ associations by R^2^. B: Beta-beta plot of pPS-protein associations with (Y-axis) and without BMI-adjustment (X-axis). The regression line is solid grey while the diagonal is dashed grey. C. Beta-beta plot of pPS-protein associations with (Y-axis) and without pQTL-adjustment (X-axis). The regression line is solid grey while the diagonal is dashed grey. See Figure 1 for a description of the colouring schemes for panels A-C.

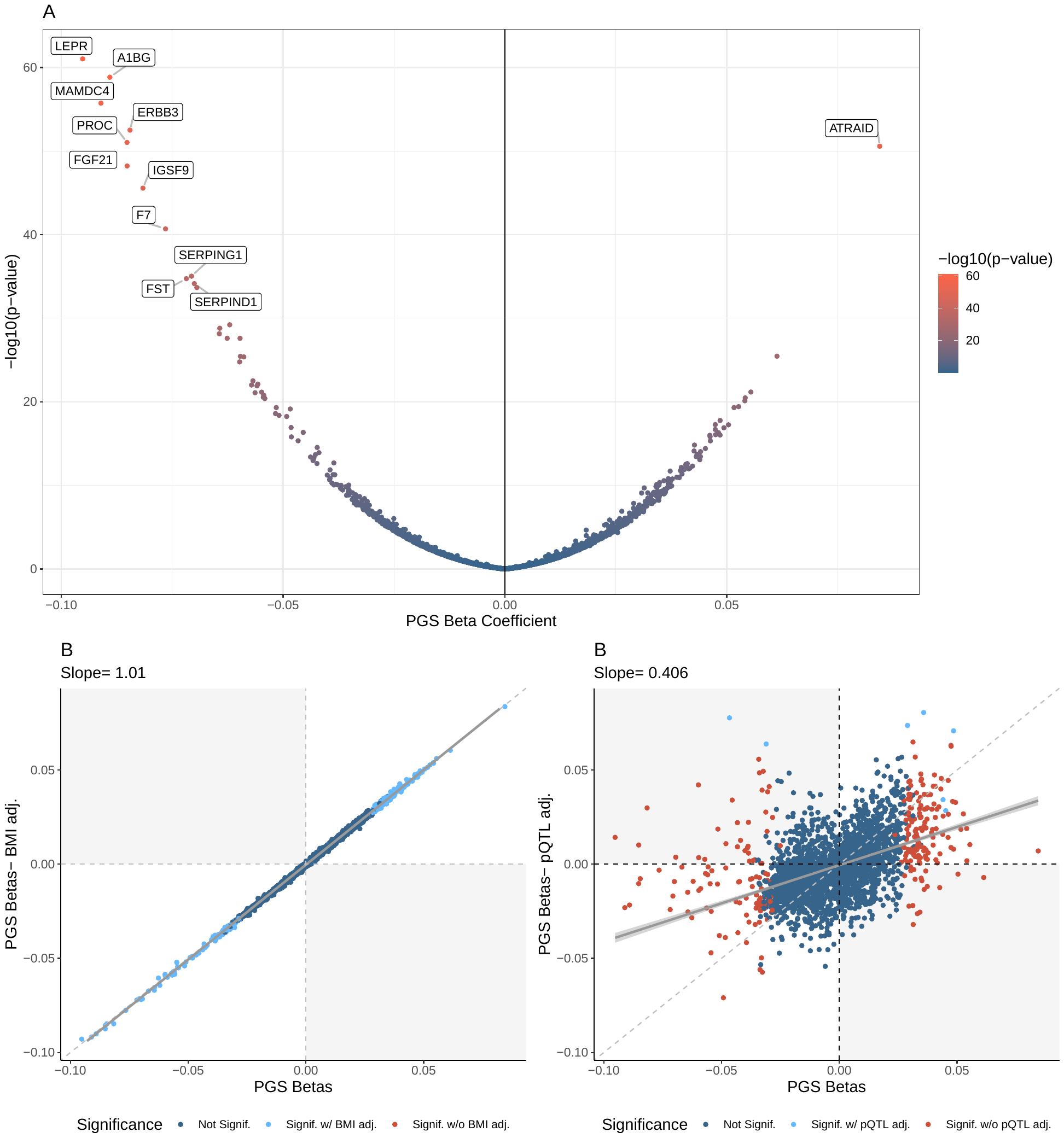

**Supplementary Figure 11:** A: Volcano plot of PGS_T2D_liver_lipid_ associations. Labelled proteins were the top 0.5% of PGS_T2D_liver_lipid_ associations by R^2^. B: Beta-beta plot of pPS-protein associations with (Y-axis) and without BMI-adjustment (X-axis). The regression line is solid grey while the diagonal is dashed grey. C. Beta-beta plot of pPS-protein associations with (Y-axis) and without pQTL-adjustment (X-axis). The regression line is solid grey while the diagonal is dashed grey. See Figure 1 for a description of the colouring schemes for panels A-C.

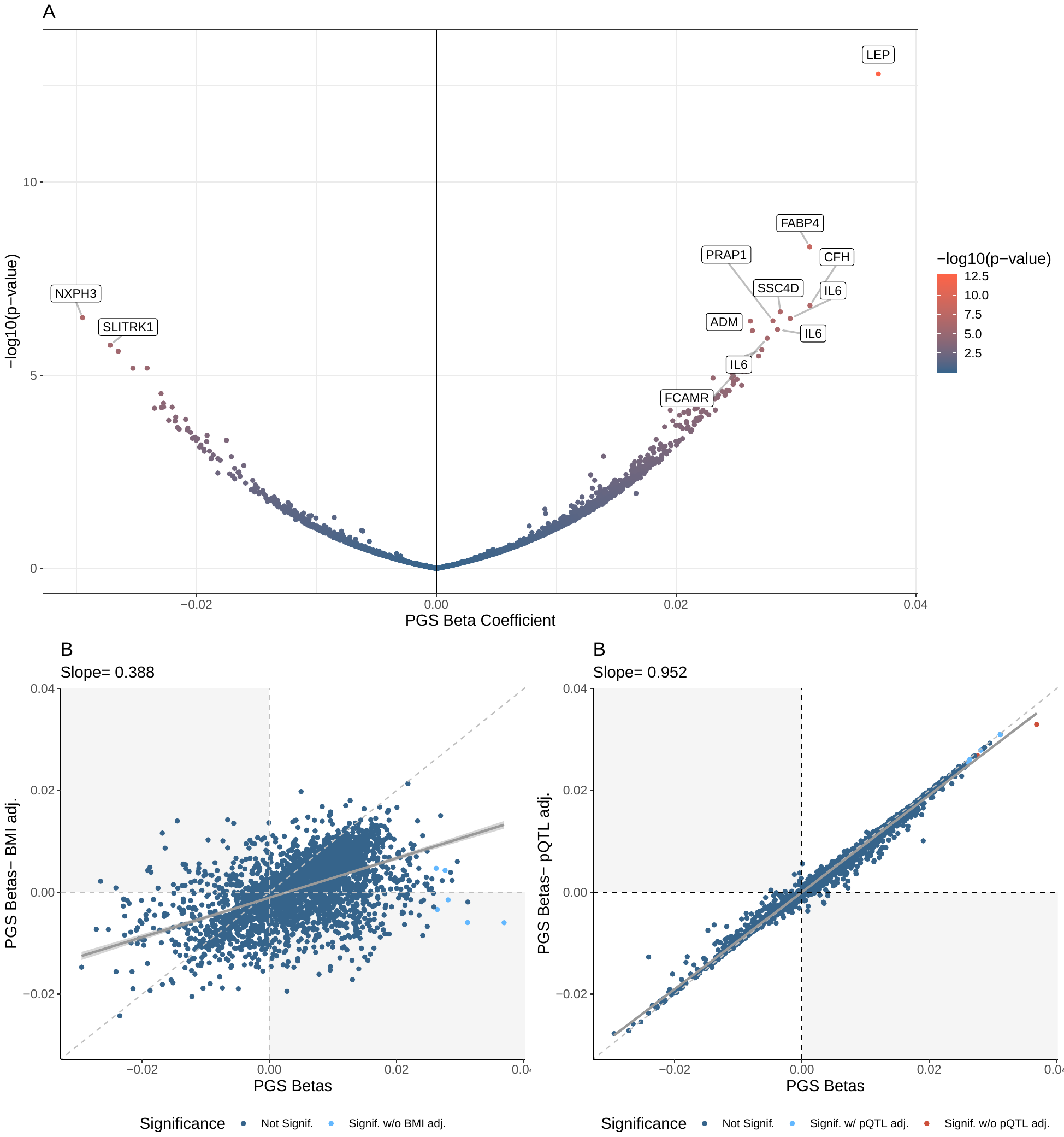

**Supplementary Figure 12:** A: Volcano plot of PGS_T2D_obesity_ associations. Labelled proteins were the top 0.5% of PGS_T2D_obesity_ associations by R^2^. B: Beta-beta plot of pPS-protein associations with (Y-axis) and without BMI-adjustment (X-axis). The regression line is solid grey while the diagonal is dashed grey. C. Beta-beta plot of pPS-protein associations with (Y-axis) and without pQTL-adjustment (X-axis). The regression line is solid grey while the diagonal is dashed grey. See Figure 1 for a description of the colouring schemes for panels A-C.

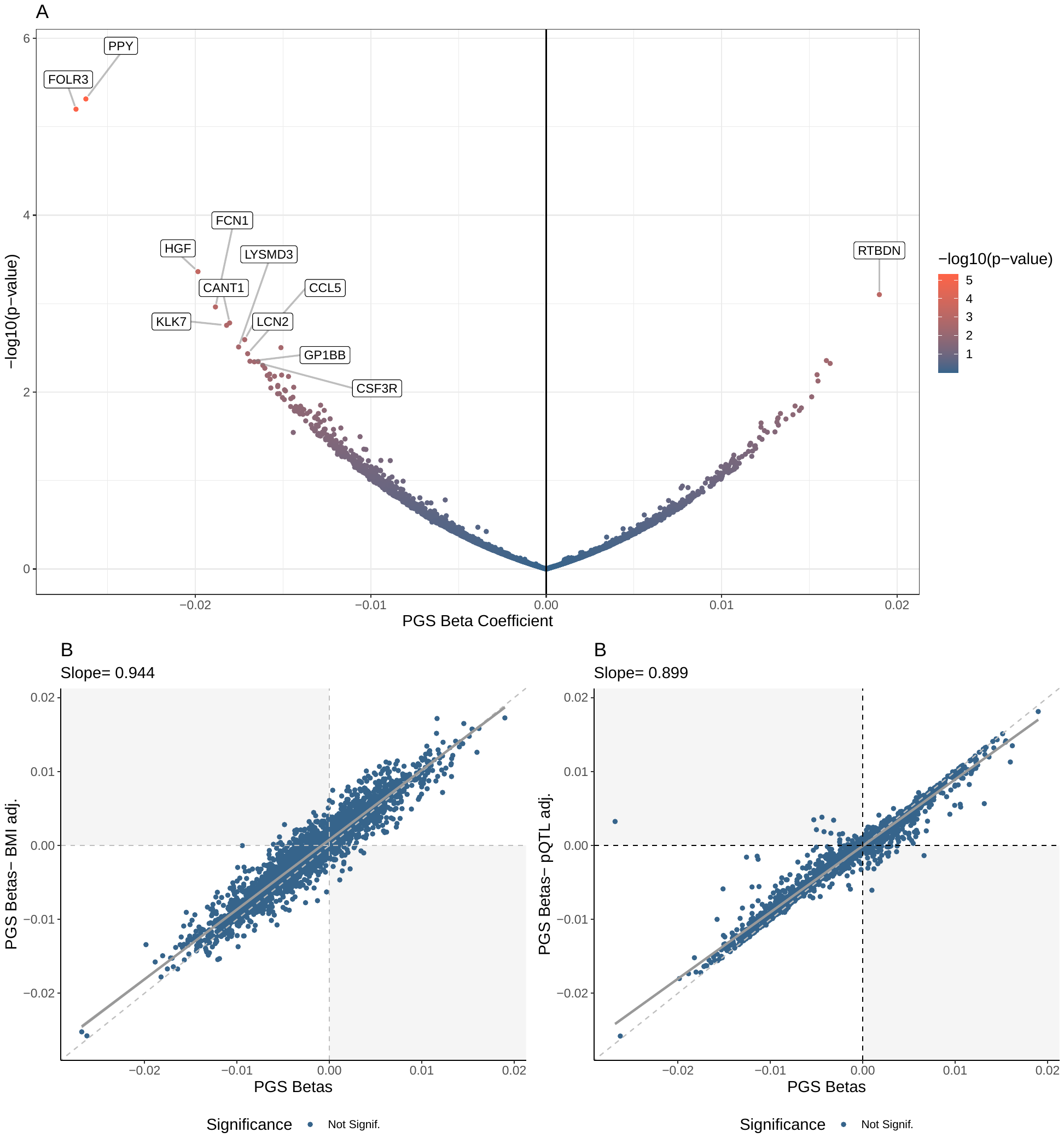

**Supplementary Figure 13:** A: Volcano plot of PGS_T2D_proinsulin_ associations. Labelled proteins were the top 0.5% of PGS_T2D_proinsulin_ associations by R^2^. B: Beta-beta plot of pPS-protein associations with (Y-axis) and without BMI-adjustment (X-axis). The regression line is solid grey while the diagonal is dashed grey. C. Beta-beta plot of pPS-protein associations with (Y-axis) and without pQTL-adjustment (X-axis). The regression line is solid grey while the diagonal is dashed grey. See Figure 1 for a description of the colouring schemes for panels A-C.

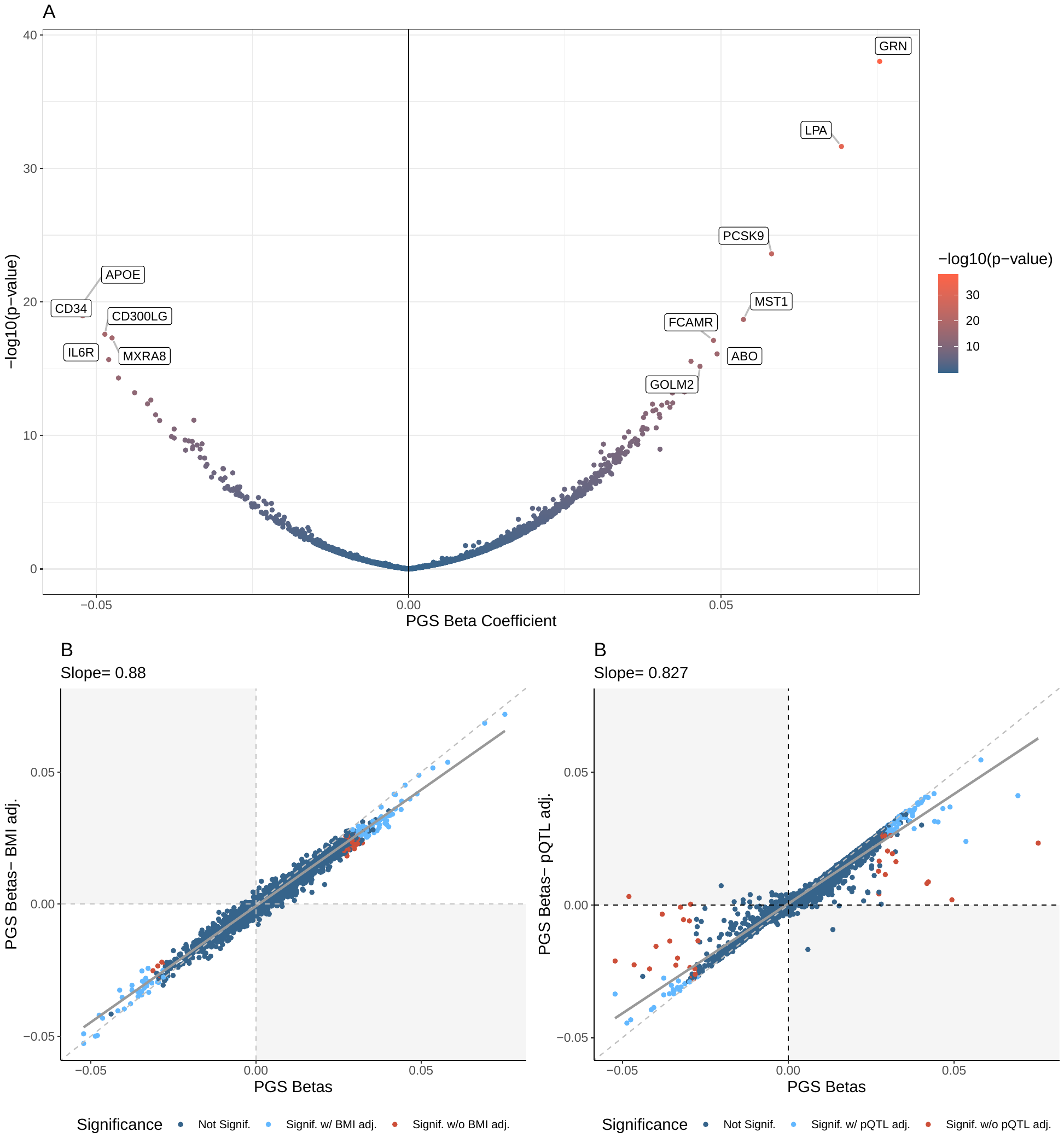

**Supplementary Figure 14: PGS_CAD_ plot.** A: Volcano plot of PGS_CAD_ associations. Labelled proteins were the top 0.5% of PGS associations by R^2^. B: Beta-beta plot of PGS-protein associations with (Y-axis) and without BMI-adjustment (X-axis). The regression line is solid grey while the diagonal is dashed grey. C. Beta-beta plot of PGS-protein associations with (Y-axis) and without pQTL-adjustment (X-axis). The regression line is solid grey while the diagonal is dashed grey. See Figure 1 for a description of the colouring schemes for panels A-C.

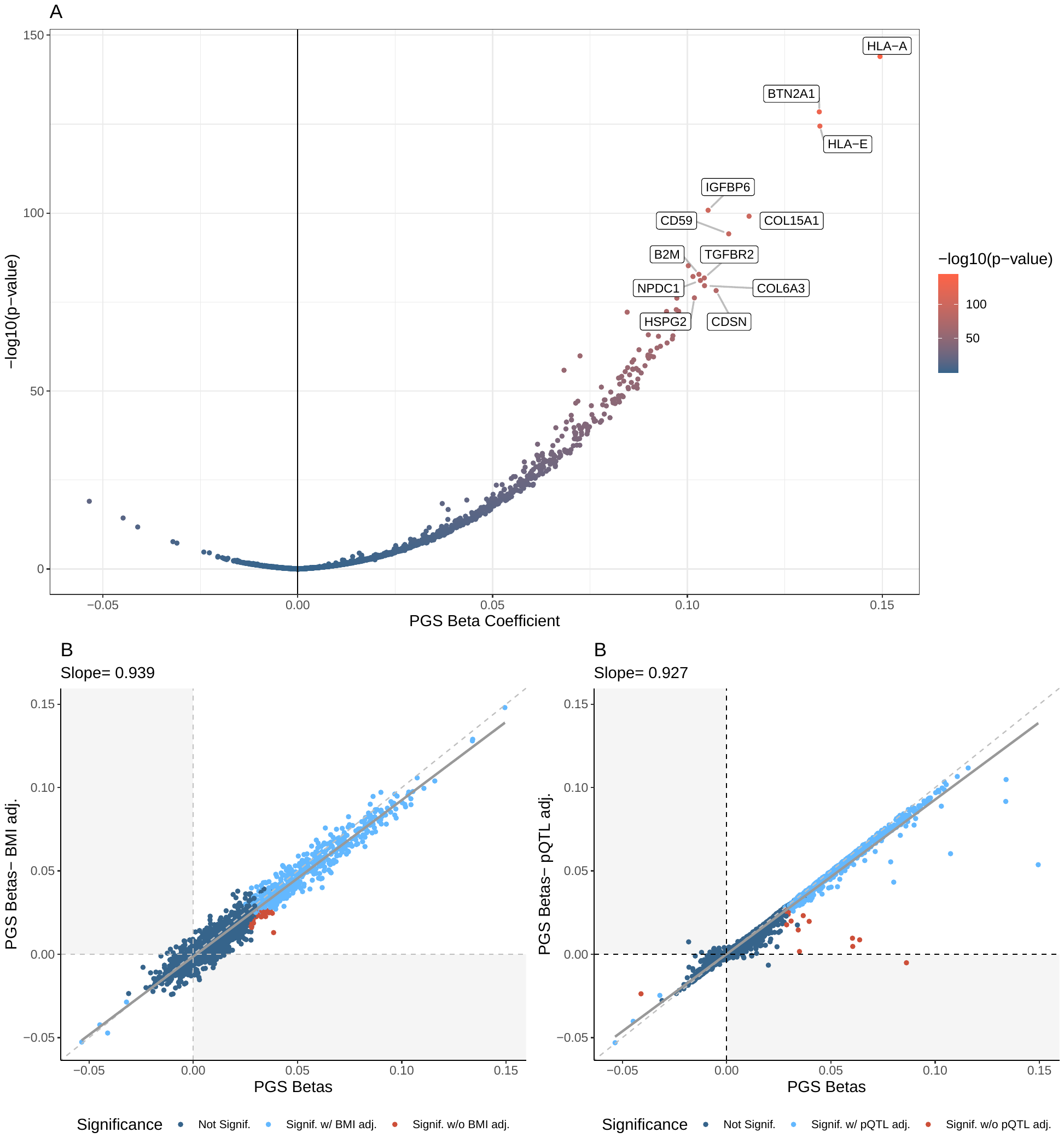

**Supplementary Figure 15: PGS_CKD_ plot.** A: Volcano plot of PGS_CKD_ associations. Labelled proteins were the top 0.5% of PGS associations by R^2^. B: Beta-beta plot of PGS-protein associations with (Y-axis) and without BMI-adjustment (X-axis). The regression line is solid grey while the diagonal is dashed grey. C. Beta-beta plot of PGS-protein associations with (Y-axis) and without pQTL-adjustment (X-axis). The regression line is solid grey while the diagonal is dashed grey. See Figure 1 for a description of the colouring schemes for panels A-C.

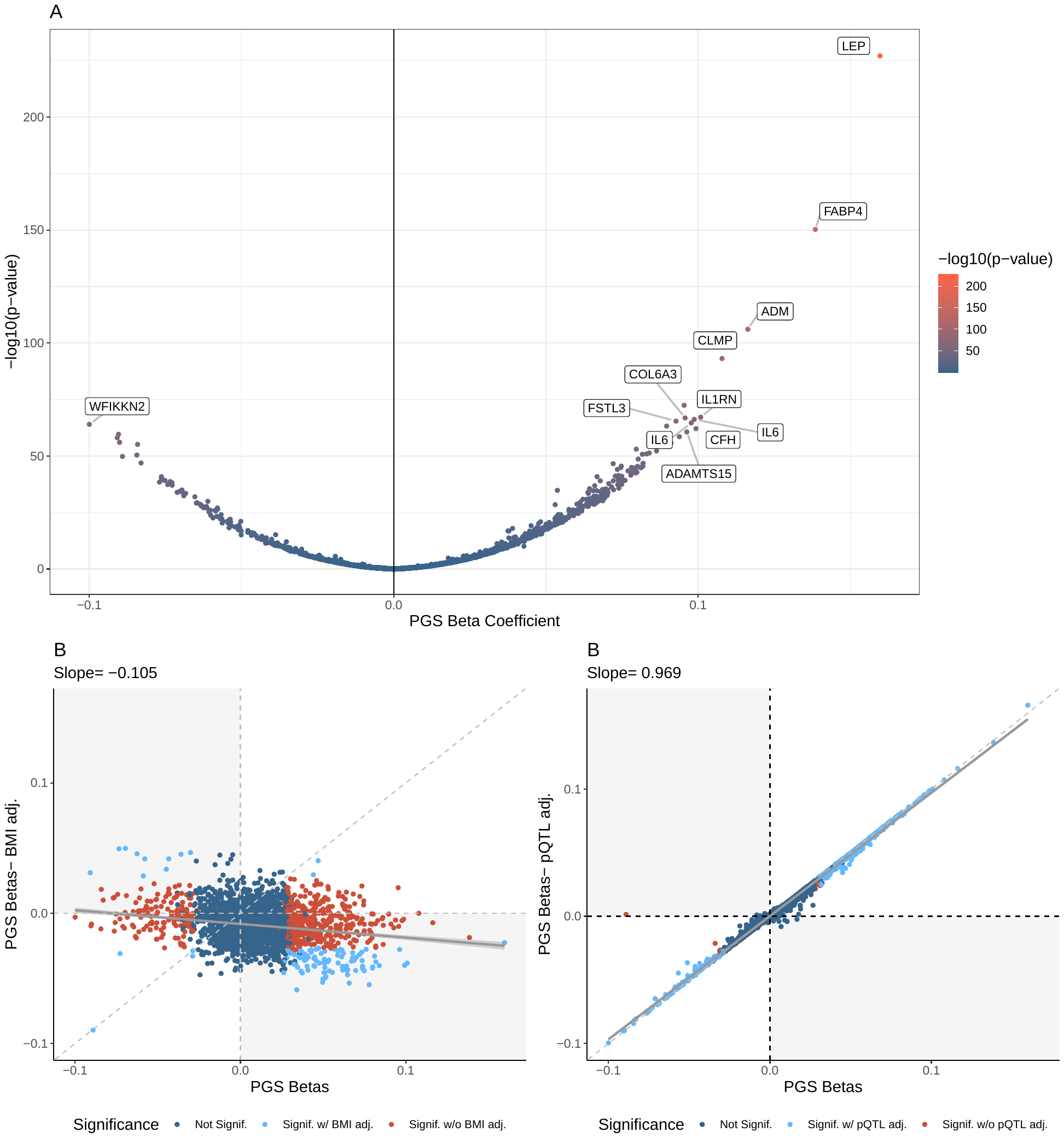

**Supplementary Figure 16: PGS_BMI_ plot.** A: Volcano plot of PGS_BMI_ associations. Labelled proteins were the top 0.5% of PGS associations by R^2^. B: Beta-beta plot of PGS-protein associations with (Y-axis) and without BMI-adjustment (X-axis). The regression line is solid grey while the diagonal is dashed grey. C. Beta-beta plot of PGS-protein associations with (Y-axis) and without pQTL-adjustment (X-axis). The regression line is solid grey while the diagonal is dashed grey. See Figure 1 for a description of the colouring schemes for panels A-C.

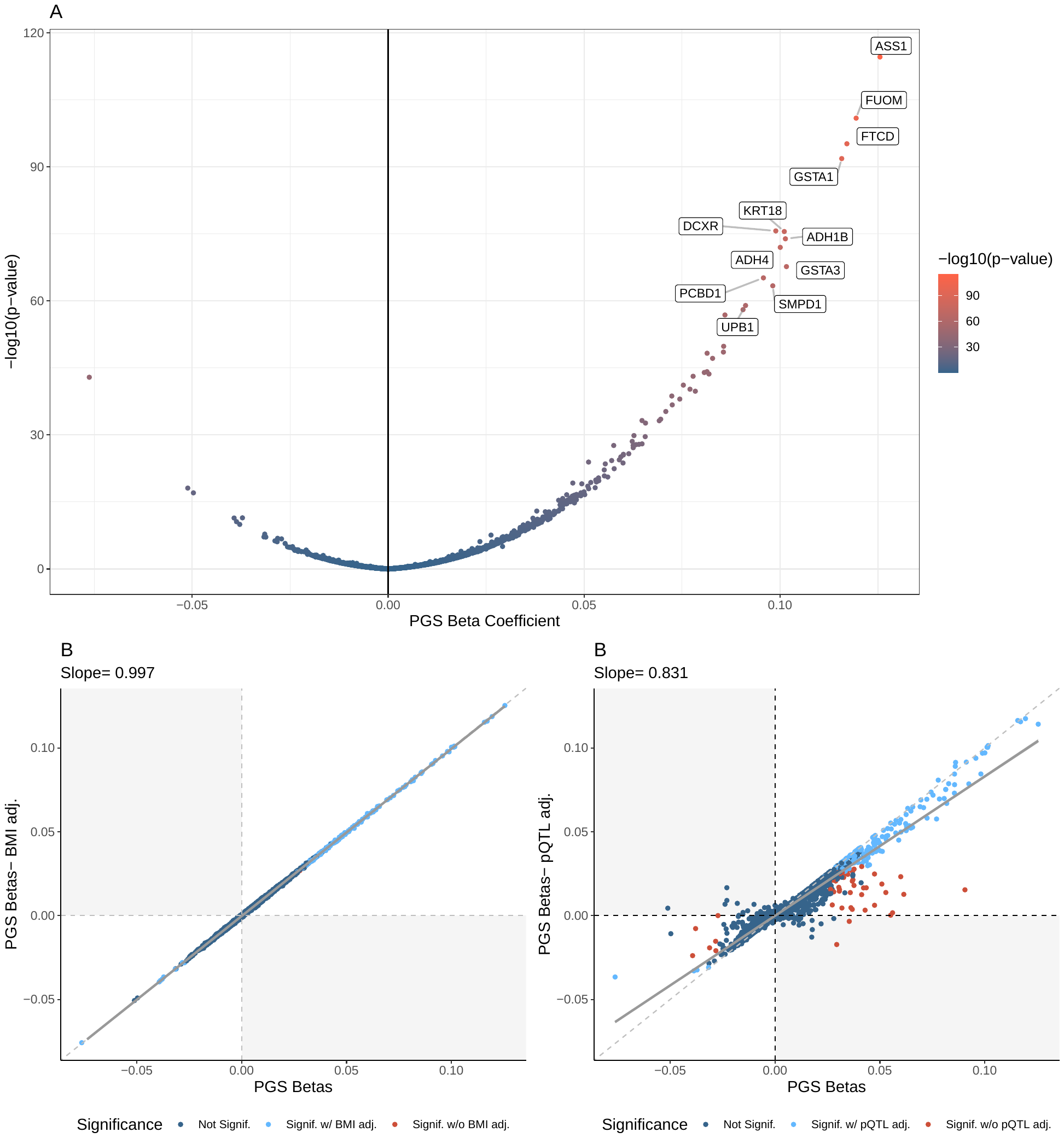

**Supplementary Figure 17: PGS_NAFLD_ plot.** A: Volcano plot of PGS_NAFLD_ associations. Labelled proteins were the top 0.5% of PGS associations by R^2^. B: Beta-beta plot of PGS-protein associations with (Y-axis) and without BMI-adjustment (X-axis). The regression line is solid grey while the diagonal is dashed grey. C. Beta-beta plot of PGS-protein associations with (Y-axis) and without pQTL-adjustment (X-axis). The regression line is solid grey while the diagonal is dashed grey. See Figure 1 for a description of the colouring schemes for panels A-C.

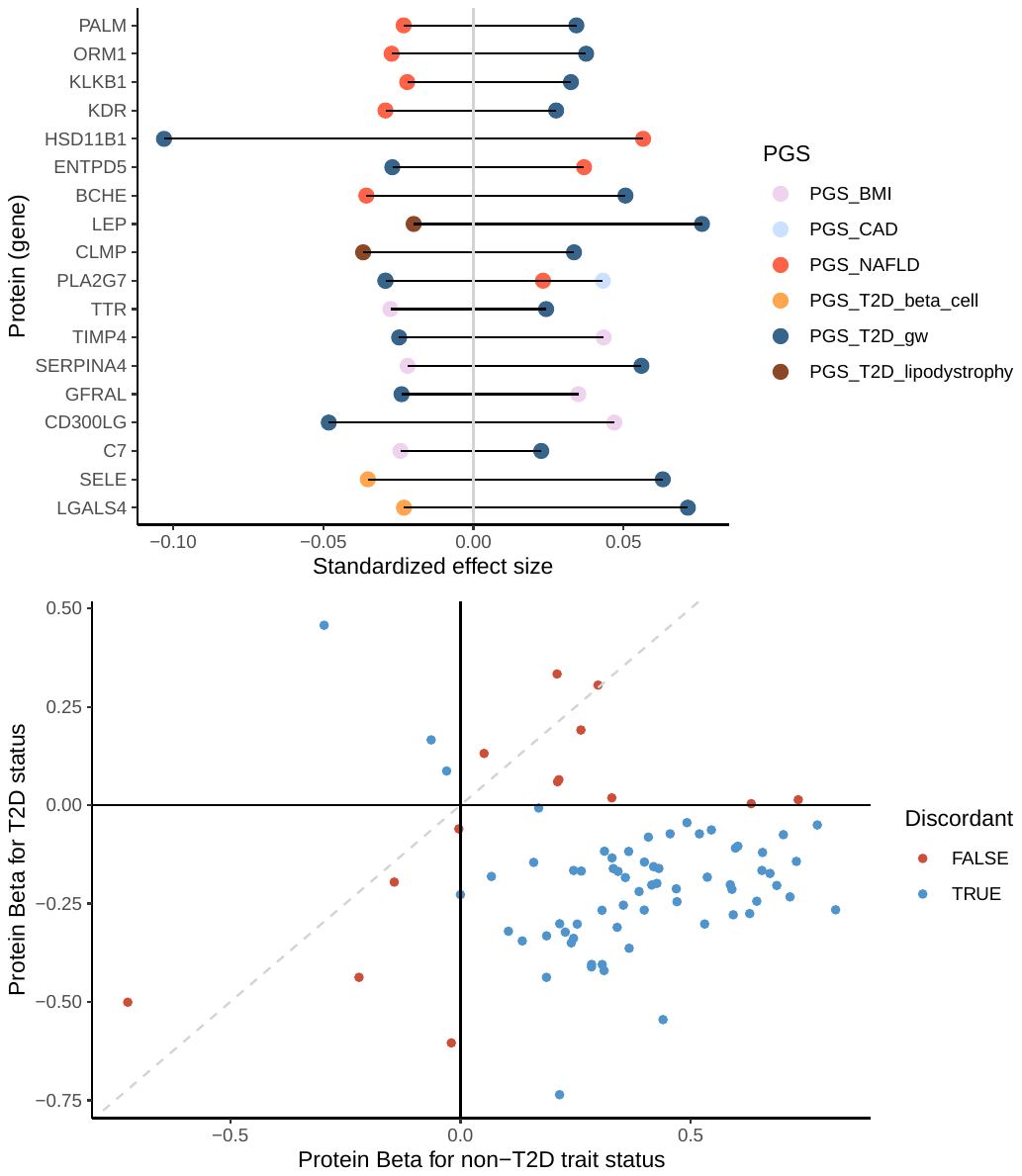

**Supplementary Figure 18: Discordant T2D PGS-protein associations.** Discordance was defined as proteins where the direction of effect of the T2D PGS and another score were opposite after a Bonferroni correction. Note that proteins discordant between the T2D PGS and CKD and liver lipid pPS are not shown due to the number of implicated proteins. B. Beta-beta plot of protein associations with T2D and either BMI, CKD, or NAFLD for the proteins identified as discordant using the PGS.

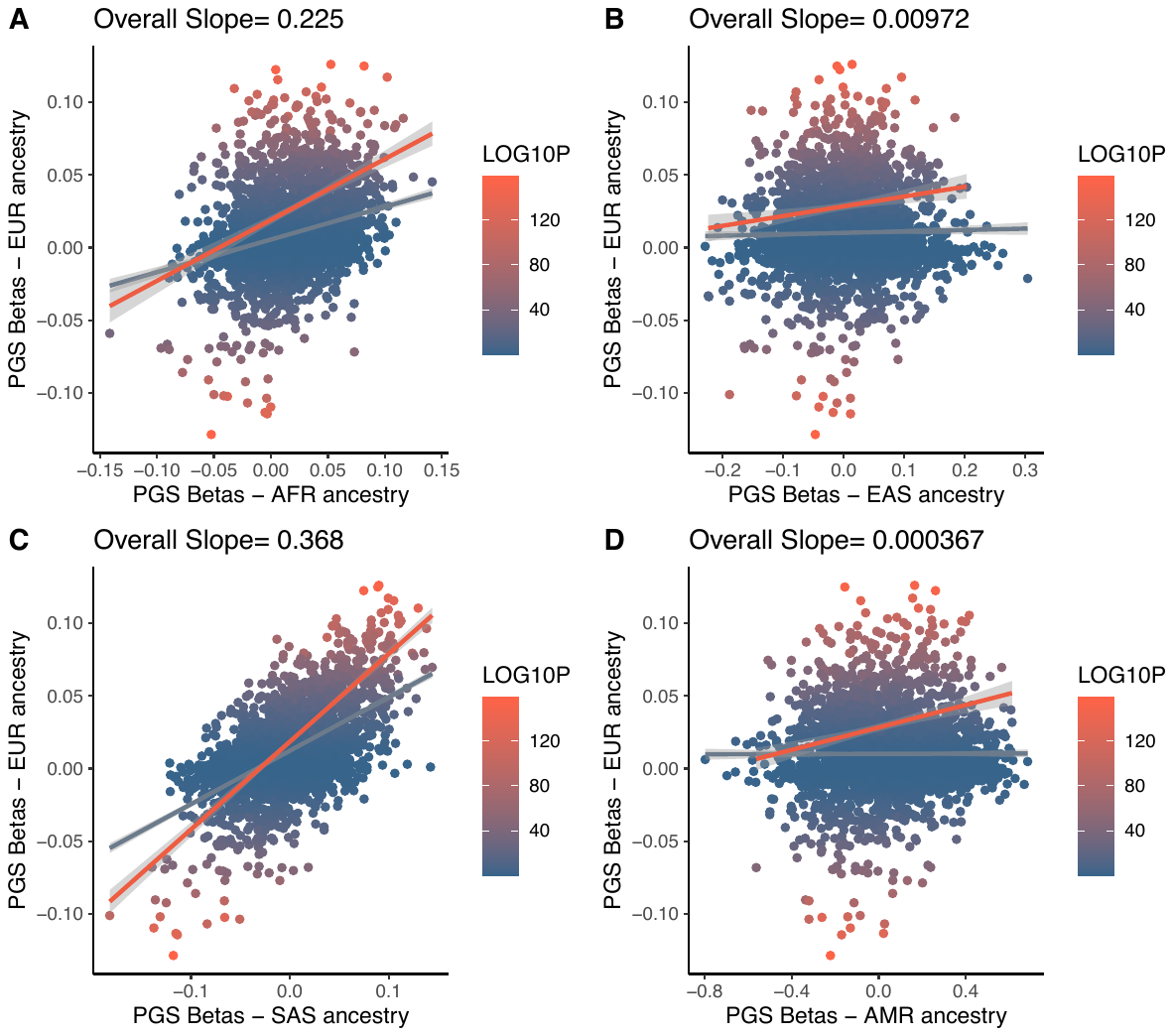

**Supplementary Figure 19: Cross-population comparison of PGS_T2D_gw_ effect sizes on circulating protein levels.** A. Beta-beta plot of PGS effect sizes in European ancestry (y-axis) and African ancestry (x-axis) UKB participants. The regression line for all proteins is silver and the regression line for only replicated proteins is in red. B. Beta-beta plot of PGS effect sizes in European ancestry (y-axis) and East Asian ancestry (x-axis) UKB participants. The regression line for all proteins is silver and the regression line for only replicated proteins is in red. C. Beta-beta plot of PGS effect sizes in European ancestry (y-axis) and South Asian ancestry (x-axis) UKB participants. D. Beta-beta plot of PGS effect sizes in European ancestry (y-axis) and Admixed American (Latin American) ancestry (x-axis) UKB participants. The regression line for all proteins is silver and the regression line for only replicated proteins is in red. Note that all ancestries were predicted using the 1000 Genomes Project as references. Also, the East Asian and Latin American subsets were severely underpowered (B and D), likely impacting the results.

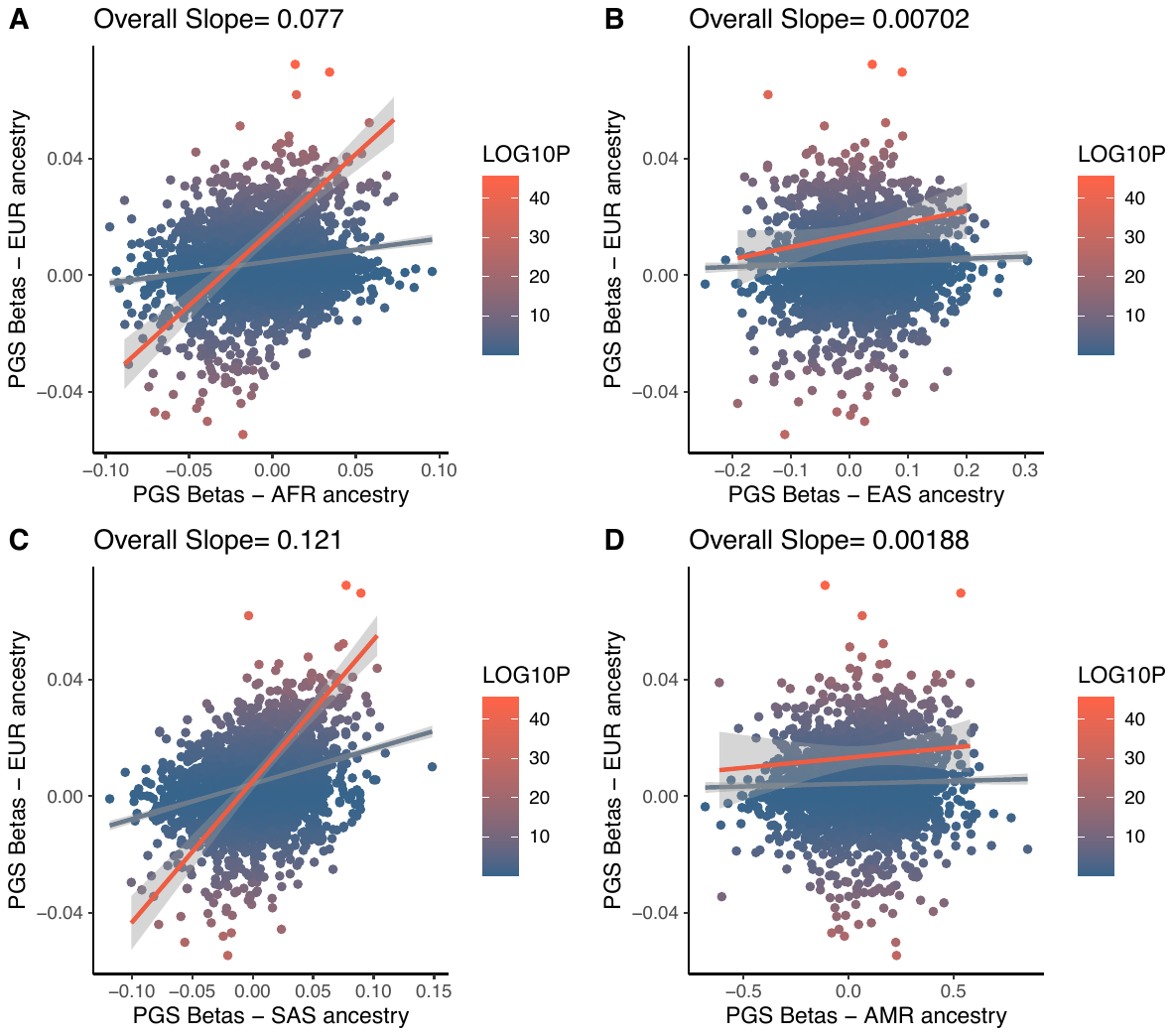

**Supplementary Figure 20: Cross-population comparison of PGS_CAD_ effect sizes on circulating protein levels.** A. Beta-beta plot of PGS effect sizes in European ancestry (y-axis) and African ancestry (x-axis) UKB participants. The regression line for all proteins is silver and the regression line for only replicated proteins is in red. B. Beta-beta plot of PGS effect sizes in European ancestry (y-axis) and East Asian ancestry (x-axis) UKB participants. The regression line for all proteins is silver and the regression line for only replicated proteins is in red. C. Beta-beta plot of PGS effect sizes in European ancestry (y-axis) and South Asian ancestry (x-axis) UKB participants. D. Beta-beta plot of PGS effect sizes in European ancestry (y-axis) and Admixed American (Latin American) ancestry (x-axis) UKB participants. The regression line for all proteins is silver and the regression line for only replicated proteins is in red. Note that all ancestries were predicted using the 1000 Genomes Project as references. Also, the East Asian and Latin American subsets were severely underpowered (B and D), likely impacting the results.

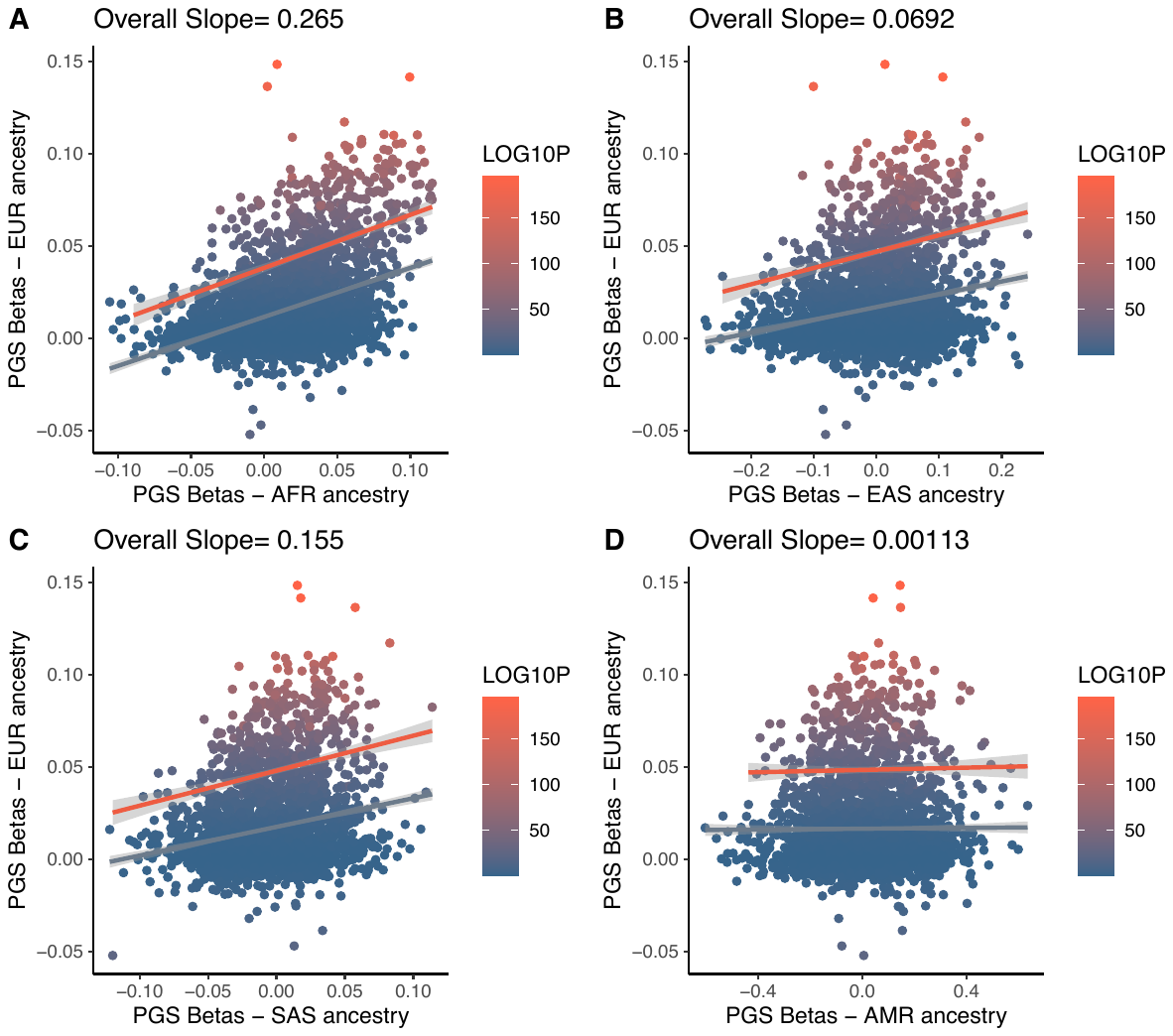

**Supplementary Figure 21: Cross-population comparison of PGS_CKD_ effect sizes on circulating protein levels.** A. Beta-beta plot of PGS effect sizes in European ancestry (y-axis) and African ancestry (x-axis) UKB participants. The regression line for all proteins is silver and the regression line for only replicated proteins is in red. B. Beta-beta plot of PGS effect sizes in European ancestry (y-axis) and East Asian ancestry (x-axis) UKB participants. The regression line for all proteins is silver and the regression line for only replicated proteins is in red. C. Beta-beta plot of PGS effect sizes in European ancestry (y-axis) and South Asian ancestry (x-axis) UKB participants. D. Beta-beta plot of PGS effect sizes in European ancestry (y-axis) and Admixed American (Latin American) ancestry (x-axis) UKB participants. The regression line for all proteins is silver and the regression line for only replicated proteins is in red. Note that all ancestries were predicted using the 1000 Genomes Project as references. Also, the East Asian and Latin American subsets were severely underpowered (B and D), likely impacting the results.

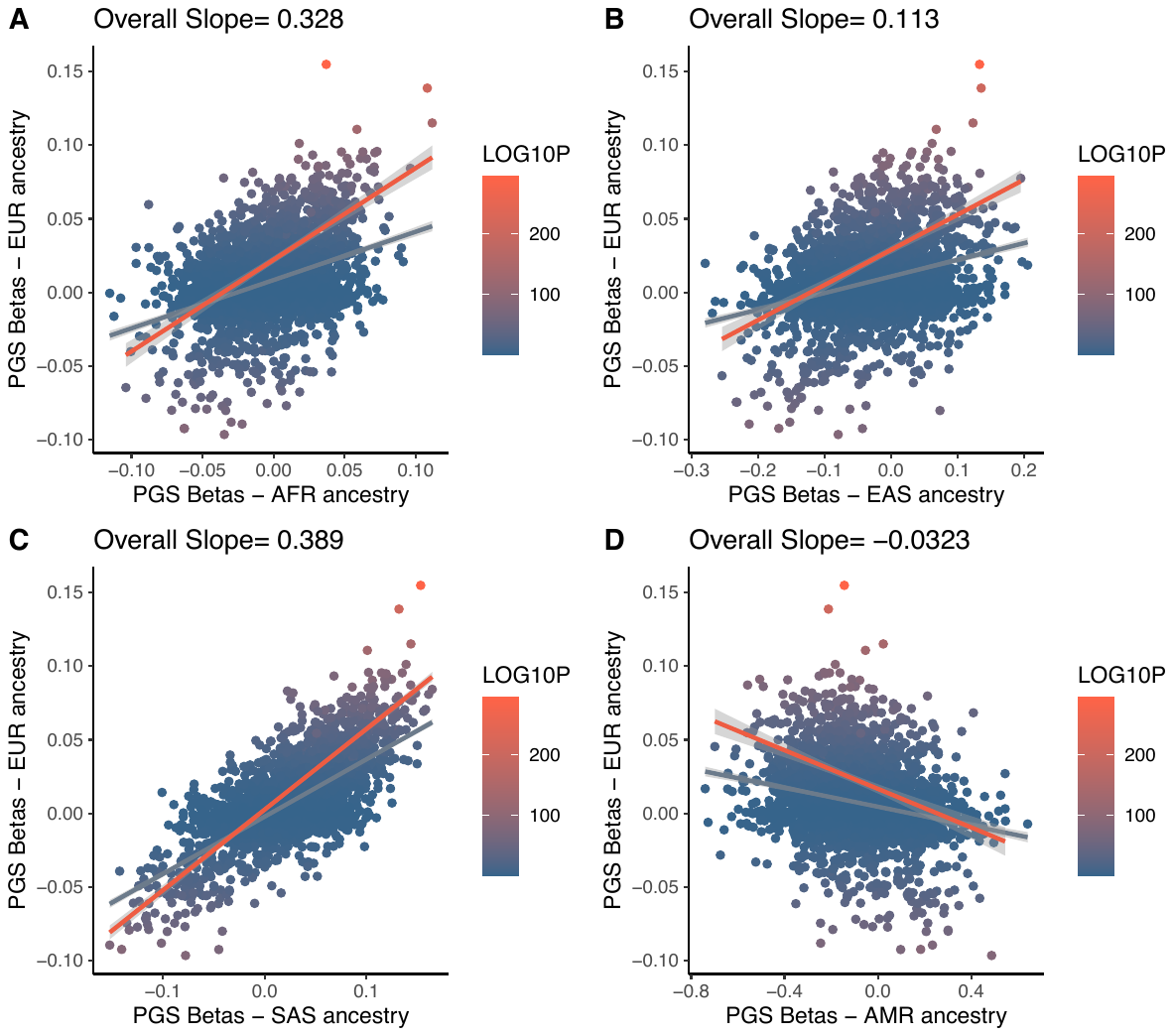

**Supplementary Figure 22: Cross-population comparison of PGS_BMI_ effect sizes on circulating protein levels.** A. Beta-beta plot of PGS effect sizes in European ancestry (y-axis) and African ancestry (x-axis) UKB participants. The regression line for all proteins is silver and the regression line for only replicated proteins is in red. B. Beta-beta plot of PGS effect sizes in European ancestry (y-axis) and East Asian ancestry (x-axis) UKB participants. The regression line for all proteins is silver and the regression line for only replicated proteins is in red. C. Beta-beta plot of PGS effect sizes in European ancestry (y-axis) and South Asian ancestry (x-axis) UKB participants. D. Beta-beta plot of PGS effect sizes in European ancestry (y-axis) and Admixed American (Latin American) ancestry (x-axis) UKB participants. The regression line for all proteins is silver and the regression line for only replicated proteins is in red. Note that all ancestries were predicted using the 1000 Genomes Project as references. Also, the East Asian and Latin American subsets were severely underpowered (B and D), likely impacting the results.

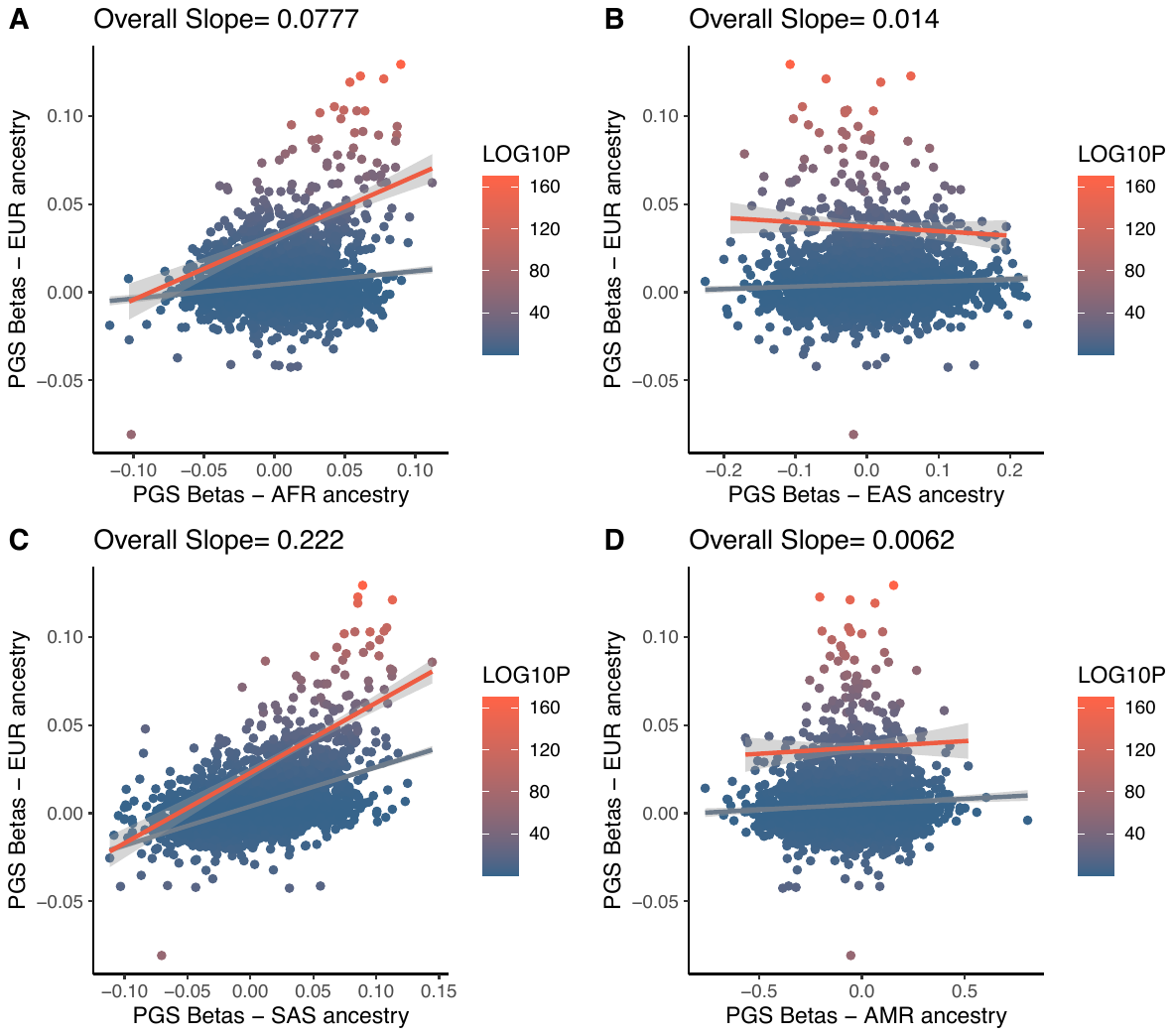

**Supplementary Figure 23: Cross-population comparison of PGS_NAFLD_ effect sizes on circulating protein levels**. A. Beta-beta plot of PGS effect sizes in European ancestry (y-axis) and African ancestry (x-axis) UKB participants. The regression line for all proteins is silver and the regression line for only replicated proteins is in red. B. Beta-beta plot of PGS effect sizes in European ancestry (y-axis) and East Asian ancestry (x-axis) UKB participants. The regression line for all proteins is silver and the regression line for only replicated proteins is in red. C. Beta-beta plot of PGS effect sizes in European ancestry (y-axis) and South Asian ancestry (x-axis) UKB participants. D. Beta-beta plot of PGS effect sizes in European ancestry (y-axis) and Admixed American (Latin American) ancestry (x-axis) UKB participants. The regression line for all proteins is silver and the regression line for only replicated proteins is in red. Note that all ancestries were predicted using the 1000 Genomes Project as references. Also, the East Asian and Latin American subsets were severely underpowered (B and D), likely impacting the results.

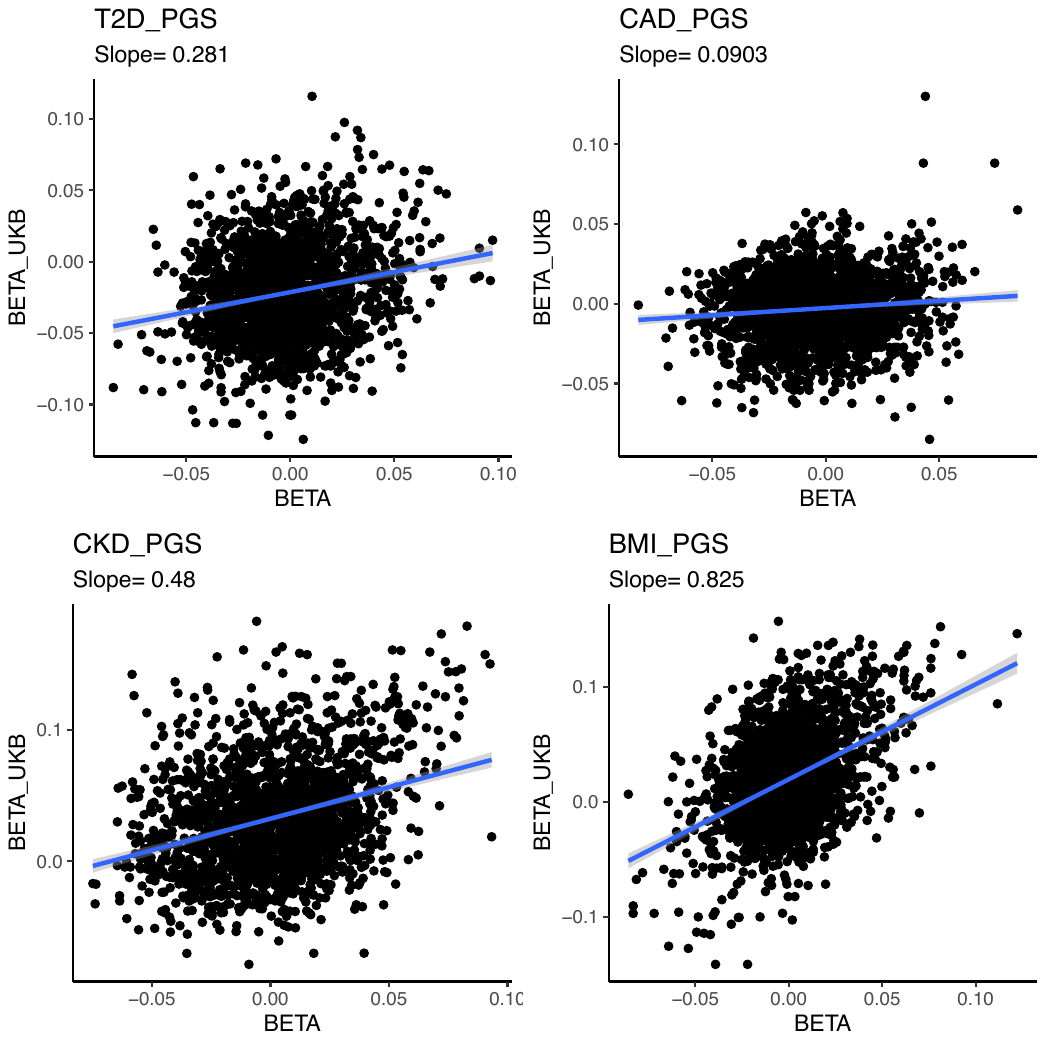

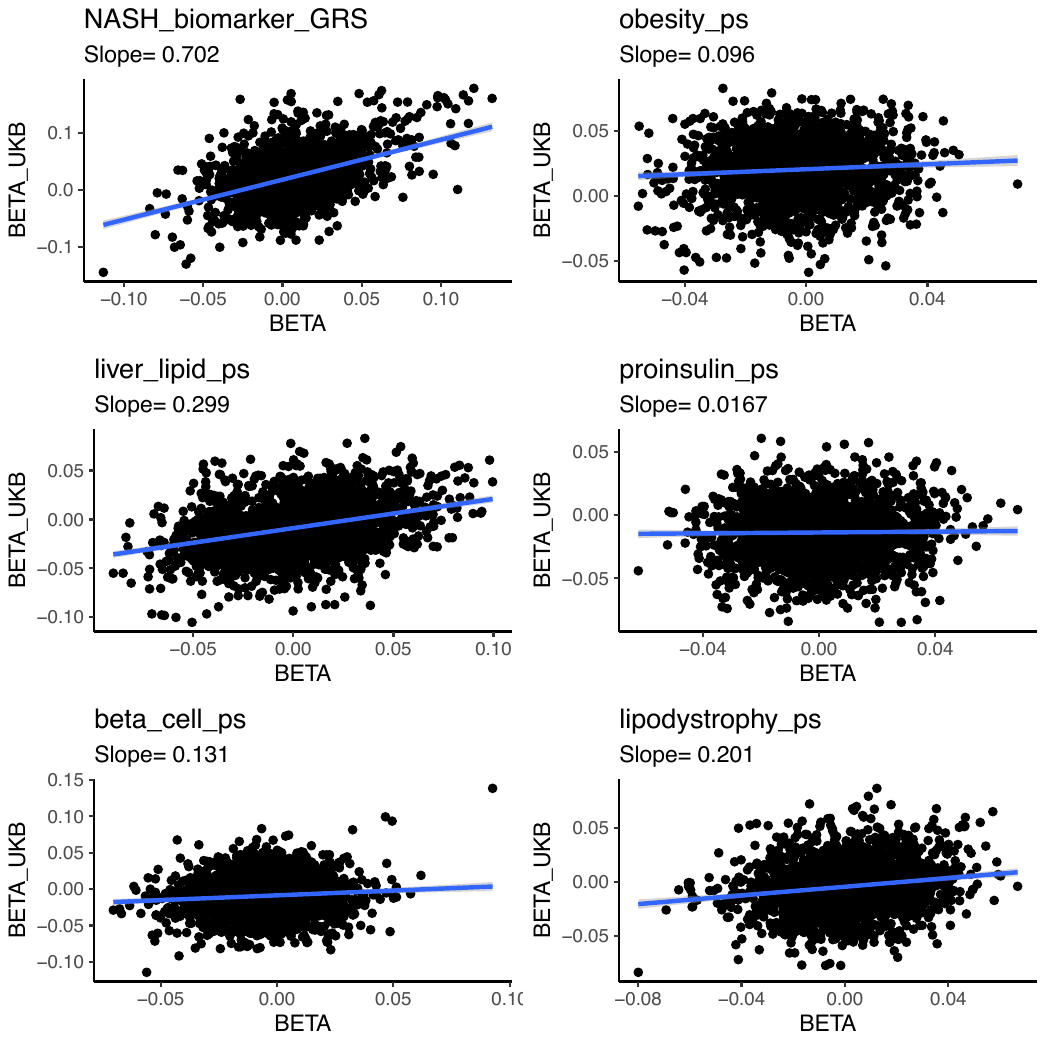

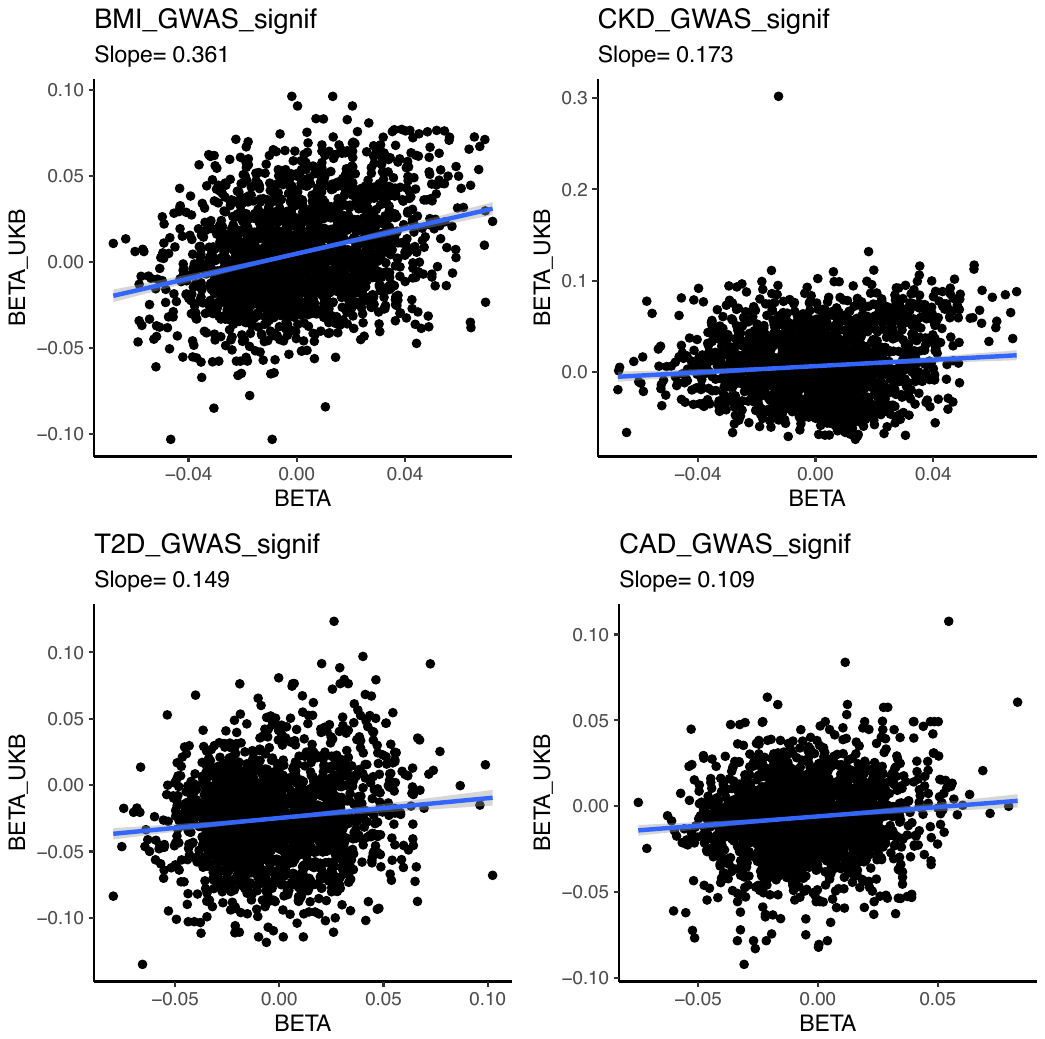

**Supplementary Figure 24: Beta-beta plot of PGS effect sizes on circulating proteins in EXSCEL vs UKB patients with diabetes.** Plots comparing beta coefficients for each PGS/pPS evaluating this study in EXSCEL with that of the UKB-PPP patients with diabetes. The regression line is in blue.

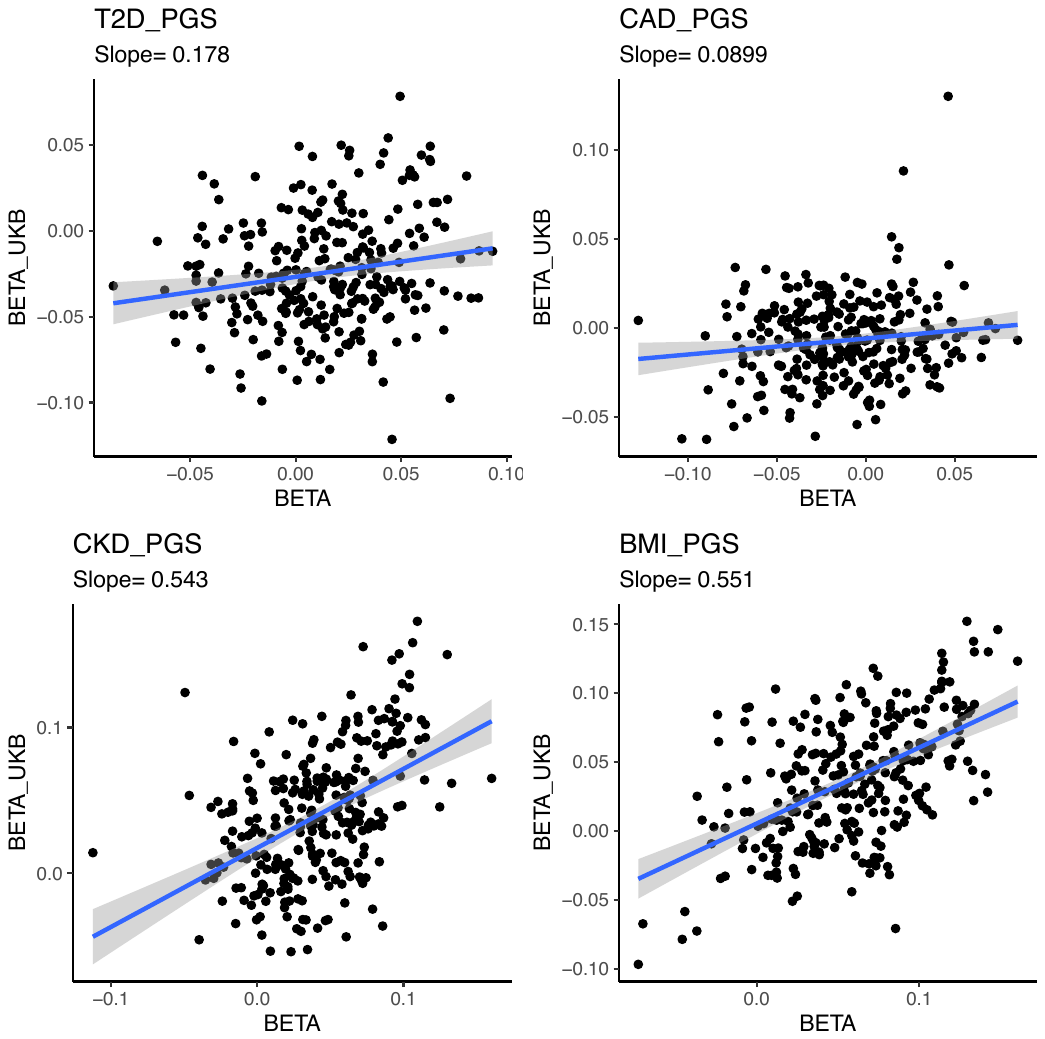

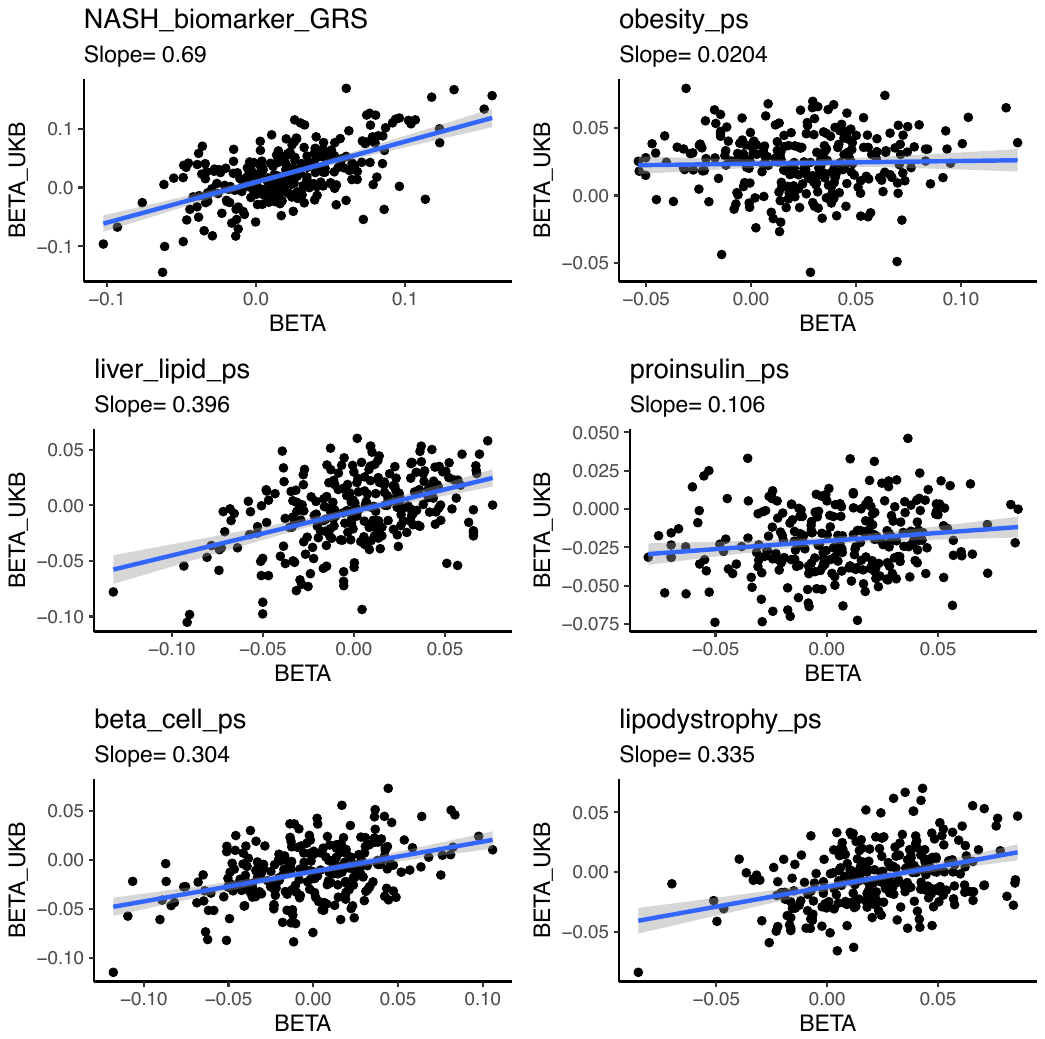

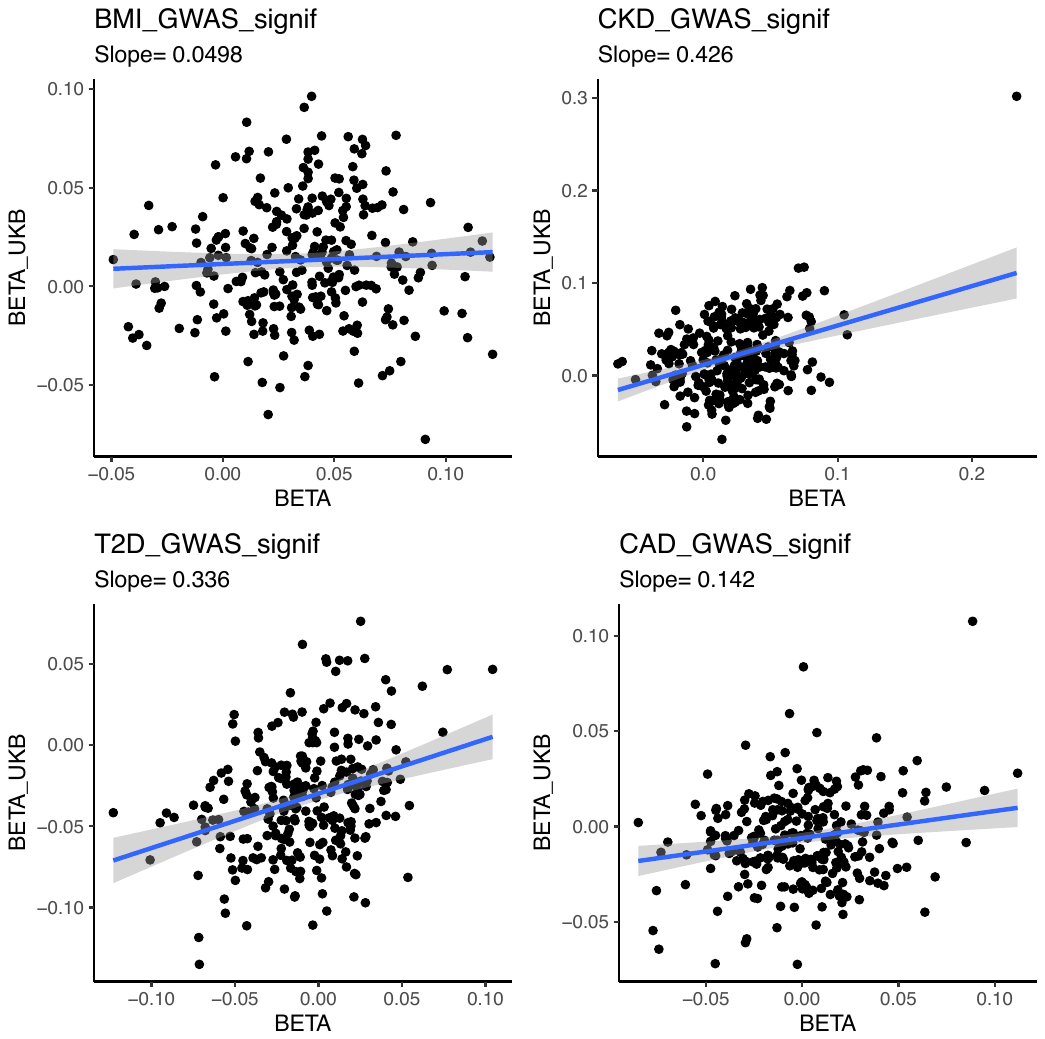

**Supplementary Figure 25: Beta-beta plot of PGS effect sizes on circulating proteins in DECLARE vs UKB-PPP patients with diabetes.** Plots comparing beta coefficients for each PGS/pPS evaluating this study in DECLARE with that of the UKB-PPP patients with diabetes. The regression line is in blue.

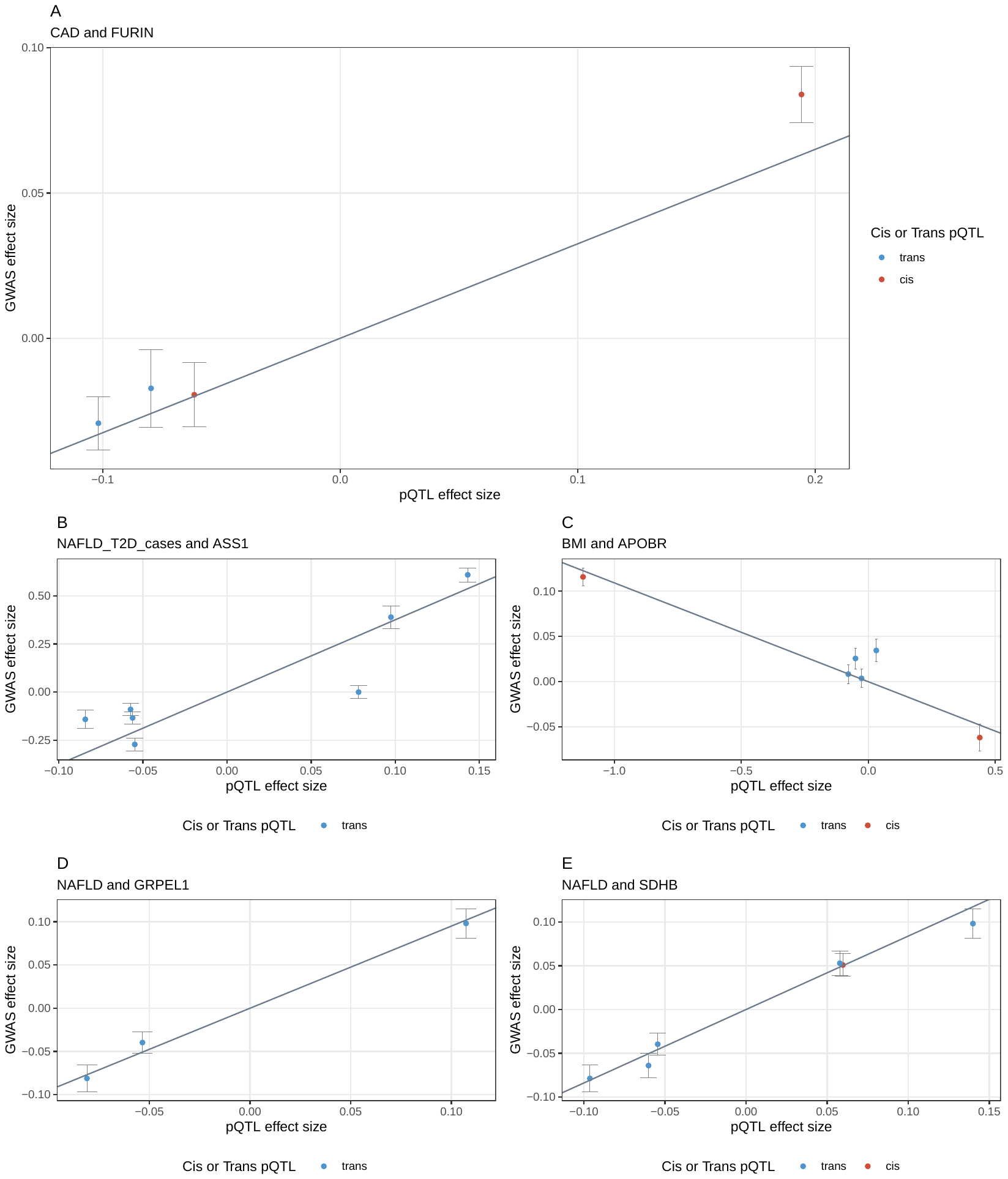

**Supplementary Figure 26: MR associations using *trans* pQTLs.** All panels display the pQTL effect size on the x-axis (beta coefficient) with trait GWAS effect size (beta coefficient) on the y-axis. Error bars are the standard error of the GWAS beta coefficient, while the slope is the ensemble MR estimate (median of IVW, weighted IVW, median, and Egger methods). Points are genetic instruments, with blue points indicated *trans* pQTLs and red indicating *cis* pQTLs. The protein/train combination is indicated in the subtitle.

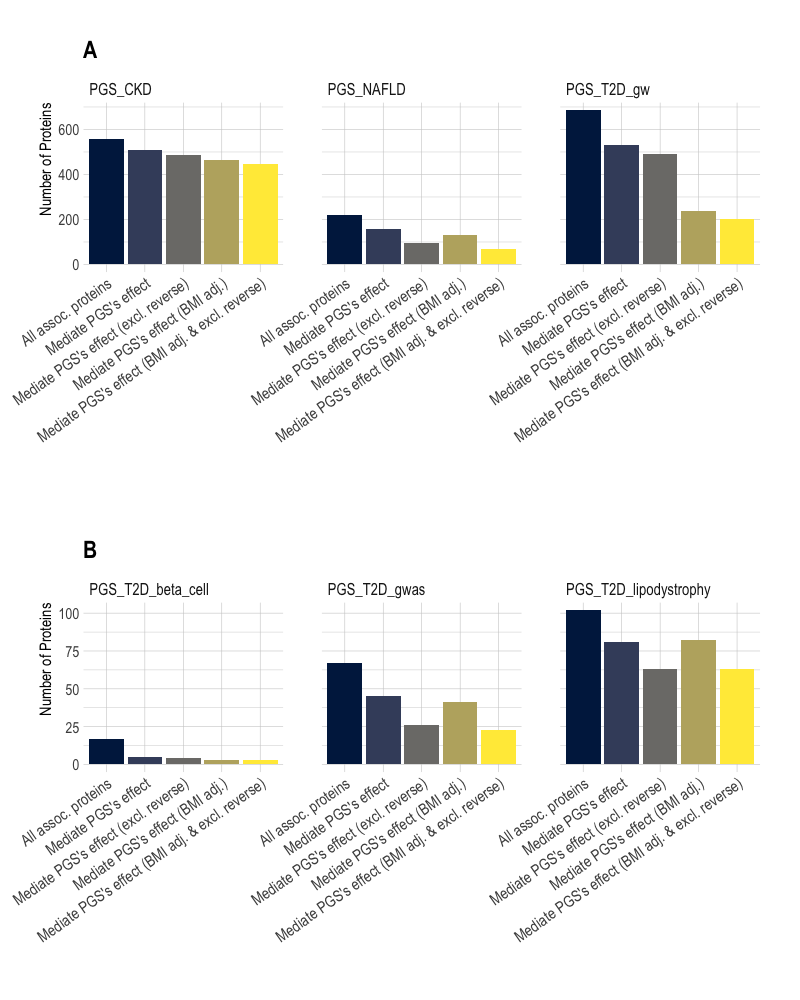

**Supplementary Figure 27: Mediation analysis summary.** From the left, the first column in each bar plot is the total number of proteins associated with a PGS, followed by the total number of proteins that significantly mediate the PGS’s effect on incident disease (CKD, T2D, CAD, or NAFLD), the number of proteins that significantly mediate the PGS’s effect on incident disease after excluding proteins found using reverse MR for the corresponding trait, the total number of proteins that significantly mediate the PGS’s effect on incident disease after BMI adjustment, and the total number of proteins that significantly mediate the PGS’s effect on incident disease after BMI adjustment and reverse MR exclusions. A: Results for cardiometabolic PGS including PGS_T2D_gw_. B: Results for GWAS-significant T2D PGS, including the partitioned polygenic scores. Note that if a partitioned polygenic score was not shown, then it was not significantly associated with either incident T2D or not significantly associated with any proteins.

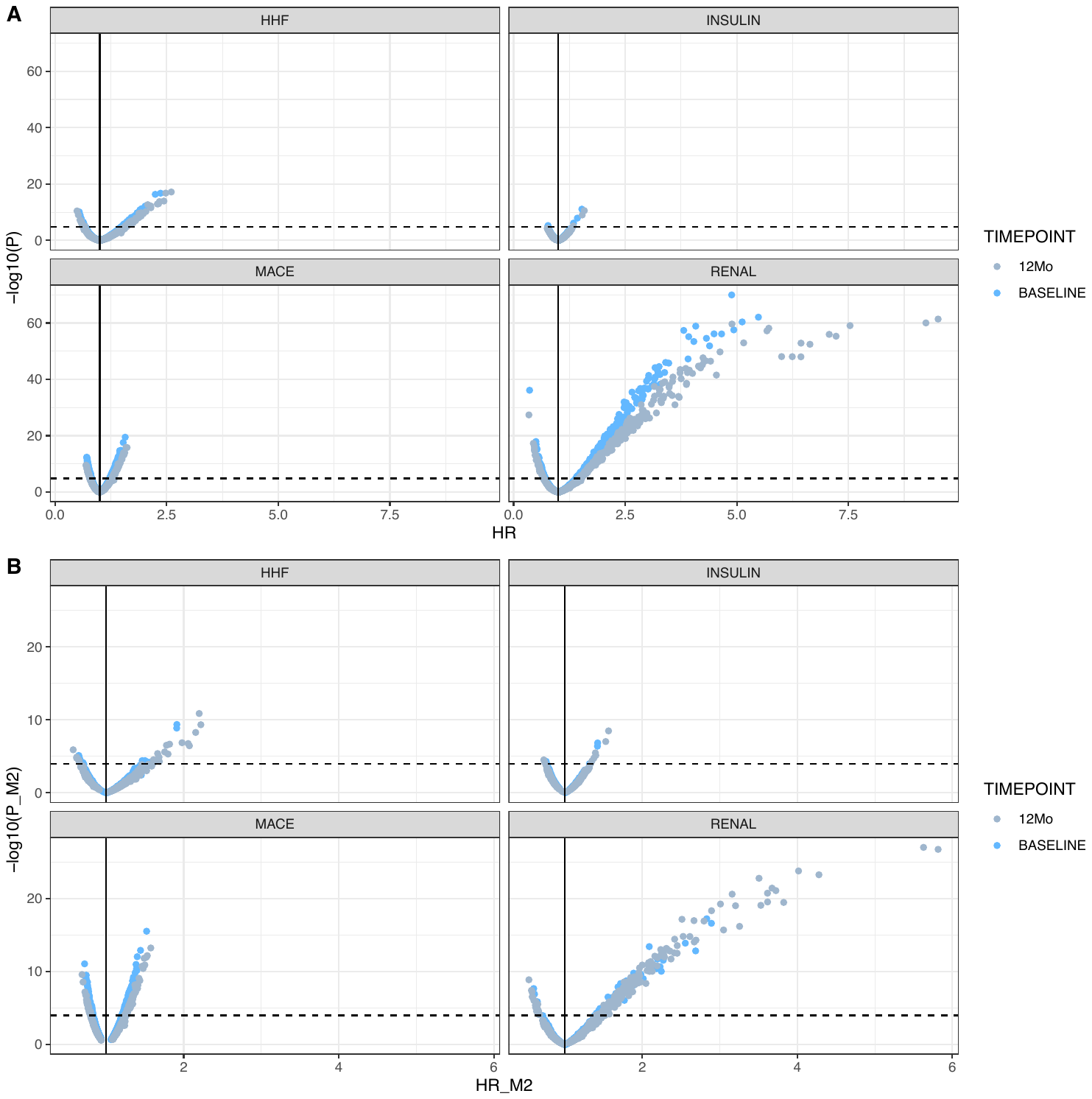

**Supplementary Figure 28: Protein associations with clinical trial outcomes in EXSCEL. A.** PGS-associated proteins were tested for association with four clinical trial endpoints in EXSCEL using Cox proportional hazards regression and standard covariates (e.g., age, sex, PCs). Blue points represent the baseline protein measurement while grey points represent the 12-month timepoint. **B.** PGS-associated proteins were tested for association with four clinical trial endpoints in EXSCEL using Cox proportional hazards regression and standard covariates (e.g., age, sex, PCs) plus outcome specific risk factors. Blue points represent the baseline protein measurement while grey points represent the 12-month timepoint.

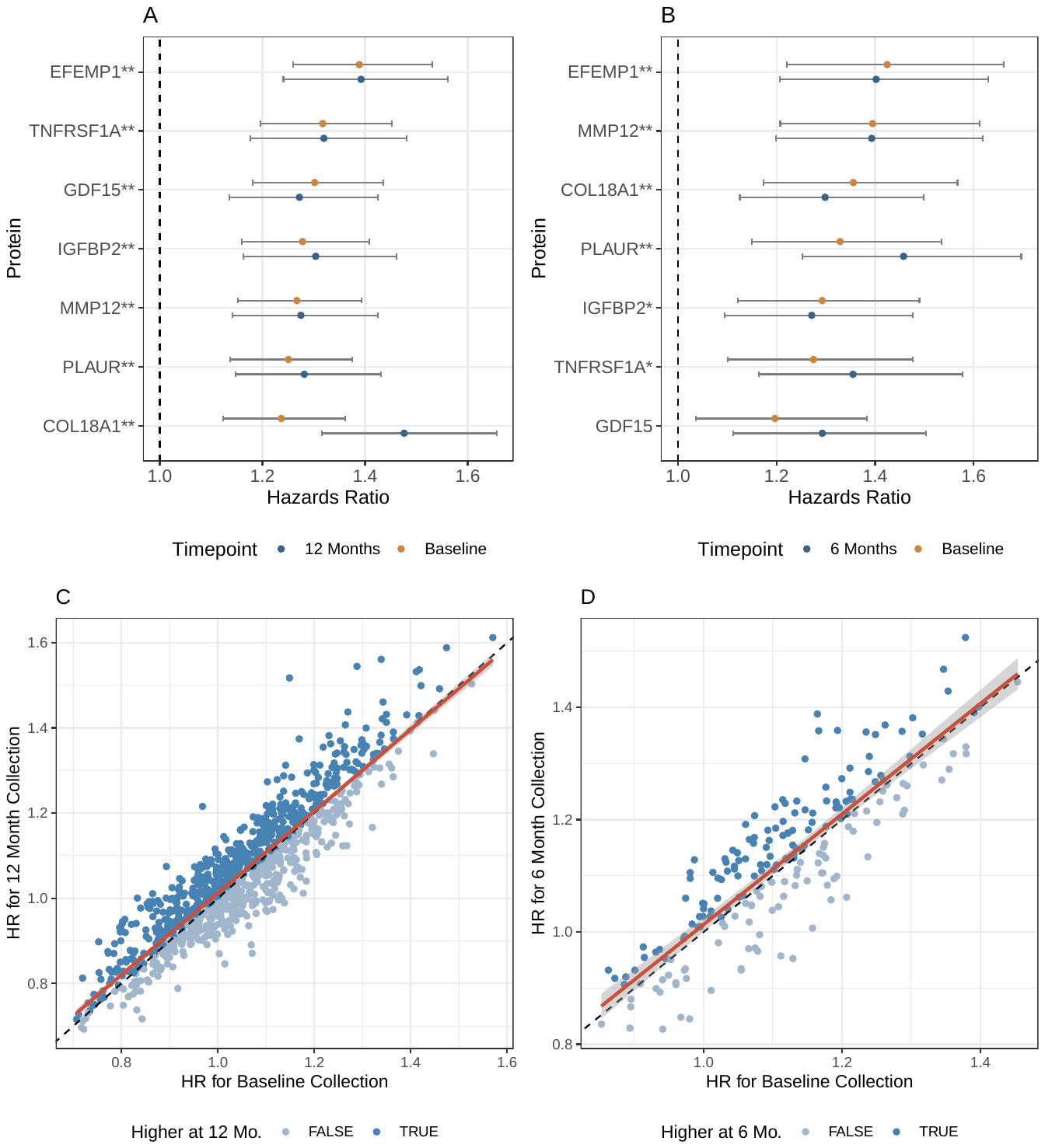

**Supplementary Figure 29: Biomarker analysis for MACE in EXSCEL and DECLARE.** *MACE outcome biomarkers in randomized controlled trials discovered in EXSCEL and replicated in DECLARE-TIMI 58. All proteins displayed here were significantly associated with MACE in EXSCEL after adjusting for clinical factors, were available in DECLARE, and were nominally associated using the demographics-only model. A. Hazard ratios of the proteins associated with EXSCEL’s MACE endpoint after adjusting for clinical risk factors, with yellow indicating the proteins were measured at baseline and blue indicating the proteins were measured as 12 months. Proteins with a “**” were significant at both timepoints, while proteins with a “*” were significant at only one timepoint. B. Hazard ratios of the proteins associated with DECLARE’s MACE endpoint after adjusting for clinical risk factors, with yellow indicating the proteins were measured at baseline and blue indicating the proteins were measured as 6 months. Proteins with a “**” significantly replicated at both time points and proteins with a “*” significantly replicated at one timepoint. C. Scatterplot of hazards ratios (unadjusted for clinical risk factors) at baseline (x-axis) and 12 months (y-axis) for EXSCEL. The regression line is in red, while the dashed black line indicates where the hazard ratios would be in complete concordance. D. Scatterplot of hazards ratios (unadjusted for clinical risk factors) at baseline (x-axis) and 6 months (y-axis) for DECLARE. The regression line is in red, while the dashed black line indicates where the hazard ratios would be in complete concordance.*

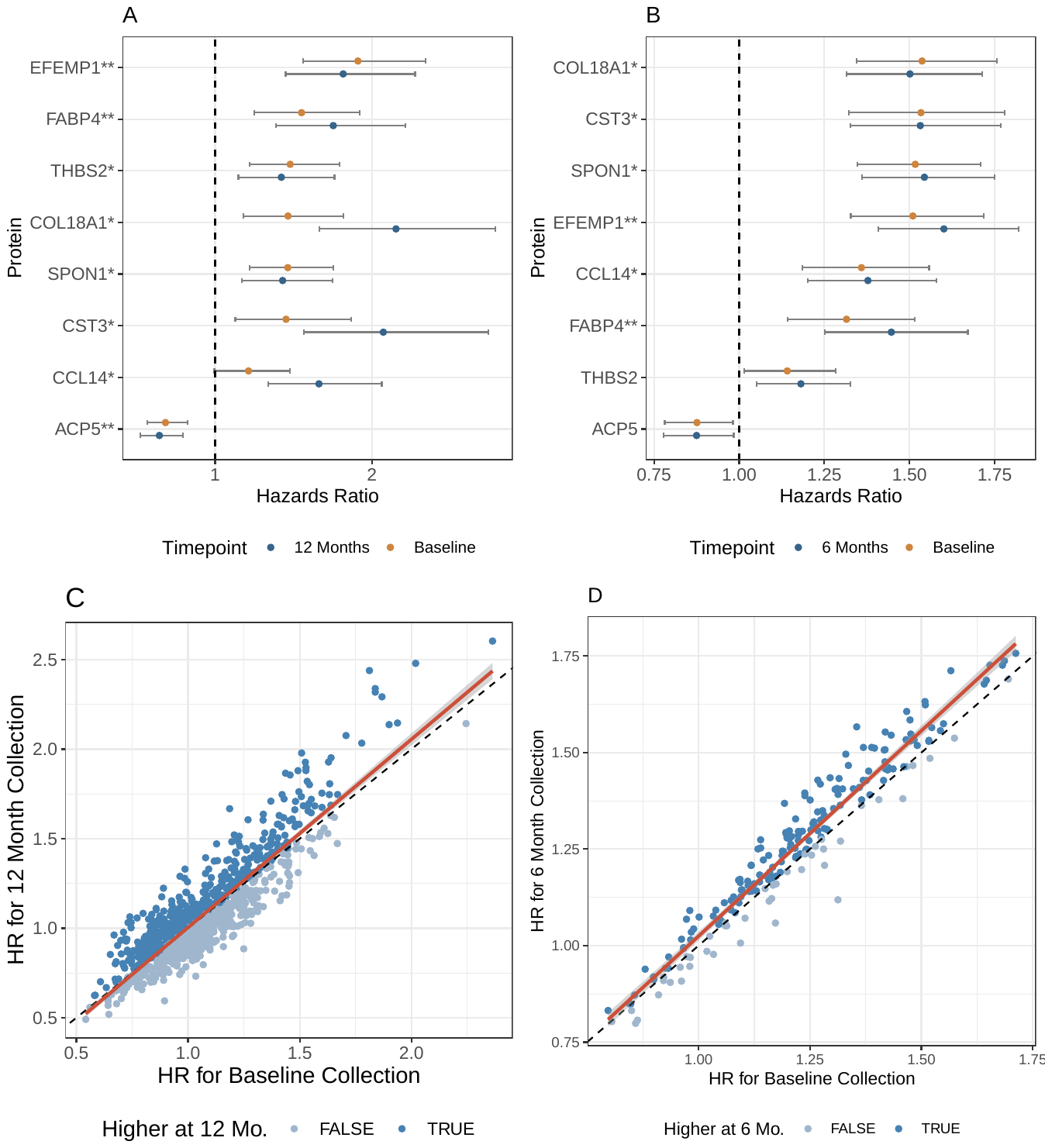

**Supplementary Figure 30: Biomarker analysis for Hospitalization for Heart Failure (HHF) in EXSCEL and DECLARE.** *HHF outcome biomarkers in randomized controlled trials discovered in EXSCEL and replicated in DECLARE-TIMI 58. All proteins displayed here were significantly associated with HHF in EXSCEL after adjusting for clinical factors, were available in DECLARE, and were nominally associated using the demographics-only model. A. Hazard ratios of the proteins associated with EXSCEL’s HHF endpoint after adjusting for clinical risk factors, with yellow indicating the proteins were measured at baseline and blue indicating the proteins were measured as 12 months. Proteins with a “**” were significant at both timepoints, while proteins with a “*” were significant at only one timepoint. B. Hazard ratios of the proteins associated with DECLARE’s HHF endpoint after adjusting for clinical risk factors, with yellow indicating the proteins were measured at baseline and blue indicating the proteins were measured as 6 months. Proteins with a “**” significantly replicated at both time points and proteins with a “*” significantly replicated at one timepoint. C. Scatterplot of hazards ratios (unadjusted for clinical risk factors) at baseline (x-axis) and 12 months (y-axis) for EXSCEL. The regression line is in red, while the dashed black line indicates where the hazard ratios would be in complete concordance. D. Scatterplot of hazards ratios (unadjusted for clinical risk factors) at baseline (x-axis) and 6 months (y-axis) for DECLARE. The regression line is in red, while the dashed black line indicates where the hazard ratios would be in complete concordance.*

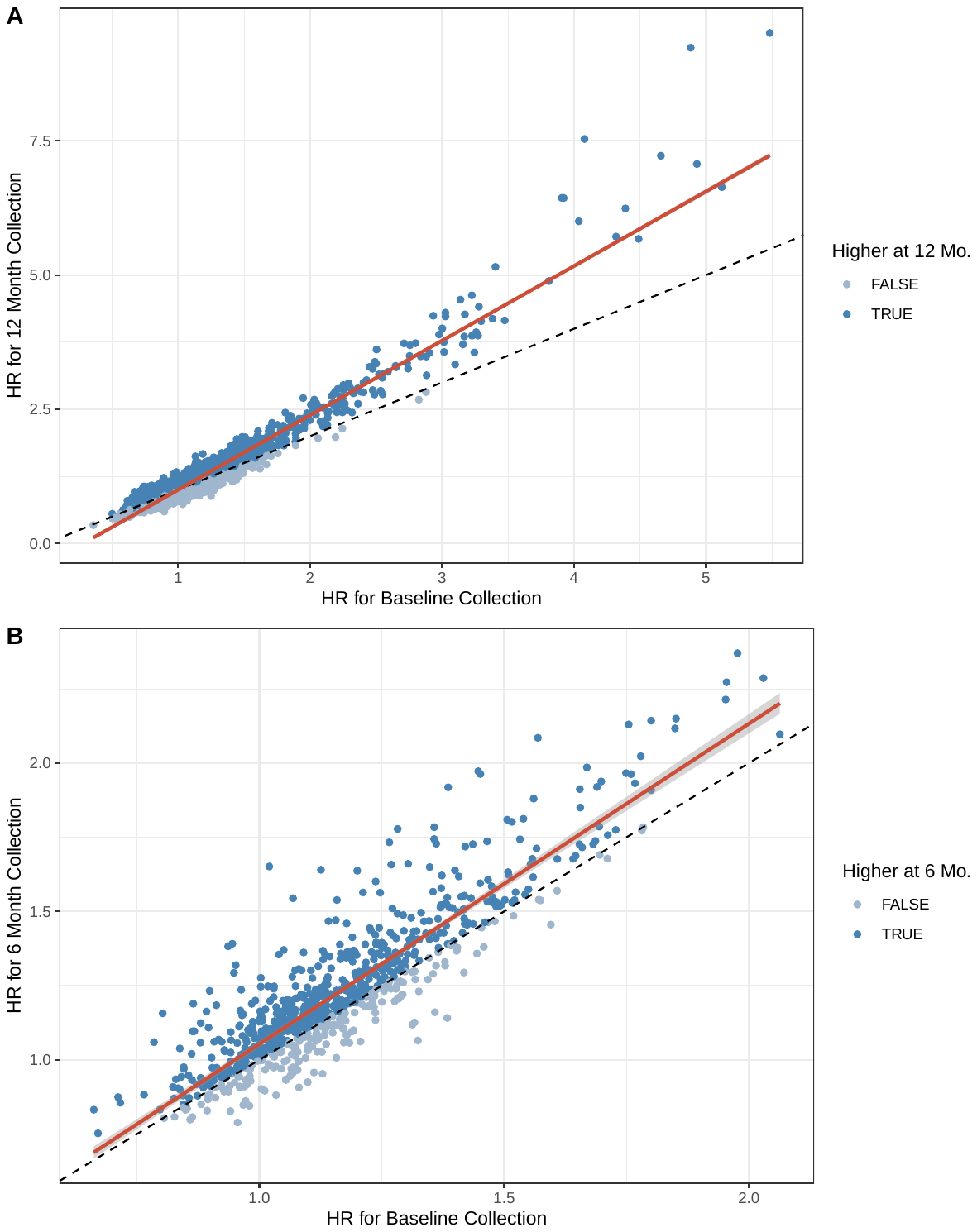

**Supplementary Figure 31: Hazards ratios at two different time points.** *A. Scatterplot of hazards ratios for all four outcomes (unadjusted for clinical risk factors) at baseline (x-axis) and 12 months (y-axis) for EXSCEL. The regression line is in red, while the dashed black line indicates where the hazard ratios would be in complete concordance. Hazard ratios were significantly different (t-test, p-value < 0.05). B. Scatterplot of hazards ratios for all four outcomes (unadjusted for clinical risk factors) at baseline (x-axis) and 6 months (y-axis) for DECLARE. The regression line is in red, while the dashed black line indicates where the hazard ratios would be in complete concordance. Hazard ratios were significantly different (t-test, p-value < 0.05).*

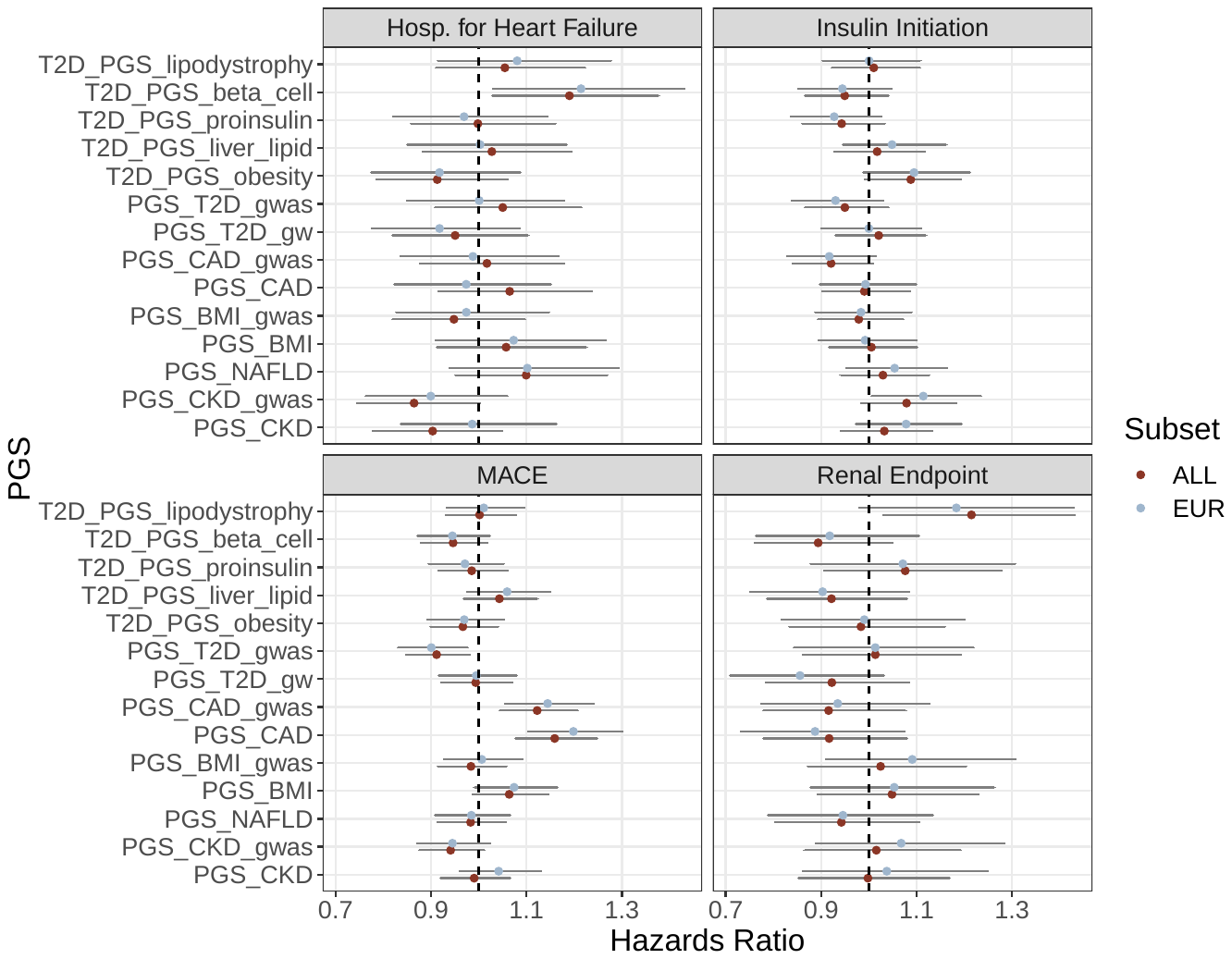

**Supplementary Figure 32: PGS associations in trial outcomes in EXSCEL.** The PGS were tested for association with four clinical trial endpoints in EXSCEL using Cox proportional hazards regression in order to determine their suitability for mediation analyses with the protein data.

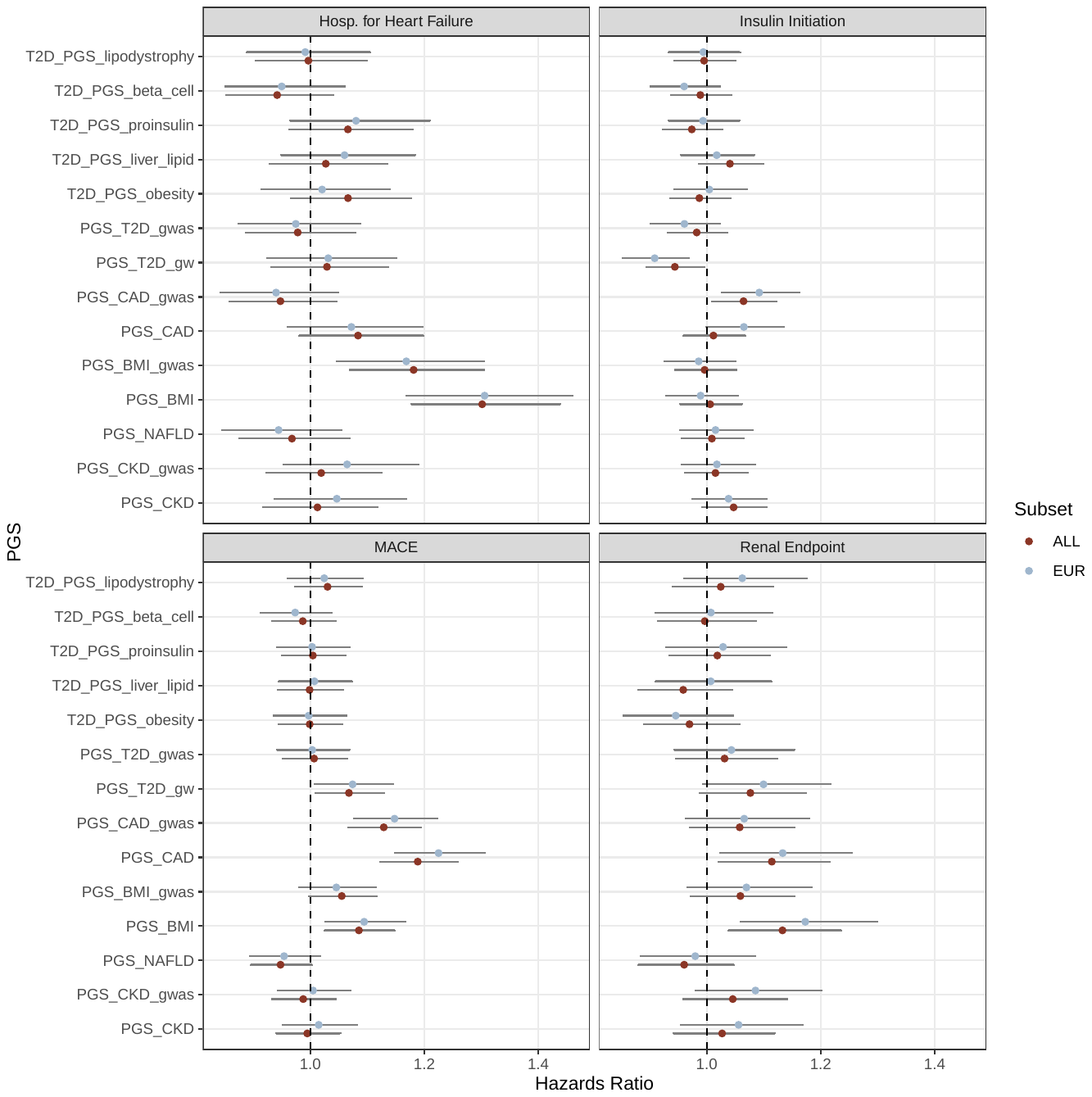

**Supplementary Figure 33: PGS associations with trial outcomes in DECLARE.** PGS tested for association with four clinical trial endpoints in DECLARE using Cox proportional hazards regression in order to determine their suitability for mediation analyses with the protein data.

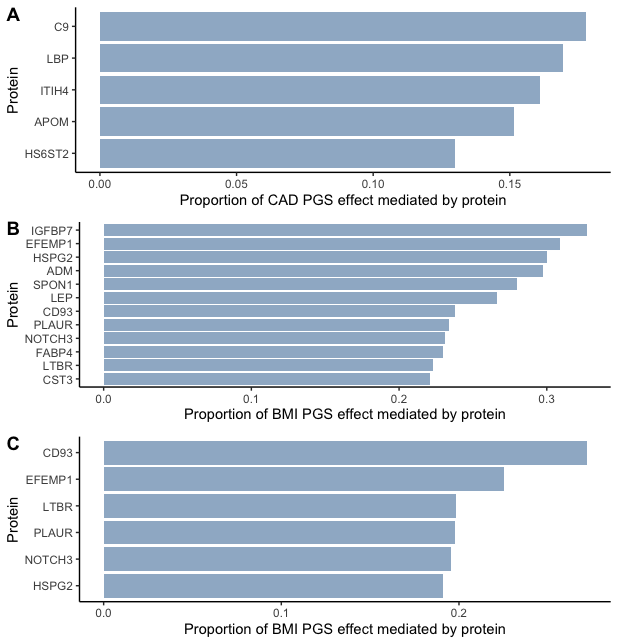

**Supplementary Figure 34: Mediation analysis in EXSCEL and DECLARE.** The mediation framework modelled the PGS as the direct effect, the protein as the indirect (mediating) effect, and the total effect (PGS + protein). The X-axis is the proportion of the PGS effect mediated by protein, defined as indirect effect (protein) / total effect (PGS + protein). Proteins in red were identified in our reverse MR analysis, suggesting the association was either due to reverse causality or a feedback mechanism. A. Mediation for CAD PGS and the MACE outcome in EXSCEL. B. Mediation for the BMI PGS and the hospitalization for heart failure endpoint. C. Mediation for the BMI PGS and the hospitalization for heart failure endpoint after adjusting for baseline BMI.

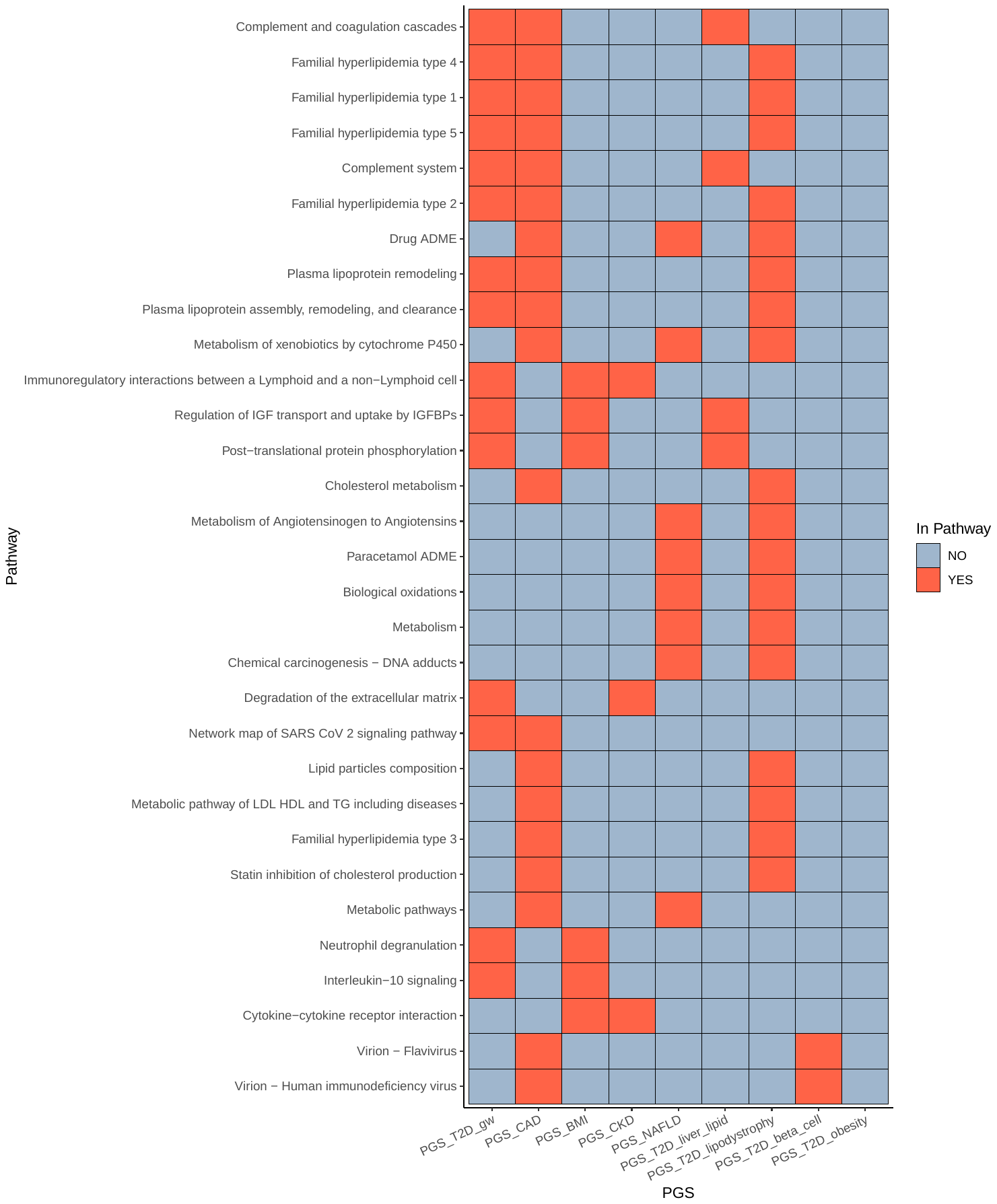

**Supplementary Figure 35: Path enrichment.** KEGG, REACTOME, and WIKIPATH pathways significantly enriched in the PGS-associated proteins. Red squares indicate if that pathway (y-axis) was also significantly enriched in the proteins associated with the given score (x-axis). Note that pathways enriched in two or more PGS-protein sets are shown here. A further 80 pathways were only enriched in one set, such as TNFs bind their physiological receptors for the PGS_CKD_.

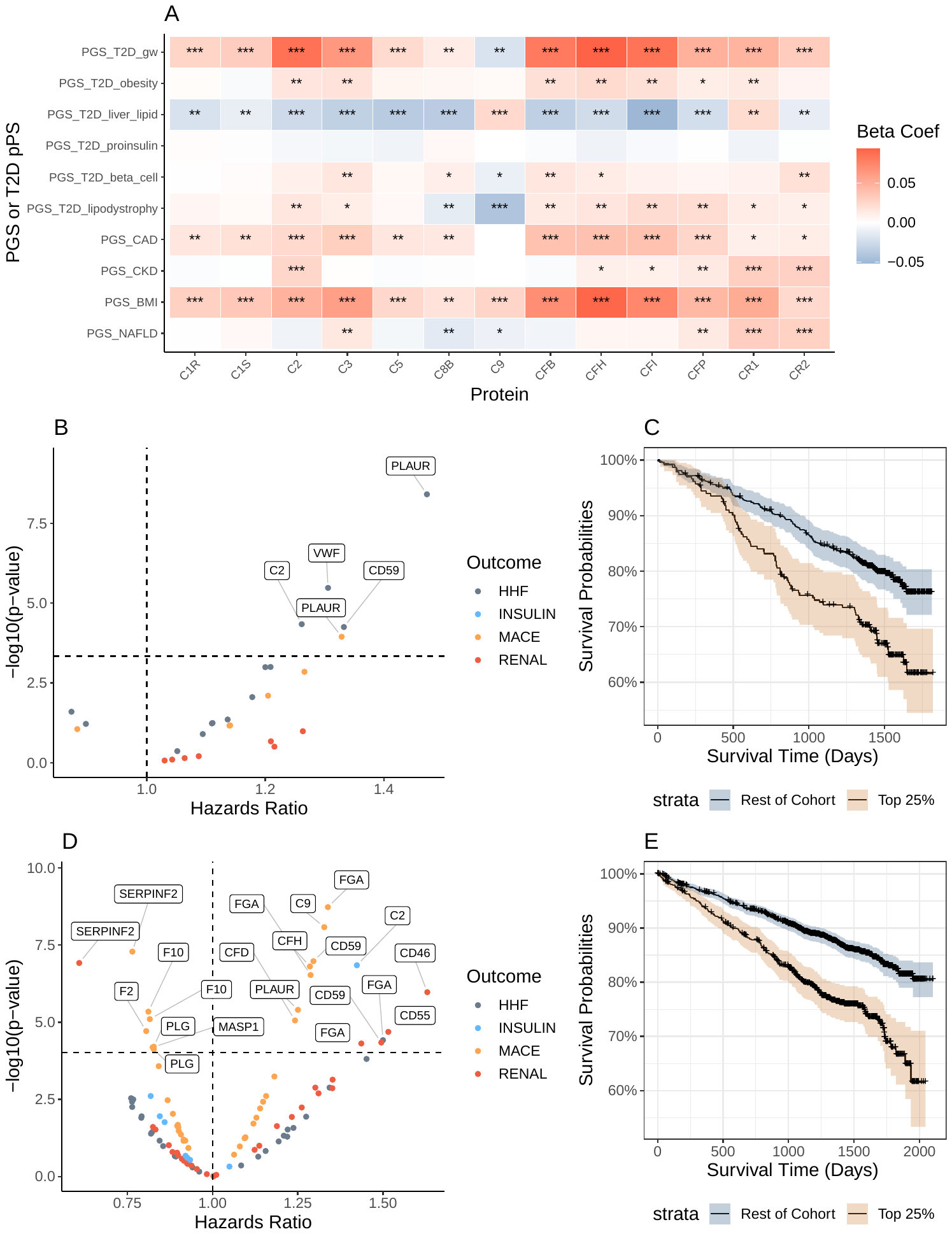

**Supplementary Figure 36: Complement and coagulation cascade pathway highlights.** A: PGS associations with complement-related proteins. A single asterisk (*) indicates the association was nominally significant, while two (**) indicates significance using FDR and three (***) indicates significance using a Bonferroni correction. B: Associations of proteins in this pathway with clinical trial outcomes in DECLARE using cox regression. C. Kaplan Meier curve demonstrating the impact of CD59 levels on the composite cardiovascular endpoint in DECLARE. D: Associations of proteins in this pathway with clinical trial outcomes in EXSCEL using cox regression. E. Kaplan Meier curve demonstrating the impact of CD59 levels on the composite cardiovascular endpoint in EXSCEL.

**A**

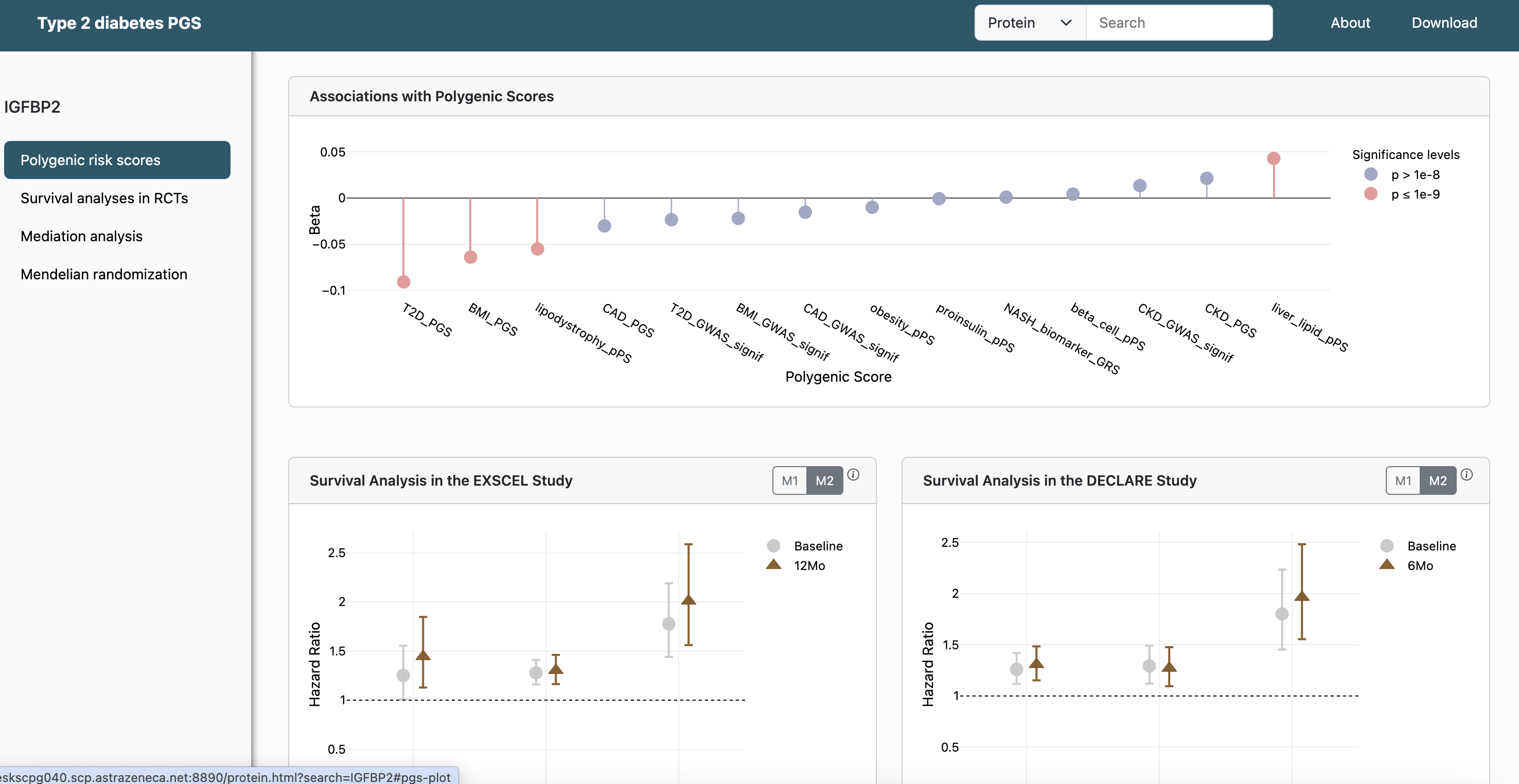

**B**

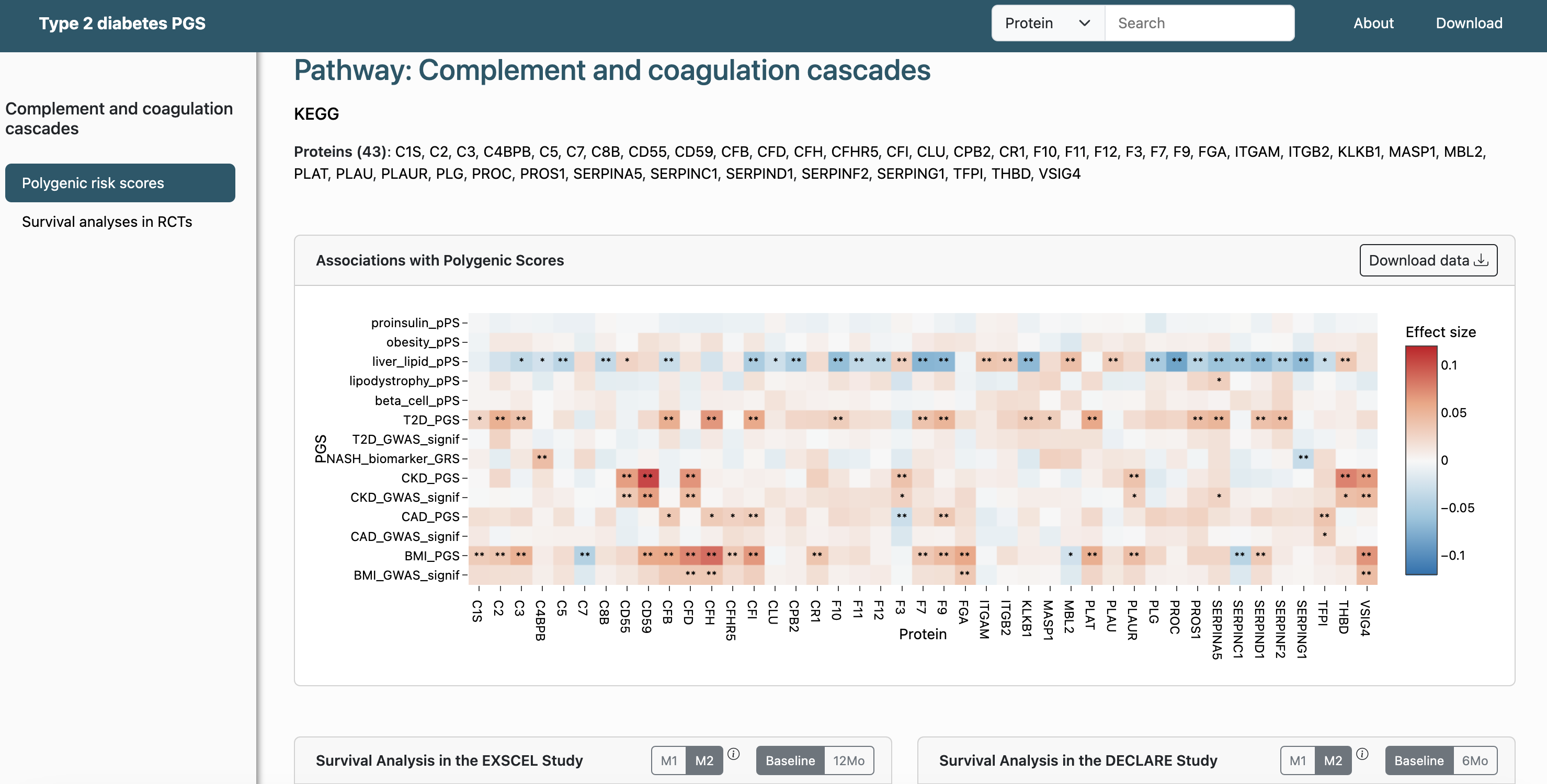

**C**

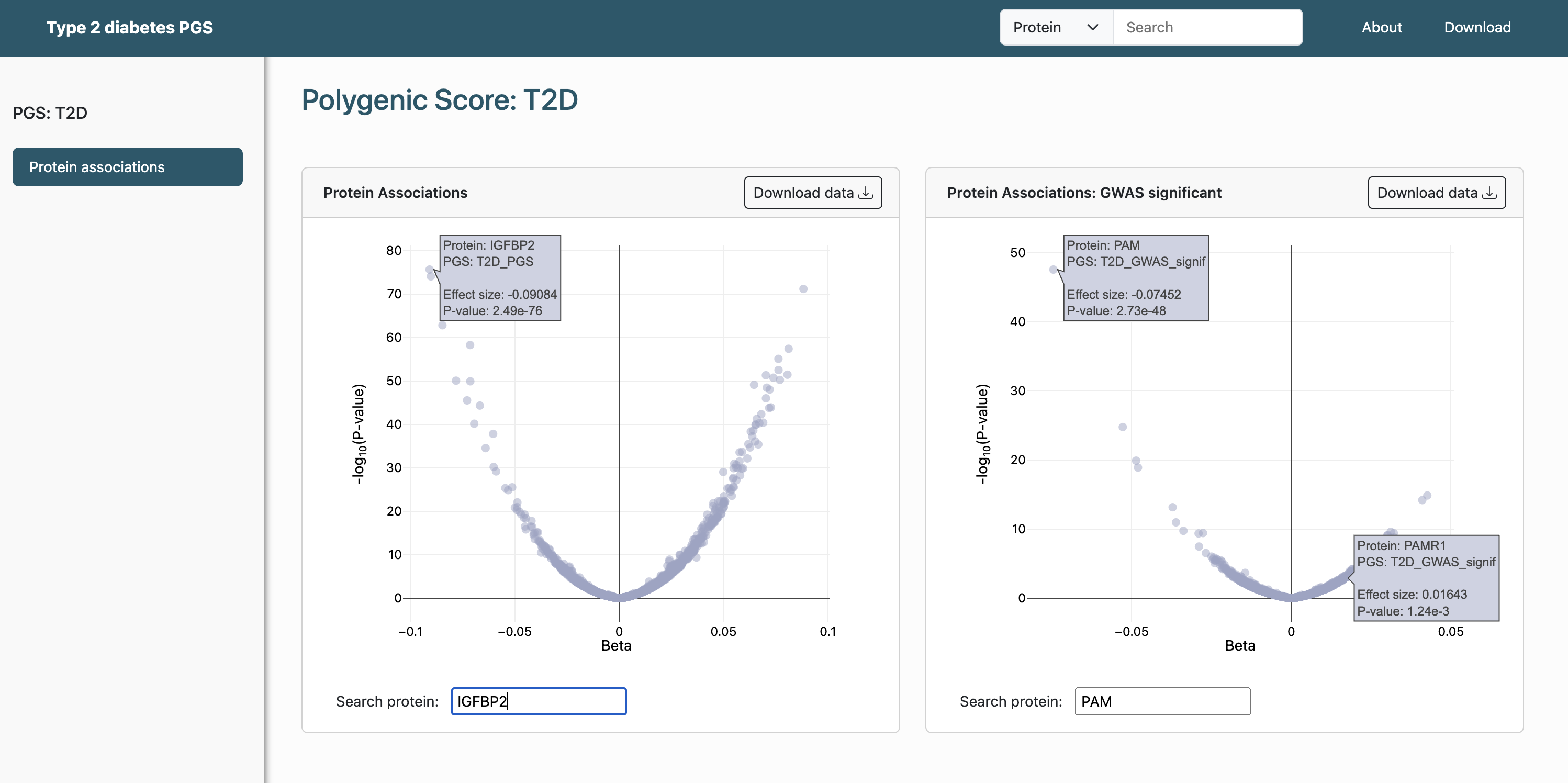

**Supplementary Figure 37: Example of Web App.** A: Example of querying by protein (IGFBP2). B: Example of querying by pathway (Complement and Coagulation Cascades). C: Example of querying by PGS (T2D).
